# Multi-agent LLM teams improve differential recall on curated cases but underperform on emergency working-label matching: a two-cohort, two-family evaluation

**DOI:** 10.64898/2026.09.24.26363897

**Authors:** Jun Feng, Yuhao Jiao, Huaiya Xie, Jiangshang Wang, Yiyao Li, Longxin Xie, Wenyu Peng, Xuefeng Sun

## Abstract

**Background:** Whether multi-agent large language model (LLM) pipelines help, add nothing, or underperform a single call may depend on the input and scoring contract rather than the architecture; these boundary conditions are unmapped.

**Methods:** We compared a single direct call (A×1), five personas in one context (P), and five isolated roles integrated by a moderator (MDT) across curated corpora (87 clinicopathological cases, 406 MedCaseReasoning cases, 392 replication vignettes) and two disjoint emergency cohorts from one repository (364 mixed-label encounters; a protocol-frozen 400-encounter disease-label-only replacement, one local serving stack). Scoring was judge-mediated top-k working-label matching. A completed factorial isolated the active ingredient; a symptom-permissive output-contract arm tested contract fit; three physicians rated discordant cases.

**Findings:** On curated corpora MDT improved differential-list recall (top-3: +5·5 points, p = 0·0079; +3·0, p = 0·0079; +3·3, p = 0·0013) but not top-1 accuracy; the factorial attributed the gain to independent generation plus moderated synthesis, not personas. On emergency presentations the sign reversed: in the disease-label-only cohort A×1 led MDT at every endpoint (top-3 −5·2 points, 95% CI −8·0 to −2·4, p = 0·0008), a second family reproduced it, and bridged cases excluded the serving stack. A symptom-permissive contract shrank the deficit’s point estimates by 52–75%, replacement-cohort intervals all spanning zero though remaining negative; the clause harmed no arm. Physician safety ratings were inconclusive (original-cohort gate missed; replacement-cohort rescue contrast unadjudicable, 95% CI −32·7 to +2·6).

**Interpretation:** Multi-agent diagnostic gains are real but conditional: the sign of the effect tracks input genre, and its magnitude tracks output-contract fit. Curated-benchmark gains should not be extrapolated to first-contact emergency text, where role-based teams underperform a single call at seven-fold token cost. The reversal is label-level and judge-mediated; clinical safety is neither established nor excluded.

**Funding:** None.

**Research in context:** *Evidence before this study:* Multi-agent LLM diagnostic pipelines report gains, but specialist personas, independent generation, synthesis, and additional test-time compute are usually varied together, so the active ingredient is unidentified. Evaluation is concentrated on curated case reports, leaving performance on first-contact notes and working-diagnosis labels unclear, and no study has tested whether the output contract demanded of the model contributes to measured underperformance.

*Added value of this study:* Across curated corpora and two disjoint emergency cohorts, we show the multi-agent effect is conditional and map two of its moderators. A completed factorial attributes the curated gain to independent generation plus moderated synthesis, not personas, and confines it to differential-list recall. The emergency deficit is established on a protocol-frozen cohort whose labels are all disease-level diagnoses, reproduced by a second model family on the same cases, and insensitive to the serving stack on bridged cases—so neither symptom-level labels nor the stack explains it. A symptom-permissive output contract closes the breadth component of the deficit while the ordering component persists. Physician safety ratings are inconclusive by design.

*Implications of all the available evidence:* Gains measured on curated benchmarks should not be extrapolated to first-contact clinical text. Where multi-call designs are used on uncurated input, plain independent samples with a moderating call avoid the role-list penalty. An output contract permitting undifferentiated symptom-level entries is free—it harmed no arm—and is a defensible default wherever reference labels may be working diagnoses. Strategy selection by input, model, and endpoint remains a hypothesis requiring prospective evaluation, with independent emergency datasets and adequately powered safety studies.

## Introduction

Diagnostic error is a leading source of preventable harm [1,2], and large language models (LLMs) now perform competitively on benchmarks built for expert diagnosticians [3,4,5,6,7]. A frontier model has reached attending-level differential agreement on MGH Case Records [25], and direct assessments on standardized cases find differential construction the weakest reasoning stage [26]. A popular proposal simulates a team through specialist personas, conversational frameworks, or voting over independent generations [8,9,10,11], drawing on multidisciplinary practice [12]. The evidence is mixed: conversational multi-agent frameworks report gains [8,13], yet system-prompt personas show no stable improvement [14], debate can entrench error [15], and automatically assembled systems often underperform strong self-consistency baselines despite higher cost [17]. Single-call chain-of-thought prompting can also improve reasoning [16], while collaboration’s benefit shrinks with capability [22] and tool-using clinical agents show modest or inconsistent gains at substantially higher cost [24].

Two issues remain entangled. “Multiple perspectives” can mean personas voiced sequentially in one context or independent opinions combined afterwards, but these configurations also differ in calls, synthesis, and intermediate output structure. In addition, evaluation concentrates on curated, information-rich case reports rather than emergency presentations, where input is incomplete and the reference is the treating clinician’s working label. We compared single-call, sequential-persona, and moderated-team strategies across corpora differing in difficulty, provenance, and label structure. Our primary contrasts were MDT−A×1 at top-3 and top-5. We treated factorial and persona contrasts as secondary, emergency performance as judge-mediated working-label matching, and cross-corpus patterns as descriptive.

## Methods

### Study design and datasets

We compared three prompting strategies on datasets differing in difficulty, provenance, and label structure; all received the same case text and returned a ranked top-5 differential, scored against the reference diagnosis. The development corpus was 87 CPC cases (2013–2016) [5], split into 41 prompt-design and 46 held-out. Additional curated evaluation used 406 MedCaseReasoning cases [20], a 2025 benchmark with acknowledged contamination risk. The third dataset, ER-Reason [21], contains real emergency encounters from a PhysioNet credentialed repository used under its data use agreement: from 3,984 encounters, after deduplication, we drew a chief-complaint-stratified random sample of 364 (fixed seed). The input is the provider note truncated before “Medical Decision Making”; labels exist at symptom (n = 168) and disease (n = 196) levels. In revision, a replacement primary cohort of 400 disease-label encounters was drawn from the same repository under a protocol frozen before any generation: all original-cohort cases excluded, every candidate label audited, fixed-seed chief-complaint-stratified sampling, identical prompts and temperatures, one local serving stack, five seeds (self-frozen protocol; Statistical analysis); the original-cohort analysis is retained unchanged (Supplementary Note S39). Encounter text was processed on self-hosted infrastructure; adjudication exchanged only labels and candidate strings.

### Workup-informed input condition

To test whether additional information changed the emergency result, we appended each encounter’s objective results to its presentation (prompts, model, decoding, seeds, judge unchanged); filters, leakage checks, and the label-in-text analysis are in Supplementary Methods S-M8.

### Condensed-report, filler-arm, and replication-cohort experiments

To compare input forms on fixed content, we harvested full narrative case reports from PubMed Central and rewrote them into teaching vignettes under a condenser prompt forbidding disclosure of the reference diagnosis, with a zero-tolerance dual audit, an exploratory length-matched filler arm, and a protocol-frozen expansion to 392 replication vignettes; construction details and the selection-asymmetry disclosure are in the supplement (Supplementary Methods S-M6; Supplementary Note S35).

### Prompting strategies and control arms

The primary CPC, MedCaseReasoning, and emergency comparisons used qwen3.8-flash (176B, provider-documented, thinking disabled) and returned a ranked top-5 list. A×1 is a single zero-shot call at requested temperature 0; P reasons sequentially from five perspectives (attending physician, pathophysiology, imaging and laboratory, epidemiology and exposure, skeptic) in one shared context at 0·3; MDT issues the same five roles as independent, mutually blinded calls integrated by a moderator (six calls). P and MDT share roles, model, and prompt material but differ in call count, candidate independence, moderator synthesis, and intermediate output structure, so P–MDT is not a single-variable contrast. Control arms included A×1+CoT, five-sample self-consistency with Borda (A×5), A×5+Mod, MDT-Borda, P-split, moderator-anatomy ablations, a prompt-corrected Pfixed, a larger backbone, and the deepseek-v4.1-flash and gpt-5.1 families (Supplementary Tables S10–S12, S17); a post-hoc reasoning-mode arm (plan frozen before generation) added thinking-enabled single calls and mode-matched teams (Supplementary Note S38).

### Models, decoding, and compliance routing

Every ER-Reason arm was regenerated on self-hosted 4-bit infrastructure with prompts byte-identical to the curated runs and no case content leaving the machine; the single cloud pathway (the same-cohort deepseek replication) used audited zero-data-retention routing pinned to one provider. Because cloud endpoints need not honour requested temperature 0, the case, not the run, is the unit of inference (self-hosted runs are exact greedy). Deployment details, the temperature-matched reanalysis, and the output-contract sensitivity arm are in the supplement (Supplementary Methods S-M7; Supplementary Notes S36 and S39).

### LLM judge and validation

Scoring used one frozen LLM judge per candidate–reference pair [19], one cached verdict per pair; the primary judge (GLM-5.3-flash) and rules were frozen before outcome analyses (shared cache 253,005 pairs). Clinician-annotation samples assessed judge–clinician agreement, initially on a disjoint 100-pair batch rated by two blinded senior physicians and then on other corpora and the self-hosted rerun (Supplementary Tables S5, S5-2, S5-3). A second judge (deepseek-v4.1-flash) applied identical rules as a sensitivity analysis (disagreement 5·69% overall, 3·6–5·7% by stratum). Pairing does not eliminate arm-dependent misclassification (Supplementary Table S8).

### Blinded physician rating of the discordant pairs

Three non-author senior physicians (≥1 emergency medicine) independently rated all 34 emergency cases where the strategies’ top-1 verdicts disagreed plus 30 concordant controls under a frozen protocol with a pre-unblinding agreement gate (design and blinding: Supplementary Methods S-M9; Supplementary Note S37).

### Blinded physician rating on the replacement cohort

Because the primary emergency analysis moved to the replacement cohort, the safety assessment was repeated there under a protocol (v1.3) frozen before any rating material existed, with the primary question inverted to case-level rescue of the losing strategy’s list and the danger comparison retained as secondary (sampling frame and instruments: Supplementary Methods S-M10; Supplementary Note S39).

### Statistical analysis

The pre-specified primary endpoints were top-3 and top-5 recall; endpoints, primary judge, and comparison families were fixed before the comparisons were run; later control arms are labelled sensitivity analyses. The study was not publicly registered; pre-specification rests on dated, append-only cache milestones and self-frozen protocol versions. Tests use per-case hit rates across seeds (paired two-sided Wilcoxon; zero differences dropped) with 95% case-level cluster bootstrap intervals (10,000 resamples). Benjamini–Hochberg correction applies within two pre-specified families: the original 27 framework contrasts (12 of 14 raw p <0·05 survive) and the replacement cohort’s three protocol-frozen primary contrasts (adjusted p 0·0159, 0·0024, 0·0129). Battery arms report raw p, as pre-committed.

## Results

### Judge agreement with clinicians is corpus-dependent but stable across emergency cohorts

Before strategy comparison, the judge was assessed against two blinded clinicians on 100 development-untouched pairs (judge–clinician κ 0·814/0·878; inter-rater 0·812), with extension to other corpora (emergency symptom-level labels: inter-rater 0·612, judge–clinician 0·640/0·580; boundary-enriched rerun: 0·877, 0·580/0·540). The same design on the fresh cohort reproduced these values (0·560/0·540; 0·849), leaving measured judge reliability unchanged for the replacement analysis. Residual discordance was directional: clinicians endorsed 20 of 50 pairs the judge rejected but rejected only 3 of 50 it accepted, mostly concurrent or aetiologic candidates encompassing the reference; a full-corpus check found reference-containing false rejects running against the single call (8·8% vs 3·4%, p = 0·012), biasing the gap toward the null (Supplementary Tables S5, S5-2, S5-3; Note S39).

### MDT improves differential-list recall, not top-1 accuracy, on curated corpora

On 87 CPC cases, MDT exceeded A×1 at both primary endpoints (top-3 +5·5 points, 95% CI 1·1 to 10·3, p = 0·0079; top-5 +4·4, p = 0·015) (Table 1, Figures 1 and 2); no held-out 46-case contrast was significant (Supplementary Figure S2a). On 406 MedCaseReasoning cases MDT again improved top-3 (+3·0, 0·9 to 5·1, p = 0·0079) and top-5 (+3·9, p = 3·8 × 10□□) without a top-1 difference (p = 0·25); temperature-matched analyses preserved the advantage (Table 1).

**Figure 1.**
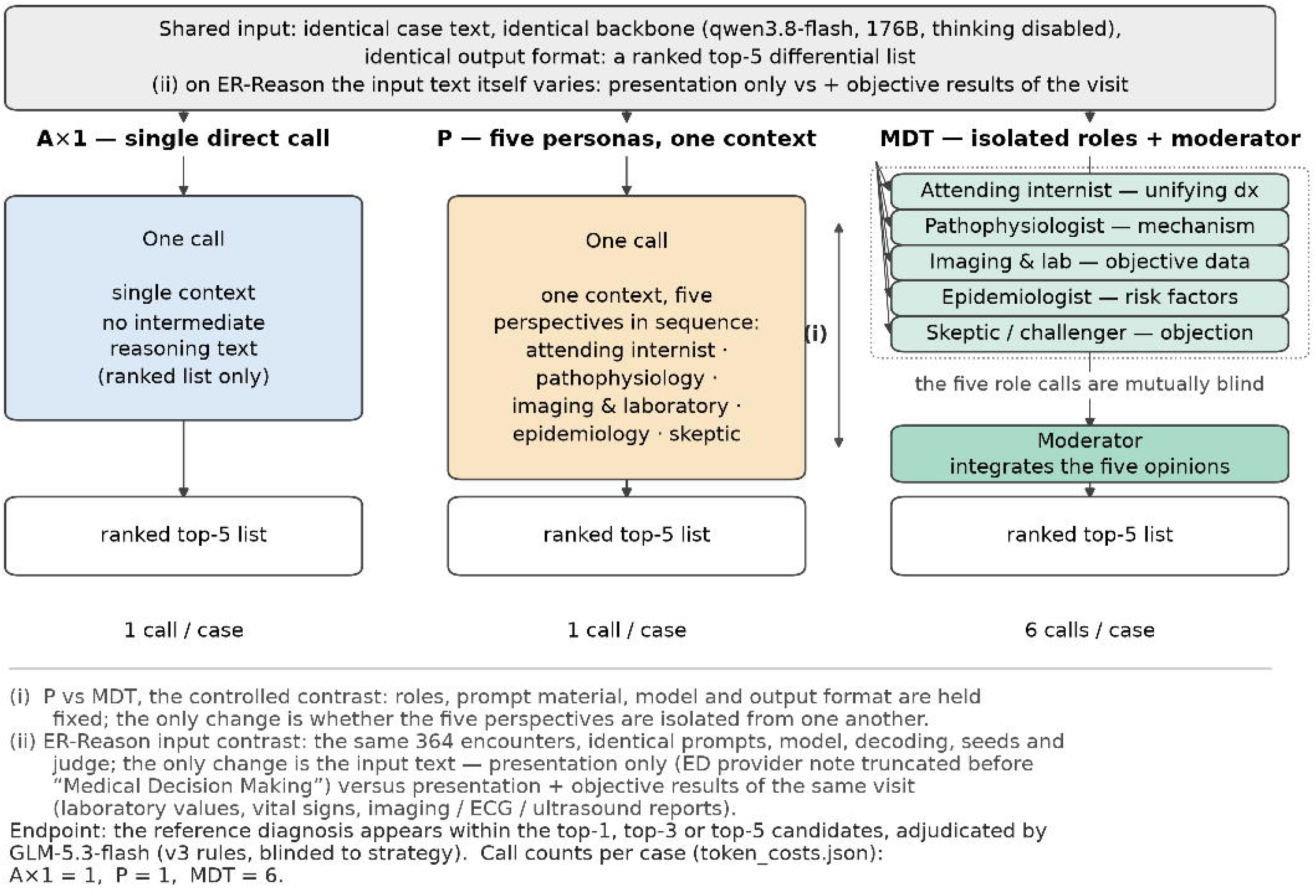
Study design: three prompting strategies and two input contrasts. A×1 (single call), P (five personas, one context), and MDT (five isolated roles plus moderator) returned ranked top-5 lists. P and MDT share roles and prompt material but differ in calls, candidate independence, moderator synthesis, and intermediate output structure. The emergency input contrast compares presentation only with presentation plus objective results. Judge: GLM-5.3-flash, frozen v3 rules, blinded to strategy.

**Figure 2.**
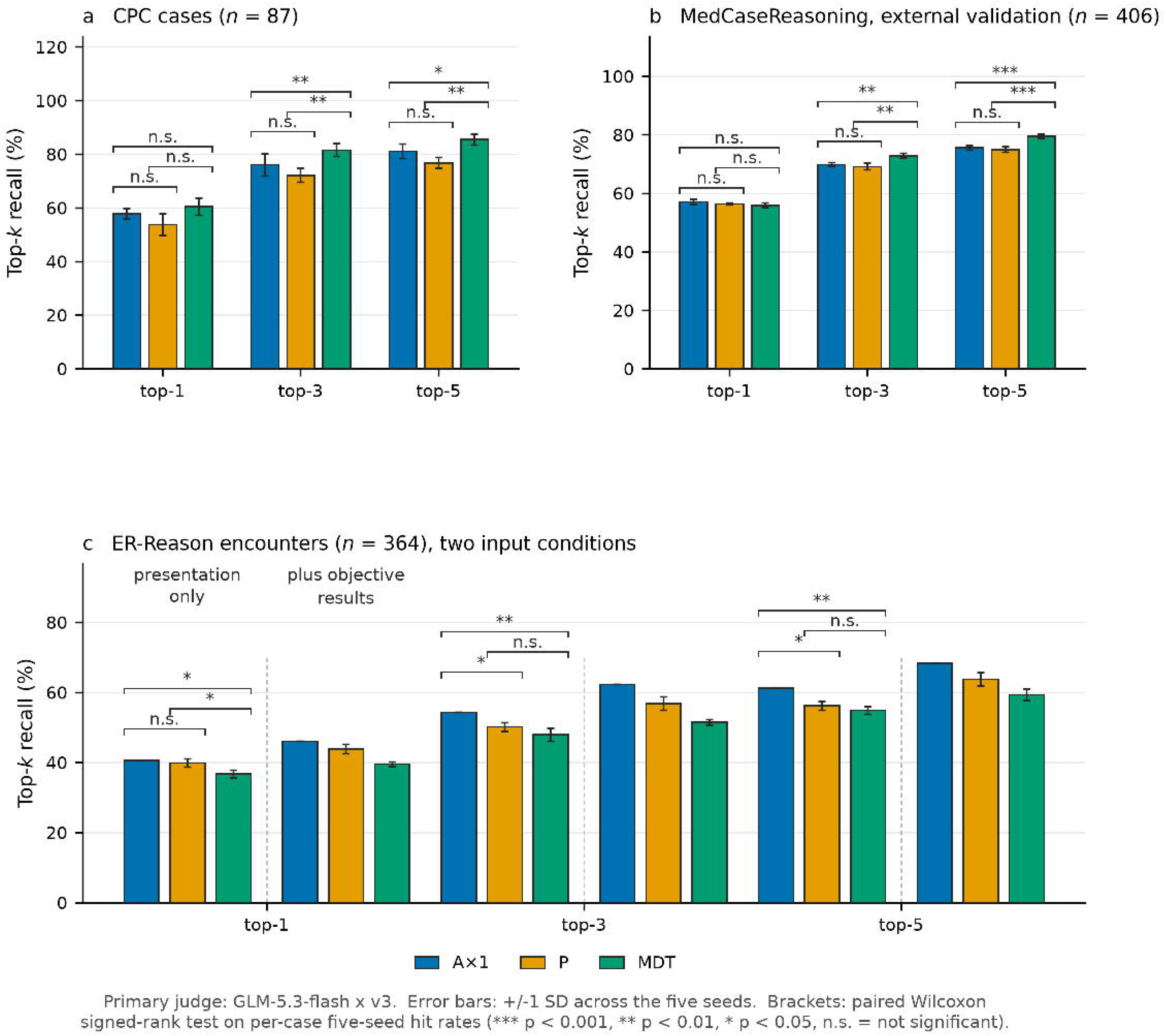
Top-k recall of the three strategies on (a) CPC, (b) MedCaseReasoning, and (c) ER-Reason under five seeds, with the ER-Reason input contrast. Bars give mean top-1/3/5 recall ±1 SD across five seeds; brackets give paired two-sided Wilcoxon tests on per-case five-seed hit rates; all p unadjusted (12 of the 14 primary-framework contrasts with p <0·05 survive Benjamini–Hochberg correction; Supplementary Table S4). In panel c the left triplet per endpoint is presentation-only input and the right triplet adds objective results. The top-3 strategy-by-input interaction disfavoured MDT (nominal p = 0·027); after exclusion of 20 label-in-text cases (n = 344), the MDT−A×1 interaction was −4·360 points (p = 0·0322).

**Table 1.**
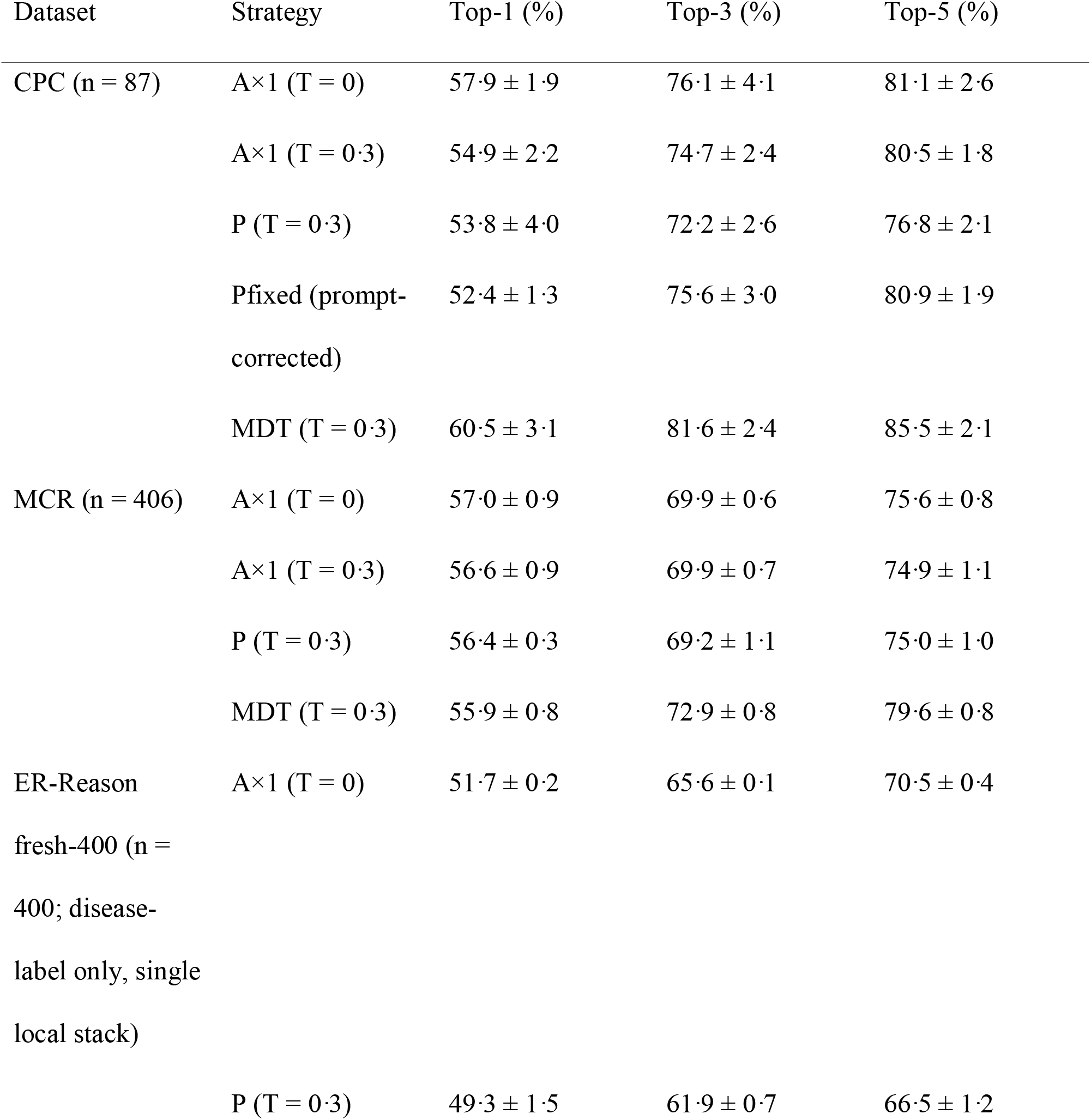
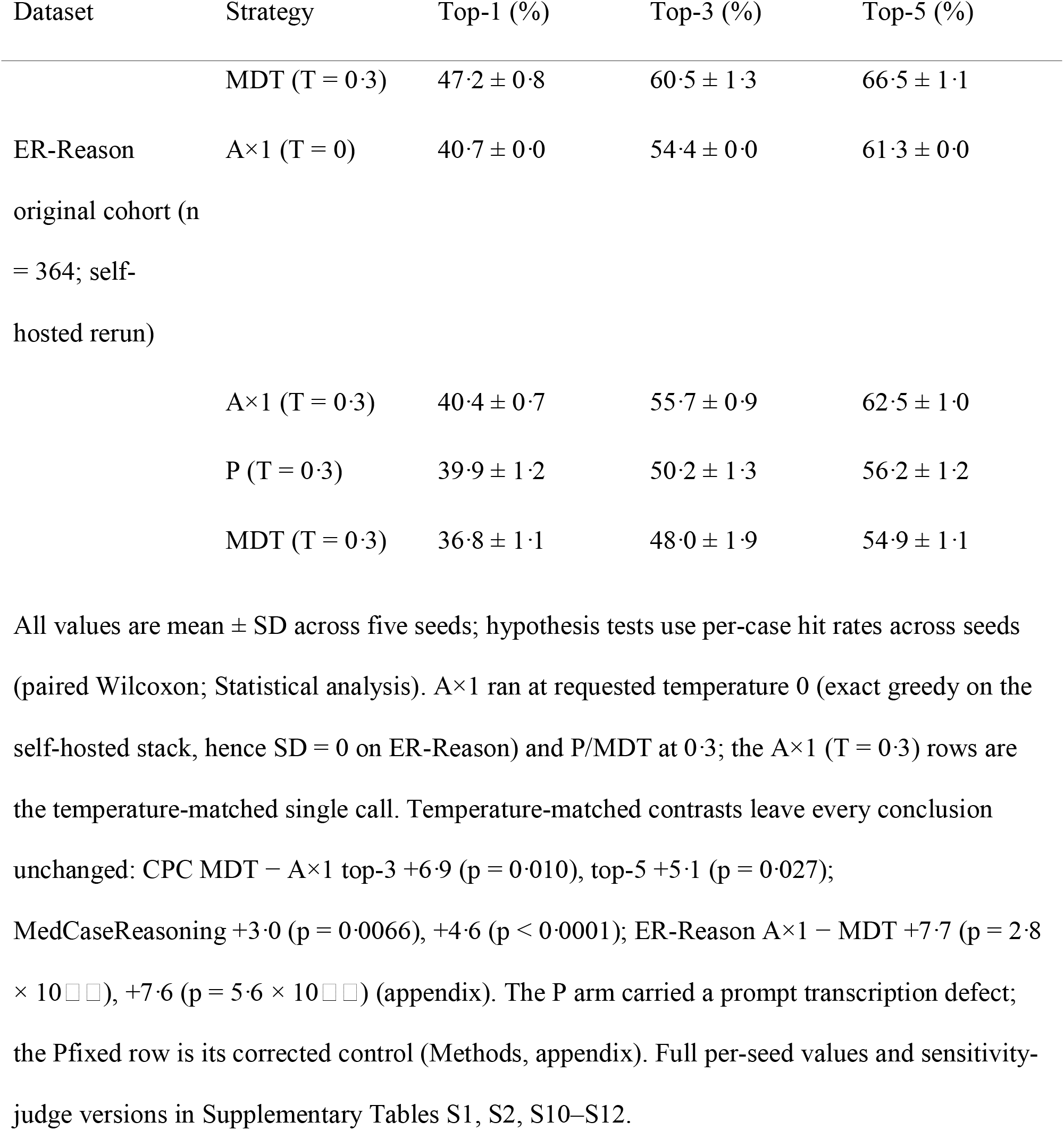
Diagnostic performance of the three strategies across datasets (primary judge, GLM-5.3-flash × v3). All values are mean ± SD across five seeds; hypothesis tests use per-case hit rates across seeds (paired Wilcoxon; Statistical analysis). A×1 ran at requested temperature 0 (exact greedy on the self-hosted stack, hence SD = 0 on ER-Reason) and P/MDT at 0·3; the A×1 (T = 0·3) rows are the temperature-matched single call. Temperature-matched contrasts leave every conclusion unchanged: CPC MDT − A×1 top-3 +6·9 (p = 0·010), top-5 +5·1 (p = 0·027); MedCaseReasoning +3·0 (p = 0·0066), +4·6 (p < 0·0001); ER-Reason A×1 − MDT +7·7 (p = 2·8 × 10□□), +7·6 (p = 5·6 × 10□□) (appendix). The P arm carried a prompt transcription defect; the Pfixed row is its corrected control (Methods, appendix). Full per-seed values and sensitivity-judge versions in Supplementary Tables S1, S2, S10–S12.

| Dataset | Strategy | Top-1 (%) | Top-3 (%) | Top-5 (%) |
|---|---|---|---|---|
| CPC (n = 87) | A×1 (T = 0) | 57·9 ± 1·9 | 76·1 ± 4·1 | 81·1 ± 2·6 |
|  | A×1 (T = 0·3) | 54·9 ± 2·2 | 74·7 ± 2·4 | 80·5 ± 1·8 |
|  | P (T = 0·3) | 53·8 ± 4·0 | 72·2 ± 2·6 | 76·8 ± 2·1 |
|  | Pfixed (prompt-corrected) | 52·4 ± 1·3 | 75·6 ± 3·0 | 80·9 ± 1·9 |
|  | MDT (T = 0·3) | 60·5 ± 3·1 | 81·6 ± 2·4 | 85·5 ± 2·1 |
| MCR (n = 406) | A×1 (T = 0) | 57·0 ± 0·9 | 69·9 ± 0·6 | 75·6 ± 0·8 |
|  | A×1 (T = 0·3) | 56·6 ± 0·9 | 69·9 ± 0·7 | 74·9 ± 1·1 |
|  | P (T = 0·3) | 56·4 ± 0·3 | 69·2 ± 1·1 | 75·0 ± 1·0 |
|  | MDT (T = 0·3) | 55·9 ± 0·8 | 72·9 ± 0·8 | 79·6 ± 0·8 |
| ER-Reason fresh-400 (n = 400; disease-label only, single local stack) | A×1 (T = 0) | 51·7 ± 0·2 | 65·6 ± 0·1 | 70·5 ± 0·4 |
|  | P (T = 0·3) | 49·3 ± 1·5 | 61·9 ± 0·7 | 66·5 ± 1·2 |
| Dataset | Strategy | Top-1 (%) | Top-3 (%) | Top-5 (%) |
|  | MDT (T = 0·3) | 47·2 ± 0·8 | 60·5 ± 1·3 | 66·5 ± 1·1 |
| ER-Reason original cohort (n = 364; self-hosted rerun) | A×1 (T = 0) | 40·7 ± 0·0 | 54·4 ± 0·0 | 61·3 ± 0·0 |
|  | A×1 (T = 0·3) | 40·4 ± 0·7 | 55·7 ± 0·9 | 62·5 ± 1·0 |
|  | P (T = 0·3) | 39·9 ± 1·2 | 50·2 ± 1·3 | 56·2 ± 1·2 |
|  | MDT (T = 0·3) | 36·8 ± 1·1 | 48·0 ± 1·9 | 54·9 ± 1·1 |

All values are mean ± SD across five seeds; hypothesis tests use per-case hit rates across seeds (paired Wilcoxon; Statistical analysis). A×1 ran at requested temperature 0 (exact greedy on the self-hosted stack, hence SD = 0 on ER-Reason) and P/MDT at 0·3; the A×1 (T = 0·3) rows are the temperature-matched single call. Temperature-matched contrasts leave every conclusion unchanged: CPC MDT − A×1 top-3 +6·9 (p = 0·010), top-5 +5·1 (p = 0·027); MedCaseReasoning +3·0 (p = 0·0066), +4·6 (p < 0·0001); ER-Reason A×1 − MDT +7·7 (p = 2·8 × 10[illegible]), +7·6 (p = 5·6 × 10[illegible]) (appendix). The P arm carried a prompt transcription defect; the Pfixed row is its corrected control (Methods, appendix). Full per-seed values and sensitivity-judge versions in Supplementary Tables S1, S2, S10–S12.

The original P arm scored below A×1 on CPC at every endpoint without significance; against Pfixed, MDT’s CPC advantages were likewise non-significant (top-3 6·0, p = 0·0599; top-5 4·6, p = 0·0676), on MedCaseReasoning borderline at top-3 (+2·4, p = 0·0501) and significant at top-5 (+3·6, p = 0·0007).

### Independent generation plus moderated synthesis, not personas, carries the curated gain

In the completed factorial (Figure 3a), mechanical aggregation never helped: Borda over five single-call samples scored 9·5 points below A×1 on MedCaseReasoning top-1, and re-aggregating team lists did not help either. The same moderator over plain independent samples (A×5+Mod) recovered the advantage and matched the role-based team; A×1+CoT changed nothing (all p ≥0·61) and A×5 did not reproduce MDT (all p ≤0·059). With the moderator-anatomy controls, the pattern is consistent with independent generation plus moderated synthesis. On emergency text, plain samples plus a moderator exceeded MDT by 6·8 points at top-3 and 7·0 at top-5 without differing from A×1 (appendix).

**Figure 3.**
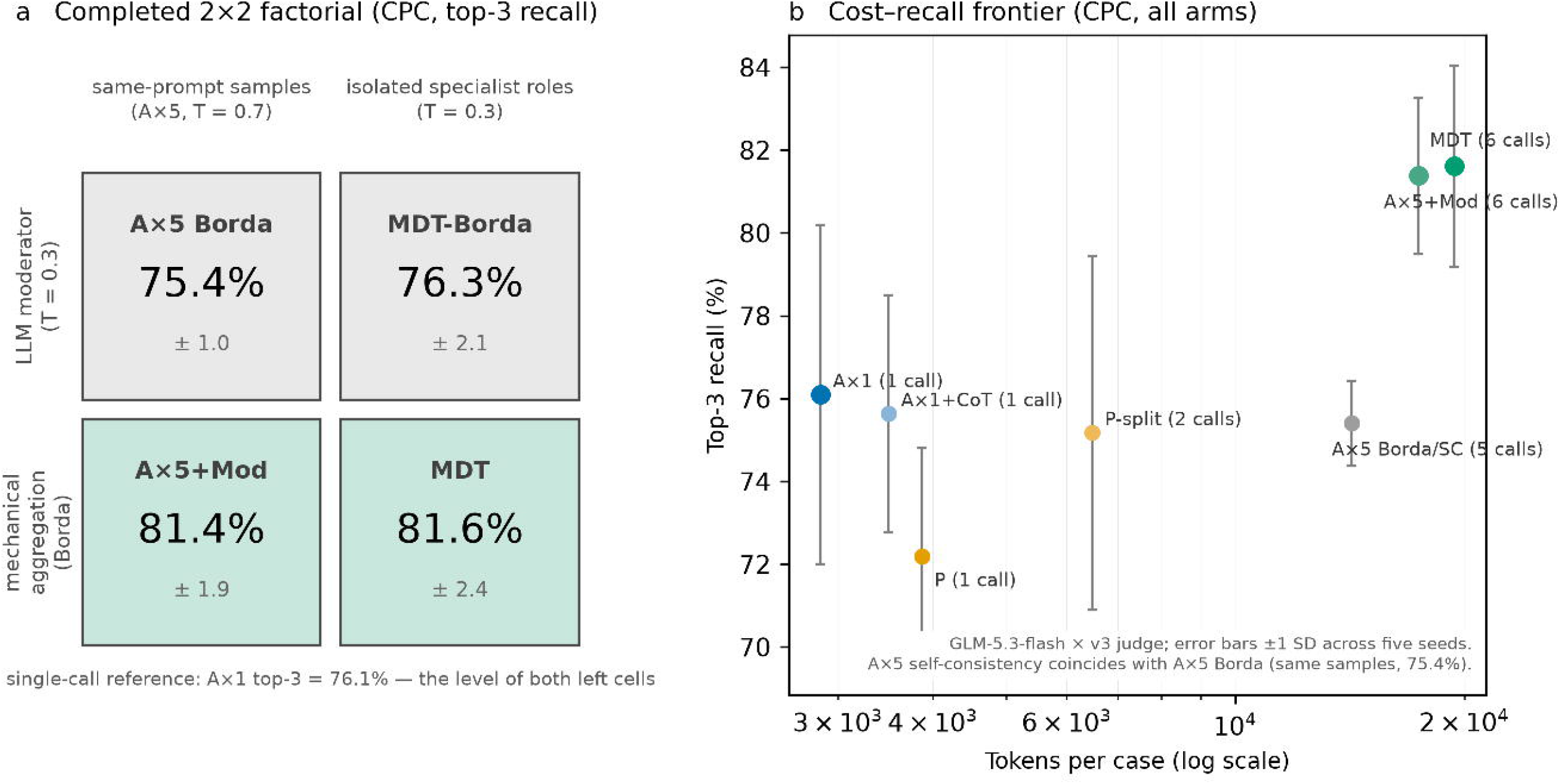
The completed factorial and the cost–recall frontier (CPC). (a) Top-3 recall of four factorial cells: candidate lists from same-prompt sampling (A×5) or five isolated specialist roles, integrated either by mechanical Borda aggregation or by the LLM moderator. Both moderator cells exceed both mechanical cells and are mutually indistinguishable, a pattern consistent with independent generation plus moderated synthesis rather than a requirement for specialist framing. (b) Top-3 recall versus tokens per case (log scale): the frontier runs A×1 → A×5+Mod → MDT; the +5·3-point gain of A×5+Mod over A×1 costs about six times A×1’s tokens. Judge: GLM-5.3-flash × v3; error bars ±1 SD across five seeds.

### On emergency presentations the effect reverses—and survives three stress tests

On the protocol-frozen 400-case emergency cohort whose reference labels are all disease-level diagnoses (Methods), A×1 outperformed MDT at every endpoint: top-1 51·7% versus 47·2% (−4·5 points, 95% CI −7·8 to −1·3, p = 0·016), top-3 −5·2 (−8·0 to −2·4, p = 0·0008), top-5 −4·0 (−6·7 to −1·5, p = 0·0086); majority-vote McNemar concordant, all three surviving in-family correction (adjusted p 0·0159, 0·0024, 0·0129). P also trailed A×1 on recall (top-3 −3·8, p = 0·047; top-5 −3·9, p = 0·021) and MDT added nothing over P, reproducing A×1 > P ≥ MDT at 7·3 times the single call’s token cost. On 50 bridged original-cohort cases the MDT−A×1 estimate shifted within noise (top-3 +2·0, top-5 +6·0, intervals spanning zero; both stacks negative), so the reversal is not a stack artefact and does not rest on symptom-level labels (Supplementary Note S39).

The original-cohort analysis (364 encounters, two stacks, 46% symptom-level labels) is retained in full: A×1 beat MDT at every endpoint (top-1 −3·9, p = 0·021; top-3 −6·4, p = 0·0022; top-5 −6·4, p = 0·0039) and in temperature-matched and judge-independent word-coverage checks (37·3% versus 33·0% P, 30·8% MDT; Supplementary Figure S2b)—lexical coverage being no clinical correctness measure (Supplementary Table S6; Figure 2c; Note S39).

A second model family reproduced the reversal on both cohorts: deepseek-v4.1-flash on the same 364 cases (top-3 −7·5, p = 1·8 × 10□□; same-cohort replication, not external clinical validation; Supplementary Table S11) and, on the fresh cohort under one local stack, the same ordering independently (−6·2/−7·1/−6·5 at top-1/3/5, p ≤ 0·0003); a fixed-50-case family bridge shifted the MDT−A×1 gap by 0·0 points at every endpoint between cloud and local servings (Note S39).

Adding objective results did not close the gap: in the original cohort they raised every strategy (+2·8 to +8·0 at top-1/3/5) but the top-3 strategy-by-input interaction disfavoured MDT (p = 0·027; Supplementary Table S6, Figure 2c), persisting after exclusion of 20 label-in-text cases (−4·360 points, 95% CI −8·256 to −0·464, p = 0·0322). On the fresh cohort the workup gain replicated for every strategy (+5·9 to +7·8, p ≤ 0·0002) and the reversal persisted (top-3 −4·9, p = 0·0031), but the interaction was null at every endpoint: the original cohort’s negative interaction did not replicate (Note S39).

### Input genre tracks the sign of the effect (a descriptive map)

Across corpora, MDT−A×1 top-3 point estimates ran +5·5 (CPC), +3·0 (MedCaseReasoning), +2·831 (condensed PMC vignettes, p = 0·0282), +1·108 (full PMC reports, p = 0·2134), and −6·4 (emergency notes) (Figure 4a). On the same 325 reports the paired condensed-minus-full interaction was +1·723 points (p = 0·2393) and the exploratory single-seed filler arm +3·1 (p = 0·096). The pooled 392-vignette replication cohort gave top-3 +3·3 (95% CI 1·5 to 5·1, p = 0·0013).

**Figure 4.**
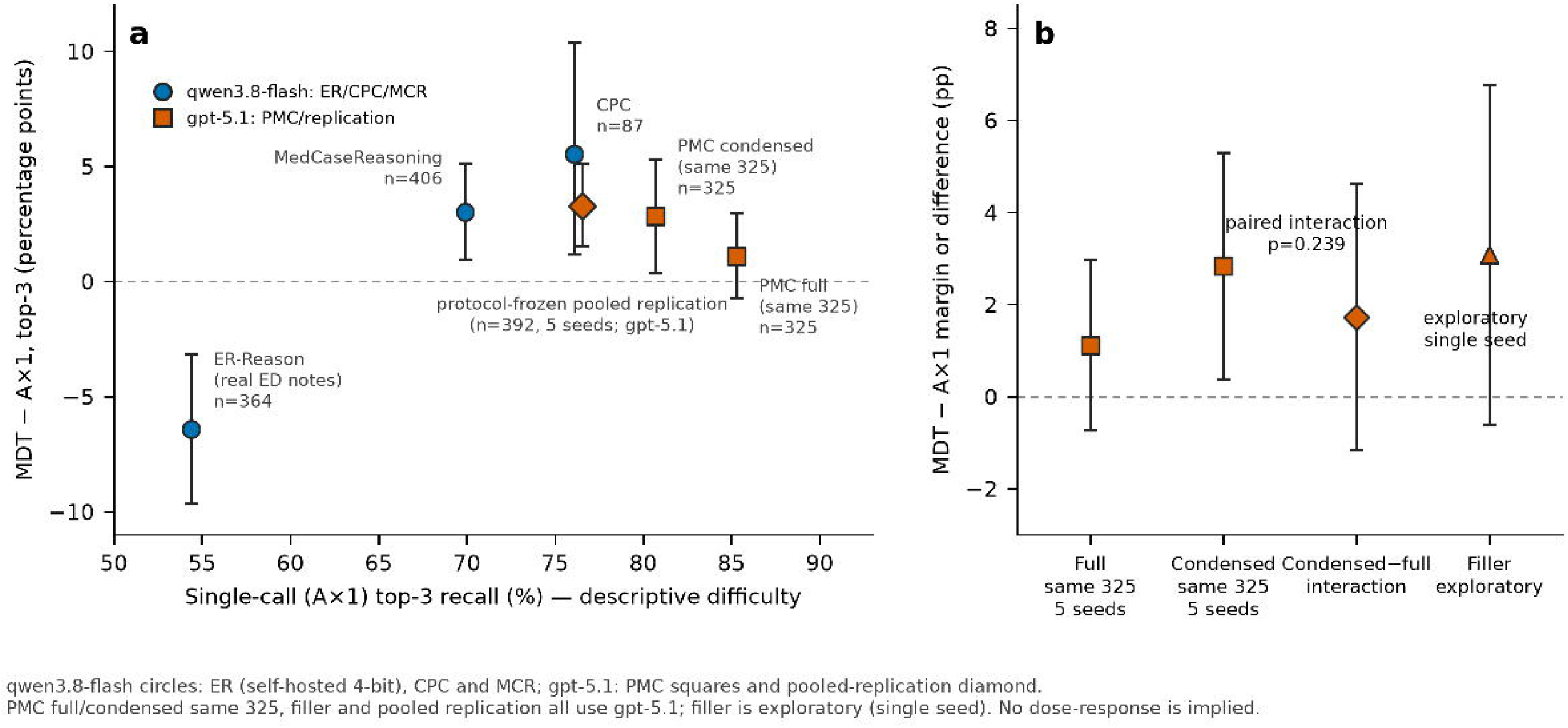
Descriptive corpus contrasts and the same-case format comparison. Both panels show case-level MDT−A×1 top-3 margins with 95% percentile bootstrap intervals and paired two-sided Wilcoxon tests on per-case five-seed hit rates. (a) Unconnected descriptive points ordered by A×1 top-3 rate: ER-Reason (−6·4 points, p = 0·0022), MedCaseReasoning (+3·0, p = 0·0079), CPC (+5·5, p = 0·0079), same-325 condensed PMC (+2·831, 95% CI 0·369 to 5·292, p = 0·0282), and same-325 full PMC (+1·108, −0·738 to 2·954, p = 0·2134). Blue circles denote qwen3.8-flash only for ER-Reason (self-hosted 4-bit), CPC, and MedCaseReasoning; orange squares denote gpt-5.1 PMC full and condensed results. The orange diamond is the gpt-5.1 pooled 392-vignette replication cohort (+3·3, 95% CI 1·5 to 5·1, p = 0·0013). Provenance, model, deployment, labels, and contamination differ across corpora; these points do not establish a dose–response. (b) On the same 325 reports, the paired condensed-minus-full interaction was +1·723 points (95% CI −1·169 to 4·615, p = 0·2393). The length-matched filler arm is exploratory and single-seed (orange triangle; +3·1, p = 0·096). Judge: GLM-5.3-flash × v3; statistics in Supplementary Note S35.

These estimates are consistent with heterogeneity across input formats but do not identify curation as its driver, exclude length effects, or support a dose–response curve: model, deployment, labels, provenance, and contamination all differ across corpora (Figure 4; Supplementary Note S35).

### Output-contract fit tracks the magnitude: a safe one-sentence partial fix

Because the strict contract demands disease-level entries while many emergency labels are working diagnoses, we tested a symptom-permissive contract—one appended sentence permitting undifferentiated entries over unsupported disease guesses (Methods; plan frozen before generation). On the original cohort it closed the breadth end of the symptom-stratum deficit (top-5 −9·6 points, p = 0·0036, to −3·2, interval spanning zero) while the ordering end persisted. On the fresh cohort, where every label is a disease diagnosis, the same contract narrowed the gap at every endpoint (point estimates shrinking 52–75%, all intervals spanning zero; gap shift +3·0 at top-5, p = 0·06): part of the reversal is an artefact of forced disease guessing under the strict contract on which the pre-specified primary result rests. The clause was safe and selective: on the original cohort it added +6·4 top-5 points to MDT and +5·1 to P (both p <0·0001) with A×1 unchanged; on the fresh cohort it left A×1 unchanged (+1·4/+2·5/+1·6 at top-1/3/5, all p ≥ 0·11) but lifted MDT by +4·0/+5·2/+4·6 (p = 0·0031, 1·8 × 10□□, and 2·7 × 10□□ at top-1/3/5)—the signature of forced differentiation penalising the team’s more dispersed candidate lists, which a generation-side clause could not produce under a judge-side artefact (Supplementary Notes S36, S39).

### Model family, reasoning mode, and cost

On CPC with deepseek-v4.1-flash, MDT had a non-significant top-1 advantage over A×1 (+4·8, p = 0·067) and exceeded P at top-3 and top-5 (p = 0·017, 0·023); on gpt-5.1, P exceeded A×1 at top-1 (+7·1, p = 0·0009) without a top-5 difference, while MDT exceeded A×1 at top-3 (+10·9, p = 0·0003); on the pooled replication cohort P again led at top-1 (+4·5, p <0·0001) and MDT improved top-3 (+3·3, p = 0·0013) (Supplementary Figure S3; Supplementary Tables S11, S17). These model-dependent P results argue against a general persona penalty. Non-thinking MDT consumed 6·8–8·4 times A×1’s tokens (Table 2).

**Table 2.** Calls and tokens per case. Token logs do not separate input from output; MDT values include all five role calls and the moderator. Control-arm token accounting is in Supplementary Tables S10–S12.

| Dataset | Strategy | Calls per case | Tokens per case (mean ± SD) |
|---|---|---|---|
| CPC (n = 87) | A×1 | 1 | 2,846 ± 1,268 |
|  | P | 1 | 3,872 ± 1,302 |
|  | MDT | 6 | 19,422 ± 7,686 |
| MCR (n = 406) | A×1 | 1 | 850 ± 123 |
|  | P | 1 | 1,797 ± 198 |
|  | MDT | 6 | 7,137 ± 864 |
| ER-Reason (n = 364, self-hosted rerun) | A×1 | 1 | 1,678 ± 472 |
|  | P | 1 | 2,673 ± 492 |
|  | MDT | 6 | 12,297 ± 2,910 |

A thinking-enabled single call closed the CPC non-thinking-team advantage and beat the non-thinking team on MedCaseReasoning, and under fully symmetric thinking the CPC team increments closed while the MedCaseReasoning top-3/5 advantages persisted; the full six-call thinking team used 73,866 tokens per case (4·92× the thinking single call). These mode-matched, not token-budget-matched, contrasts are in Supplementary Note S38.

### Physician safety ratings are inconclusive

Three physicians rated seed-1 lists for all 34 top-1-discordant original-cohort cases. S1 danger (mean AC1 0·185), dangerous-omission (0·423/0·458), and D5 preference (0·196) missed the 0·6 agreement gate; only misleading-inclusion ratings met it (0·834/0·726). The safety comparison is therefore inconclusive. Descriptively, 23 of 34 majority verdicts were ties (non-tied 6:5; MDT-more-dangerous rate 0·545, 95% CI 0·234 to 0·833); omission flags favoured A×1 (26·5% versus 14·7%), misleading-inclusion flags MDT (0% versus 14·7%), but power covered only preferences beyond ∼72% (Supplementary Figure S2d; Supplementary Note S37).

On the replacement cohort the redesigned rating passed the gate for every family except D5 preference (mean AC1 0·836 danger, 0·819/0·992 omission/inclusion flags, 0·822 rescue; D5 0·467, descriptive), with perfect intra-rater agreement (14/14 embedded repeats each). The primary question—did the losing list contain an acceptable entry encompassing the reference?—was answered yes for 29 of 35 team-lost cases (82·9%) and 18 of 18 single-call-lost cases (100%); the −17·1-point difference (95% Newcombe CI −32·7 to +2·6) leaves the forced-differentiation mechanism neither validated nor refuted, in the direction opposite to the hypothesis (baseline rescue on concordant misses 6/15, 40%). The secondary danger comparison was null (MDT more dangerous in 3 of 11 non-tied units, p = 0·23; omission flags 8:3, p = 0·23; misleading inclusion 0:0; no side preference in controls) (Supplementary Note S39).

## Interpretation

Whether a multi-agent team helped, added nothing, or underperformed depended on configuration, not architecture. On curated corpora the moderated team improved the differential’s breadth without robustly improving its head; the factorial attributed the gain to independent generation plus moderated synthesis. On uncurated emergency presentations the sign reversed—persisting with all disease-level labels, in a second model family, and across serving stacks—and multi-call inference is not the cause: five plain samples under the same moderator matched the single call while the role-based team sat 6·8 points below it at top-3.

Two moderators organise this heterogeneity. Input genre: the margin ran from positive on curated vignettes through null on self-disclosing narratives to negative on uncurated emergency text, the within-case contrast pointing the same way. Output-contract fit is the actionable moderator: a one-sentence clause permitting undifferentiated entries closed the breadth component of the emergency deficit (halving point estimates even where every label was a disease diagnosis), the ordering component persisting. Two strictnesses must be distinguished: scoring strictness fell on both arms and explains the gap in neither direction; contract strictness was not arm-neutral—the clause preferentially restored the team’s recall (+4 to +5 versus +1 to +2 points), the signature of a dispersed candidate list forced to guess beyond the evidence. The reversal is therefore partly an artefact of forced differentiation and partly a genuine ordering deficit; the frozen-protocol primary result rests on the strict contract shared with the other corpora. The safety evidence, inconclusive on both cohorts, establishes neither clinical equivalence nor that the reversal is merely artefactual.

These results reconcile mixed literature: positive studies generated contributions separately and aggregated them [8,13]; negative studies voiced personas in one context [14] or debated without feedback [15]; reports of models beating physicians on emergency cases used complete records and a comprehensiveness score that rewarded inclusion without penalising irrelevance [27], unlike our first-contact working-label design. Collaboration benefits vary with capability and reasoning configuration [22,23]; agent count and persona framing show no uniform effect across clinical tasks [17,18,24].

Curated-benchmark gains should not be extrapolated to first-contact clinical input—the one setting whose labels no model could have memorised. On uncurated input, plain independent samples with a moderating call are the defensible multi-call form, and a contract permitting undifferentiated entries is a defensible default wherever labels may be working diagnoses.

Strategy selection by input, model, endpoint, and reasoning mode remains a routing hypothesis for prospective evaluation; and because the reversal is label-level, judge-mediated, and clinically unadjudicated, these results argue for caution about deployment claims, not clinical harm.

Several limitations bound inference. The primary emergency input stops before objective results, and the workup condition added objective content only; findings do not extend to complete-chart reasoning. Both emergency cohorts come from one repository, use working diagnoses, and were scored by an imperfect judge—the replacement cohort removes label granularity but shares repository, working-label construct, and judge, whose clinician-validated κ on the new candidate distribution (0·54–0·56 against clinician–clinician 0·85) matches the original, leaving the reversal resting partly on a noisy instrument (Note S39). The second-family result is same-cases model replication, not external validation; the same-rule second judge is a sensitivity analysis, not independent clinical validation; and the annotation samples support no unbiased arm-specific false-accept/false-reject estimates. Both physician ratings were inconclusive: the original-cohort danger, omission, and preference families missed the agreement gate, and the replacement-cohort rescue contrast was unadjudicable (95% CI −32·7 to +2·6 points; power limited to ∼35–40-point differences; D5 descriptive; 8 of 83 Part A units with a pre-declared seed-1 display deviation). In the original cohort the deficit was directionally present in both label strata but separable only in the symptom stratum (top-3 and top-5 q = 0·049 and 0·022), and its magnitude was serving-stack-sensitive (top-3 −9·0 cloud to −6·4 self-hosted, 4-bit quantisation a candidate explanation), though bridged cases bounded the stack contribution (50 cases, one seed; Note S39). The output-contract arm was added after the primary analyses under a pre-generation frozen plan, and its fresh-cohort intervals all span zero. Clinician judge assessment did not cover the closed-model arms; part of the condensed-corpus adjudication used an alternative endpoint (94·99% agreement after equivalence checking). Curated-corpus contamination and strategy-by-memorisation interactions cannot be excluded. Cross-corpus contrasts confound model, deployment, labels, provenance, and contamination. Analyses were not externally registered, and thinking arms were post hoc, excluded the emergency corpus, and yielded corpus-specific results.

## Data Availability

The three datasets used differ in availability. The 87 CPC case texts are copyrighted by the publisher and cannot be redistributed; the released repository provides case identifiers with pointers to the source journal (Case Records of the Massachusetts General Hospital) and to the curating benchmark. The 406 MedCaseReasoning cases are publicly available under a CC-BY license. The 364 ER-Reason encounters are available from PhysioNet to users who complete credentialing and sign its data use agreement; per that agreement, per-case ER-Reason outputs and ER-Reason entries of the judgment caches are excluded from the public archive, and case identifiers are one-way hashed. No new primary patient data were generated in this study.

## Data sharing

The full analysis pipeline (prompts, per-arm scripts, run files, and both judge caches) is openly released (GitHub; Zenodo DOI at publication). CPC case text is copyrighted and is released as identifiers with pointers to the source; MedCaseReasoning is publicly available under CC-BY; the PMC replication corpora are released as article identifiers, per-stage curation and audit metadata, and the condensed vignette texts that cleared the dual audit, under the redistribution terms of the source articles’ open-access licenses. Per-case ER-Reason outputs and the ER-Reason-derived entries of the judge caches are excluded from the public archive under ER-Reason’s data use agreement, with case identifiers one-way hashed; the zero-retention routing audit log of the deepseek replication (provider per request) is retained by the authors and available on reasonable request. No individual-level data from the emergency corpus can be shared by the authors beyond what PhysioNet credentialled access already provides.

## Ethics

This study is a secondary analysis of existing, de-identified data with no human-subjects recruitment and no access to identifiable information, and the institutional ethics committee reviewed the protocol and granted an exemption from institutional review board review (No. I-26ZM0145). It was conducted in accordance with the ethical standards of the 1964 Declaration of Helsinki and its later amendments. ER-Reason consists of clinical notes de-identified by the UCSF Information Commons under HIPAA Safe Harbor and Expert Determination methods and was used under the PhysioNet data use agreement. Consistent with PhysioNet’s guidance on credentialed data and large language models, all automated processing of these de-identified records met the guidance’s conditions for acceptable automated processing: the emergency-corpus arms ran on self-hosted infrastructure with no transmission of case content off the machine, and the one replication that sent case text to a cloud provider used a zero-data-retention endpoint (no retention, no training use, no human review of case content), with the serving provider logged and audited on every request. An amendment documenting this processing pathway was approved by the data provider’s credentialing committee before the replication ran. The physician rating component involved three practising clinicians assessing anonymised diagnosis lists derived from the same de-identified corpora on a local server, with no patient involvement and no case content leaving the rating environment; it is covered by the same exemption (No. I-26ZM0145).

## Code availability

All analysis code, prompt templates, pipeline scripts, and the judgment caches are openly released (https://github.com/yepsun/multi-agent-diagnostic-reasoning; Zenodo concept DOI 10.5281/zenodo.22935059, resolving to the latest version; the v1.1.1 snapshot is 10.5281/zenodo.23995615). The caches contain every adjudication used for CPC and MedCaseReasoning; entries derived from ER-Reason are excluded and case identifiers one-way hashed, in line with the ER-Reason data use agreement.

## Declarations

### Contributors

Conceptualisation and supervision: XS. Methodology, software, formal analysis, and investigation: JF and YJ (equal contribution). Data curation and validation: YL, LX, WP, HX, and JW. Writing — original draft: JF and YJ. Writing — review and editing: all authors. XS is the guarantor and accepts full responsibility for the work and the conduct of the study.

### Declaration of interests

We declare no competing interests.

### Declaration of AI use

Large language models were used to assist with drafting and language editing; the authors reviewed, verified, and take full responsibility for all content, including every reported number and citation.

### Funding

This research received no specific grant from any funding agency in the public, commercial, or not-for-profit sectors.

## Notes

### Competing Interest Statement

The authors have declared no competing interest.

### Author Declarations

This study is a secondary analysis of existing, de-identified data with no human-subjects recruitment and no access to identifiable information; the institutional ethics committee reviewed the protocol and granted an exemption from institutional review board review (No. I-26ZM0145). The CPC cases are published case reports; MedCaseReasoning is derived from open-access case reports in PubMed Central. ER-Reason consists of clinical notes de-identified by the UCSF Information Commons under HIPAA Safe Harbor and Expert Determination methods and was used under the PhysioNet data use agreement. Consistent with PhysioNet's guidance on credentialed data and large language models, all automated processing of these de-identified records met the guidance's conditions for acceptable automated processing: the emergency-corpus arms ran on self-hosted infrastructure with no transmission of case content off the machine, and the one replication that sent case text to a cloud provider used a zero-data-retention endpoint, with the serving provider logged and audited on every request; an amendment documenting this processing pathway was approved by the data provider's credentialing committee before that replication ran. The physician rating component involved three practising clinicians assessing anonymised diagnosis lists derived from the same de-identified corpora on a local server, with no patient involvement and no case content leaving the rating environment; it is covered by the same exemption (No. I-26ZM0145).

### Summary of Updates

In this revision the emergency-department evidence base is rebuilt under a protocol frozen before any new generation, and the paper is reframed as a two-cohort, two-model-family evaluation. First, the primary emergency analysis moves to a new fully held-out replacement cohort: 400 encounters whose reference labels are all disease-level diagnoses, drawn from the same repository with every original-cohort case excluded, after a per-label audit of all candidate labels (symptom, incidental-finding, status, and injury-mechanism forms removed), generated entirely on one local serving stack across five seeds. The reversal reported in the original posting persists on this stricter cohort at every endpoint (team minus single call: top-3 -5.2 points, 95% CI -8.0 to -2.4, p = 0.0008), so it does not rest on symptom-level labels. Second, a full pre-registered battery accompanies the new cohort: a workup arm (adding each visit's objective results) shows the reversal is input-invariant; a symptom-permissive output-contract arm narrows the deficit's point estimates by 52-75% with intervals spanning zero, locating part of the deficit in forced disease-level guessing; a second model family (DeepSeek) under the same local stack reproduces the ordering independently; and fixed-case bridges show the reversal's magnitude is serving-stack-invariant in both families even where absolute accuracy moves. Third, the LLM judge was revalidated against two blinded clinicians on the new cohort's candidate distribution (agreement essentially identical to the original validation), and the physician rating was redesigned under a pre-frozen protocol with the primary question inverted to case-level rescue of the losing list; its agreement gate largely passed, and the rescue contrast was unadjudicable (95% CI -32.7 to +2.6 points), leaving the safety question inconclusive. The original 364-encounter analysis is retained in full in the main text. The reasoning-mode sensitivity arm of the previous revision is unchanged. Title, abstract, and interpretation are updated to the two-cohort framing; multiplicity correction is applied within two pre-specified families and all replacement-cohort primary contrasts survive. No previously reported result was altered; the public data archive was re-released (v1.1.1) with stricter screening of judge-cache entries under the data use agreement.

## References

1. National Academy of Medicine. Improving Diagnosis in Health Care. Washington, DC: National Academies Press; 2015. doi:10.17226/21794.

2. Newman-Toker DE, et al. Burden of serious harms from diagnostic error in the USA. BMJ Qual Saf. 2024;33:109–120. doi:10.1136/bmjqs-2021-014130.

3. Singhal K, et al. Large language models encode clinical knowledge. Nature. 2023;620:172–180. doi:10.1038/s41586-023-06291-2.

4. Kanjee Z, Crowe B, Rodman A. Accuracy of a generative artificial intelligence model in a complex diagnostic challenge. JAMA. 2023;330:78–80. doi:10.1001/jama.2023.8288.

5. Buckley TA, et al. Teaching large language models to reason like expert diagnosticians. arXiv preprint arXiv:2509.12194. 2025.

6. McDuff D, et al. Towards accurate differential diagnosis with large language models. Nature. 2025;642:451–457. doi:10.1038/s41586-025-08869-4.

7. Tu T, et al. Towards conversational diagnostic artificial intelligence. Nature. 2025;642:442–450. doi:10.1038/s41586-025-08866-7.

8. Chen X, et al. Enhancing diagnostic capability with multi-agents conversational large language models. npj Digit Med. 2025;8:159. doi:10.1038/s41746-025-01550-0.

9. Wang J, et al. Mixture-of-agents enhances large language model capabilities. In: Proceedings of the International Conference on Learning Representations (ICLR). 2025.

10. Wang X, et al. Self-consistency improves chain of thought reasoning in language models. In: Proceedings of the International Conference on Learning Representations (ICLR). 2023.

11. Li J, et al. More agents is all you need. Trans Mach Learn Res. 2024.

12. Lamb BW, et al. Quality of care management decisions by multidisciplinary cancer teams: a systematic review. Ann Surg Oncol. 2011;18:2116–2125.

13. Tang X, et al. MedAgents: large language models as collaborators for zero-shot medical reasoning. In: Findings of the Association for Computational Linguistics: ACL 2024. 2024.

14. Zheng M, et al. When “a helpful assistant” is not really helpful: personas in system prompts do not improve performances of large language models. In: Findings of the Association for Computational Linguistics: EMNLP 2024. 2024. doi:10.18653/v1/2024.findings-emnlp.888.

15. Oh J, Jeong M, Ko J, Yun SY. From belief entrenchment to robust reasoning in LLM agents. Trans Assoc Comput Linguist. 2026;14:1286–1307. doi:10.1162/tacl.a.728.

16. Wei J, et al. Chain-of-thought prompting elicits reasoning in language models. In: Advances in Neural Information Processing Systems 35 (NeurIPS). 2022.

17. Jwalapuram J, et al. The Illusion of Multi-Agent Advantage. arXiv preprint arXiv:2606.13003. 2026.

18. Yuan GC, Zhang X, Kim SE, Rajpurkar P. Do mixed-vendor multi-agent LLMs improve clinical diagnosis? In: Proceedings of the 1st Workshop on Linguistic Analysis for Health (HeaLing). 2026. doi:10.18653/v1/2026.healing-1.1.

19. Zheng L, et al. Judging LLM-as-a-judge with MT-Bench and Chatbot Arena. In: Advances in Neural Information Processing Systems 36 (NeurIPS), Datasets and Benchmarks Track. 2023.

20. Wu K, et al. MedCaseReasoning: evaluating and learning diagnostic reasoning from clinical case reports. arXiv preprint arXiv:2505.11733. 2025.

21. Golchini N, Zack T, Bamman D, Molina M, Alaa A. ER-Reason: a benchmark dataset for LLM-based clinical reasoning in the emergency room. arXiv preprint arXiv:2505.22919; 2025. Dataset: PhysioNet; 2025. doi:10.13026/55s7-3c27.

22. Kim Y, et al. Capable language models can outgrow the benefits of collaboration. Nat Mach Intell. 2026;8:1157–1172. doi:10.1038/s42256-026-01268-y.

23. Tran D, Kiela D. Single-agent LLMs outperform multi-agent systems on multi-hop reasoning under equal thinking token budgets. arXiv preprint arXiv:2604.02460. 2026.

24. Liu Y, et al. Benchmarking large language model-based agent systems for clinical decision tasks. npj Digit Med. 2026;9:259.

25. Brodeur PG, et al. Performance of a large language model on the reasoning tasks of a physician. Science. 2026;392:524–527. doi:10.1126/science.adz4433.

26. Rao AS, et al. Large language model performance and clinical reasoning tasks. JAMA Netw Open. 2026;9(4):e264003. doi:10.1001/jamanetworkopen.2026.4003.

27. Wei J, et al. Performance evaluation of large language models in the diagnosis of emergency internal medicine diseases: a retrospective study. Front Public Health. 2026;14:1780425. doi:10.3389/fpubh.2026.1780425.

