## Supplementary Material: Methods, Tables S1-S17, Figures S1-S3, Notes S34-S39 for "Multi-agent LLM teams improve differential recall on curated cases but underperform on emergency working-label matching: a two-cohort, two-family evaluation"

**Contents**

- Supplementary Methods: full prompt text
- Case Presentation
- Diagnostic Perspectives
- Synthesis Task
- Output Requirement (final block)
- Supplementary Table S1. Per-seed accuracy of the three strategies (primary judge, GLM-5.3-flash × v3)
- Supplementary Table S2. Main results under the sensitivity judge (deepseek-v4.1-flash × v3 rules)
- Supplementary Table S3. Legacy pooled-seed McNemar statistics versus the primary case-level analysis
- Supplementary Table S4. Benjamini–Hochberg correction across the 27 tests of the primary analysis framework
- Supplementary Table S5. Judge validation: Cohen’s κ against two blinded clinician raters
- Supplementary Table S6. Workup-informed versus presentation-only input on ER-Reason (n = 364, primary judge, five seeds)
- Supplementary Table S7. Held-out (46) and development (41) subsets, primary case-level analysis
- Supplementary Table S8. Judging agreement between the two LLM judges, by dataset and by ER label granularity
- Supplementary Table S9. Workup-informed input excluding cases with no objective content (n = 330)
- Supplementary Note. ER-Reason encounter sampling and input construction
- Supplementary Note. Temperature-matched analyses
- Supplementary Table S10. Matched control arms on the CPC set (n = 87, five seeds, primary judge)
- Supplementary Table S11. Second model family (deepseek-v4.1-flash) on the CPC and ER-Reason sets (five seeds, primary judge)
- Supplementary Table S12. Backbone scale (176B versus 2400B) and the isolated team versus a single larger call
- Supplementary Note. Prompt transcription defect and the corrected-prompt control
- Supplementary Note. Data integrity repair
- Supplementary Table S15. GEE sensitivity analysis: the 27 primary tests re-fit as case-clustered observation-level models
- Supplementary Table S16. Majority-vote threshold sensitivity: the 27 primary tests at t = 2/3/4/5 of five seeds
- Supplementary Figure S1
- Supplementary Figure S2
- Supplementary Figure S3
- Supplementary Note S34. Judge-independent word coverage on CPC and MedCaseReasoning
- Supplementary Note S35. Input format and cross-corpus descriptive analyses
- Supplementary Note S36. The output-contract sensitivity arm: partitioning the emergency deficit by endpoint
- Supplementary Note S37. Blinded physician rating of the discordant emergency pairs
- Supplementary Note S38. Reasoning-mode sensitivity arm: thinking-enabled single call and fully symmetric thinking on CPC and MedCaseReasoning
- Supplementary Note S39. Protocol-frozen disease-label-only emergency cohort (fresh-400): replacement primary emergency analysis and cross-stack bridge
- Archived superseded text (separate file, not part of this supplement)

**Multi-agent LLM teams improve differential recall on curated cases but underperform on emergency working-label matching: a two-cohort, two-family evaluation**

**Version and provenance.** This peer-review version retains Supplementary Methods, Tables S1–S17, Supplementary Figures S1–S3, and Supplementary Notes S34–S39. Earlier drafts and the archived Notes S18–S33 are preserved separately; their superseded numerical results are not part of this supplement.

Unless a table states otherwise, reported values are offline recomputations from the frozen per-case outputs and judge caches, with the case as the unit of inference and five-seed mean hit rates. Current authoritative artifacts take precedence over interim run logs: emergency analyses use the self-hosted rerun and routing_study/results/local_rerun/; sensitivity-judge case-level contrasts use routing_study/results/s2b_caselevel_ds.json; the same-325 PMC analysis uses pmc_harvest/results/overnight/same325_format_analysis.md; and symmetric thinking analyses use routing_study/results/sym_thinking_{cpc,mcr}.{json,md}. Cache totals and dated run histories are retained in the project archive rather than repeated here.

**Cross-reference map.**

| Item | Content |
| --- | --- |
| Supplementary Methods | Full prompts, model identifiers, serving configuration, and judge rules |
| S1–S4 | Per-seed results; sensitivity-judge results; legacy pooled-seed comparisons; multiplicity correction |
| S5, S8 | Judge–clinician agreement and judge–judge agreement, respectively |
| S6, S9 | Workup-informed emergency analyses, including exclusion of encounters without objective content |
| S7 | Held-out and development CPC subsets |
| S10 | Matched controls and factorial/moderator ablations, including emergency and MedCaseReasoning extensions; S10f-2 is withdrawn |
| S11, S12, S17 | Second-family and same-cohort emergency replication; backbone scale; closed-flagship CPC extension |
| S13, S14 | Prompt-transcription audit/control and data-integrity repair |
| S15, S16 | GEE and majority-vote-threshold sensitivity analyses |
| Figure S1 | Top-k performance across corpora and emergency input conditions |
| Unnumbered notes | Emergency sampling/input construction and temperature-matched analyses |
| S34 | Judge-independent word coverage and differential misclassification risk |
| S35 | Same-325 full/condensed contrasts, replication, mediation, and cross-corpus descriptive heterogeneity |
| S36 | Emergency output-contract sensitivity arm |
| S37 | Exploratory blinded physician ratings and inter-rater reliability |
| S38 | Asymmetric and fully symmetric thinking-mode sensitivity analyses |
| S39 | Protocol-frozen disease-label-only replacement emergency cohort (fresh-400): primary emergency analysis, workup and output-contract arms, second-family replication, cross-stack bridges, judge revalidation, and replacement-cohort physician rating |

### Supplementary Methods: full prompt text

**Model identifiers and serving configuration.**

| Model | Role | Serving path and identifier | Notes |
| --- | --- | --- | --- |
| qwen3.8-flash — ModelScope identifier **Qwen3.8-Flash-Next** (176B, provider-documented) | backbone, all primary arms and control arms | curated corpora: provider direct API (requests 2026-09-10 to 09-19); emergency rerun and output-contract arm: self-hosted open-weight deployment, 4-bit weight-only quantization (2026-10-02 to 04 and 2026-10-17 to 20), thinking disabled | per-request logs retained; the two deployments are byte-identical in prompts (the reversal’s magnitude shifted between them — S2b-2 serving-stack note) |
| deepseek-v4.1-flash — ModelScope identifier **DeepSeek-V4.1-Flash** | second model family; sensitivity judge | family: provider API under its serving alias deepseek-flash; emergency replication: OpenRouter zero-data-retention route pinned to one provider (zdr: true, data_collection: deny, fallbacks disabled), all 15,141 requests logged; sensitivity judge: frozen v3 rules, temperature 0, thinking disabled | the alias-versus-identity distinction is documented in the released pipeline |
| gpt-5.1 (closed flagship) | family extension | OpenAI-compatible endpoint, snapshot gpt-5.1-2025-11-13 (the protocol-frozen replication-cohort arm), five seeds | not independently reproducible; all gpt-5.1 adjudications rest on the primary judge (S17 quality-control note) |
| GLM-5.3-flash — ModelScope identifier **GLM-5.3-Flash** | primary judge | provider official endpoint (open.bigmodel.cn), thinking enabled at low reasoning effort (8,192-token budget) — a model constraint; frozen v3 rules; one cached verdict per candidate–reference pair (key = gold[:150]+"\|\|"+cand[:150]) | cache 253,005 pairs at the last adjudication, 0 unresolved |

Model identifiers confirmed against ModelScope (2026-10-21); serving aliases (the lowercase API strings used in the pipeline) are retained in the released scripts.

All prompts are reproduced verbatim from the analysis code. Curly-brace tokens ({case_text}, {structured_case}, {gold}, {pred}, role fields) are the Python format placeholders filled per case at runtime. Thinking mode was disabled for all strategy calls (A×1, the A×1+CoT control of S-M5, P, and both MDT stages, in both model families) and for the deepseek-v4.1-flash sensitivity judge. The GLM-5.3-flash judge cannot be run with thinking disabled and was therefore called with thinking enabled at low reasoning effort (8,192-token budget), a model constraint rather than a design choice. Decoding: A×1 requested temperature 0 and P, the MDT role agents, and the MDT moderator requested temperature 0·3. Temperature 0 is a request for greedy decoding rather than a guarantee of it, because the serving endpoint is not fully deterministic; each case was therefore run five times, and the case, not the individual run, is the unit of inference (main text, Models and decoding).

#### S-M1. Strategy A×1 (single-pass direct prompting)

Source: A_TOPN_PROMPT in routing_study/scripts/topn_cpc_promptv2.py.

You are an expert internist reviewing an MGH CPC case.

Produce a ranked differential diagnosis: the 5 most likely diagnoses, most likely first. Be specific (disease name plus the key qualifier that matters for this case).

Guidelines:
- The diagnosis under question is not necessarily a structural/organic lesion: a diagnosis already stated in the history (including a psychiatric diagnosis), or the main reason for the current admission, may itself be the answer to determine.
- A striking organic finding (mass, lesion) may be an incidental companion finding; if two separate diagnostic lines exist, evaluate both and rank each by how well it explains the whole case.
- Combination diagnoses are allowed when they reflect the real process (e.g., "post-influenza bacterial superinfection pneumonia"). For infectious candidates, name specific pathogens as separate items when clinically distinct (e.g., mucormycosis vs aspergillosis).

Respond with ONLY a JSON object, no other text:
{"top5": [{"rank": 1, "diagnosis": "..."}, {"rank": 2, "diagnosis": "..."}, {"rank": 3, "diagnosis": "..."}, {"rank": 4, "diagnosis": "..."}, {"rank": 5, "diagnosis": "..."}]}

Case:
{case_text}

#### S-M2. Strategy P (five personas in one context)

Source: PERSPECTIVE_PROMPT in scripts/scheme_perspective.py, concatenated with P_TOPN_SUFFIX in routing_study/scripts/topn_cpc_promptv2.py. The suffix overrides the closing output format so that all strategies return the same ranked top-5 JSON.

**Fidelity note.** Part 2 below is the *intent* version of the suffix, printed with single braces as it would read after Python .format() folding. In the code the suffix is a plain string that is never passed through .format(), so the text actually sent to the model carries literal doubled braces ({{, }}) in that JSON example; the byte-exact sent text, with SHA-256 fingerprints of each constant, is deposited as routing_study/results/prompt_actually_sent.md. The defect, its effect on the second model family, and the corrected-suffix control are described in the main text Methods (“Prompt transcription defect and its control”), in the Supplementary Note on the prompt-transcription defect (S13), and, for the control arm itself, in Supplementary Table S10c; the disclosed behaviour is the one actually used for every P-arm number reported here.

Part 1 — perspective prompt:

```
You are an expert diagnostic team analyzing a complex medical case from multiple complementary perspectives. Each perspective contributes a brief analysis; you then synthesize them into a single final diagnosis.

## Case Presentation
{structured_case}

## Diagnostic Perspectives

Analyze the case from each of the following perspectives. Be concise (2-4 sentences each).

**1. Attending Physician Perspective**
What is the most coherent unifying diagnosis? Which clinical features are most discriminating?

**2. Pathophysiology Perspective**
What underlying mechanism could produce this constellation of findings? Are there hallmark laboratory, histologic, or molecular clues?

**3. Imaging and Laboratory Specialist Perspective**
How should the objective data (imaging, labs, vitals, procedures) be interpreted? What patterns or paradoxes stand out?

**4. Epidemiology and Exposure Perspective**
What role do demographics, geography, travel, diet, medications, toxins, occupational exposures, or comorbidities play? Are there hidden risk factors in the narrative?

**5. Skeptic / Challenger Perspective**
What is the strongest argument AGAINST the leading diagnosis? What alternative diagnoses could explain MORE findings with FEWER contradictions? What findings remain unexplained?

## Synthesis Task
Integrate the five perspectives above and provide:
- The single most likely diagnosis
- A ranked differential diagnosis (3-5 alternatives)
- Brief reasoning explaining how the multi-perspective analysis led to your choice

Respond in EXACTLY this format:
MOST_LIKELY_DIAGNOSIS: [final best diagnosis]
DIFFERENTIAL_DIAGNOSIS: [differential 1], [differential 2], [differential 3], [differential 4]
REASONING: [concise multi-perspective synthesis]
CONFIDENCE_SCORE: [0-100]
```

Part 2 — appended top-5 suffix (same guidelines block as A×1):

Guidelines:
- The diagnosis under question is not necessarily a structural/organic lesion: a diagnosis already stated in the history (including a psychiatric diagnosis), or the main reason for the current admission, may itself be the answer to determine.
- A striking organic finding (mass, lesion) may be an incidental companion finding; if two separate diagnostic lines exist, evaluate both and rank each by how well it explains the whole case.
- Combination diagnoses are allowed when they reflect the real process (e.g., "post-influenza bacterial superinfection pneumonia"). For infectious candidates, name specific pathogens as separate items when clinically distinct (e.g., mucormycosis vs aspergillosis).

\## Output Requirement (final block)
After your five-perspective analysis, conclude with one JSON code block (the only JSON in your answer):
{"top5": [{"rank": 1, "diagnosis": "..."}, {"rank": 2, "diagnosis": "..."}, {"rank": 3, "diagnosis": "..."}, {"rank": 4, "diagnosis": "..."}, {"rank": 5, "diagnosis": "..."}]}
Exactly 5 items, ranked most to least likely. Nothing may follow the JSON block.

#### S-M3. Strategy MDT (isolated roles + moderator)

Source: ROLES and the prompt builders in routing_study/scripts/mdt_cpc.py. Each role is issued as an independent call that cannot see any other role’s output; a moderator call then integrates the five opinions (six calls per case in total). Role calls and the moderator call used temperature 0·3 and max_tokens 4096.

The five roles (title — assigned lens), verbatim:

1. **Attending Internist** — “overall clinical synthesis: which unifying diagnosis best explains the whole case, and which features are most discriminating”
2. **Pathophysiologist** — “underlying mechanism: what process could produce this constellation of findings, and hallmark histologic, laboratory, or molecular clues”
3. **Imaging & Laboratory Specialist** — “objective data: how to interpret the imaging, laboratory, and vital-sign patterns, and which patterns or paradoxes stand out”
4. **Epidemiologist** — “demographics, geography, travel, exposures, medications, occupation, and comorbidities — hidden risk factors in the narrative”
5. **Skeptic / Challenger** — “the strongest argument AGAINST the leading hypothesis: alternatives that explain more findings with fewer contradictions, and findings that remain unexplained”

Role-agent prompt template ({role_title} and {role_brief} filled from the list above):

You are the {role_title} on an MDT panel reviewing an MGH CPC case.

Your assigned lens: {role_brief}

Through this lens, list the 5 candidate diagnoses that best fit the case, ranked most to least likely FROM YOUR PERSPECTIVE. Include candidates another specialist might overlook if the findings support them.

Guidelines:
- A diagnosis already stated in the history (including a psychiatric diagnosis), or the main reason for the current admission, may itself be the answer.
- A striking organic finding may be an incidental companion finding.
- Combination diagnoses are allowed. For infections, name specific pathogens as separate items when clinically distinct.

Respond with ONLY a JSON object, no other text:
{"top5": [{"rank": 1, "diagnosis": "...", "rationale": "one line through your lens"}, ... exactly 5 items]}

Case:
{case_text}

Moderator (synthesis) prompt template ({opinions_text} lists each role’s ranked candidates with one-line rationales):

You are the moderator of an MDT panel. Five specialists independently reviewed the MGH CPC case below, each through their own lens, without seeing each other's opinions. Their ranked candidate lists are given.

Integrate them into the final ranked top-5 for the case:
- Candidates supported by multiple specialists generally rise.
- A unique candidate with specific, case-grounded support must NOT be dropped merely because only one specialist listed it.
- Resolve conflicts by re-checking against the case text.
- Combination diagnoses are allowed.

Respond with ONLY a JSON object, no other text:
{"top5": [{"rank": 1, "diagnosis": "..."}, ... exactly 5 items]}

Case:
{case_text}

Panel opinions:
{opinions_text}

#### S-M4. LLM judge, rule version 3 (primary and sensitivity judges)

Source: V3_PROMPT in routing_study/scripts/judge_v3.py. The same prompt was used for the primary judge (GLM-5.3-flash) and the sensitivity judge (deepseek-v4.1-flash), at temperature 0.

```
You are a medical evaluation judge. A model produced a diagnosis for a clinical case.
Reference (correct) diagnosis: {gold}
Model's diagnosis: {pred}

Judging rules (apply in order):
1. CORRECT (YES) if the model's diagnosis names the same DISEASE as the reference —
   synonyms, abbreviations, and translations are acceptable.
2. The model adding extra findings, complications, etiologies, or secondary diagnoses
   does NOT make it wrong, as long as the reference disease is named as (part of) the
   main diagnosis.
3. Missing qualifiers in the reference (disease stage, severity, "in remission",
   anatomic subtype, etiologic form) do NOT make the model wrong.
3b. A BROADER disease-family or category term that clearly includes the reference
   disease as one of its subtypes or variants is CORRECT (e.g., "lymphoma" for
   "Hodgkin lymphoma"; "thyroiditis" for "Hashimoto thyroiditis"; "muscular
   dystrophy" for "limb-girdle muscular dystrophy type 2B").
3c. Naming the dominant component of a COMPOUND reference diagnosis is CORRECT
   (e.g., "autoimmune hemolytic anemia" for "Evans syndrome (AIHA +
   thrombocytopenia)"; "CMV colitis" for "CMV colitis and inflammatory bowel disease").
3d. A NEIGHBORING or less-specified subtype within the same disease entity is
   CORRECT (e.g., "borderline lepromatous leprosy" for "lepromatous leprosy";
   "basaloid squamous cell carcinoma" for "squamous cell carcinoma").
4. WRONG (NO) only if the model names a genuinely DIFFERENT disease entity as its
   main diagnosis — a distinct entity, not merely a broader category, a component
   of a compound diagnosis, or a neighboring subtype.
Answer with exactly one word: YES or NO.
```

Rules 3b–3d are the v3 additions motivated by error analysis of the 19 judge–human discordant pairs in validation batch 1 (see Supplementary Table S5).

#### S-M5. Control arm A×1+CoT (single call with elicited reasoning)

Source: A_TOPN_COT_PROMPT in routing_study/scripts/ax1_cot_cpc.py. This arm isolates the effect of eliciting reasoning text from the effect of the multi-perspective structure: it is the same single call as A×1, with the same guidelines, JSON schema, model, temperature (0), case text, and call count, and the only change is the request for a short step-by-step justification before the list. max_tokens is 4,096 here against 2,048 for A×1, so that the justification is not truncated; the JSON code block is fenced, as in P, because prose and parseable JSON have to coexist in one answer.

You are an expert internist reviewing an MGH CPC case.

Produce a ranked differential diagnosis: the 5 most likely diagnoses, most likely first. Be specific (disease name plus the key qualifier that matters for this case).

Guidelines:
- The diagnosis under question is not necessarily a structural/organic lesion: a diagnosis already stated in the history (including a psychiatric diagnosis), or the main reason for the current admission, may itself be the answer to determine.
- A striking organic finding (mass, lesion) may be an incidental companion finding; if two separate diagnostic lines exist, evaluate both and rank each by how well it explains the whole case.
- Combination diagnoses are allowed when they reflect the real process (e.g., "post-influenza bacterial superinfection pneumonia"). For infectious candidates, name specific pathogens as separate items when clinically distinct (e.g., mucormycosis vs aspergillosis).

First reason through the case in a concise step-by-step analysis (5-10 sentences):
1. State the discriminating features: the key positives and the key negatives.
2. Generate candidate diagnoses and weigh them against each other on how well each explains the whole picture.
3. Explain why the leading candidate is favoured over the strongest alternatives.
Then finish with exactly one JSON code block (the only JSON in your answer), preceded by no other JSON and followed by nothing:
```json
{"top5": [{"rank": 1, "diagnosis": "..."}, {"rank": 2, "diagnosis": "..."}, {"rank": 3, "diagnosis": "..."}, {"rank": 4, "diagnosis": "..."}, {"rank": 5, "diagnosis": "..."}]}

Exactly 5 items, ranked most to least likely.

Case: {case_text}

The JSON example is shown here with single braces because the constant is passed through `.format()`, as for A×1. The per-call justification length is recorded in the released run files (`reasoning_chars`), and Table S10a reports the results.

```{=openxml}
<w:p><w:r><w:br w:type="page"/></w:r></w:p>

#### S-M6. Condensed-report construction, filler arm, and replication-cohort protocol

To compare input forms on fixed content, we harvested full narrative case reports from PubMed Central (514 reports) and rewrote them into teaching vignettes (180–250 words, ending before the diagnosis) under a condenser prompt forbidding disclosure or implication of the reference diagnosis; every condensate was screened against its reference by two judge models under a zero-tolerance criterion. A filler arm re-elongated the 325 survivors to original lengths with neutral, diagnosis-free padding (seed 1). The replication cohort was expanded to 392 under a protocol-frozen plan (v2.6–v2.7; self-frozen, not externally registered), fixed before any expansion outcome was computed: three predefined supply windows, a frozen stopping rule, unchanged instruments, and vignette-stage inclusion by dual audit. Every replication publication postdates the evaluated model’s release. Per-window presentation-audit rates differed with source-article style and are disclosed as selection asymmetry, not as an inclusion criterion (Supplementary Note S35). Mediation analyses, filler arm, and expansion were protocol-frozen before outcomes were computed; seeds were five except the exploratory filler.

#### S-M7. Models, decoding, and compliance routing

Every ER-Reason arm (all conditions and factorials, five seeds each) was regenerated on self-hosted infrastructure (2026-10-02 to 04; open-weight 4-bit), with prompts byte-identical to the curated runs and no case content leaving the machine. The deepseek-v4.1-flash same-cohort emergency replication—the one emergency pathway sending case text off-machine—ran through OpenRouter with zero-data-retention routing pinned to a single provider (zdr: true, data_collection: deny, fallbacks disabled); all 15,141 generation requests were logged and served by the pinned endpoint. Requested temperature 0 is not guaranteed greedy on cloud endpoints, so the case, not the run, is the unit of inference; self-hosted it is exact greedy. A×1 ran at requested temperature 0 and the multi-perspective arms at 0·3; a temperature-matched reanalysis at 0·3 left the main conclusions unchanged (Tables 1 and 2; appendix). A protocol-frozen output-contract sensitivity arm appended one sentence to each prompt, permitting symptom-level entries over unsupported disease guesses; seeds and judge were unchanged (Supplementary Note S36).

#### S-M8. Workup-informed input condition

To test whether the emergency result changed with additional information, we appended each encounter’s objective results to its presentation (prompts, model, decoding, seeds, judge unchanged). Rule-based filters blind to the reference retained laboratory values, vital signs, and imaging impressions and removed assessment, plan, and disposition text; manual review found no asserted final diagnosis. 240 cases (65·9%) had ≥200 characters of objective content and 34 none (conditions identical by construction); 20 label-in-text cases were excluded in a sensitivity analysis.

#### S-M9. Blinded physician rating of the discordant pairs (original cohort)

Three non-author senior physicians (≥1 emergency medicine) independently rated all 34 emergency cases where the strategies’ top-1 verdicts disagreed (13 symptom-level, 21 disease-level; all-take rule) plus 30 concordant controls. Each unit showed the case with two anonymised top-5 lists in random A/B order; raters were blind to strategy, reference, hypotheses, and each other. Instruments addressed which list’s errors were more dangerous (primary), dangerous-omission and misleading-inclusion flags, acceptance and first-position judgements, and workup sufficiency (seed-1 lists, the pre-specified display). Protocol, randomisation seed, and the low-agreement descriptive-path amendment were frozen before any rating was viewed; the agreement gate (Gwet AC1 ≥0·6) was evaluated before unblinding (Supplementary Note S37).

#### S-M10. Blinded physician rating on the replacement cohort

The safety assessment was repeated on the replacement primary cohort under a protocol revision (v1.3) frozen on 2026-11-05 before any rating material existed, with primary and secondary questions inverted relative to the original rating: the primary question was case-level mechanism verification—did the losing strategy’s list contain an acceptable entry encompassing the reference diagnosis (undifferentiated-but-pointing, superordinate, or aetiologic-chain formulations)?—and the secondary was the danger comparison, retained with structured operational anchors after the original rating’s danger agreement collapse. All 53 top-1 majority-vote-discordant replacement-cohort cases (35 A×1 wins, 18 MDT wins), 30 concordant-hit controls, and 15 concordant-miss baselines were rated, plus a 10-unit calibration batch and 14 embedded duplicate units (175 units); three non-author senior physicians were blind to strategy, reference, hypotheses, and each other, with the agreement gate again evaluated before unblinding. Part A showed two anonymised seed-1 lists per case (danger comparison, omission and misleading-inclusion flags, initial-differential preference); Part B, after reference revelation, showed the losing arm’s list for the rescue judgement (Supplementary Note S39).

### Supplementary Table S1. Per-seed accuracy of the three strategies (primary judge, GLM-5.3-flash × v3)

Recomputed offline from the raw outputs against the frozen primary judge cache by routing_study/scripts/make_supplement_tables.py; the ER-Reason panel uses make_supplement_tables_local.py and the self-hosted rerun. Values are percentages of cases with the reference diagnosis within the top-k entries. Mean ± SD is across five seeds, and all tabulated runs have complete primary-judge adjudication.

**CPC (n = 87), five seeds**

| Strategy | Metric | Seed 1 | Seed 2 | Seed 3 | Seed 4 | Seed 5 | Mean ± SD |
| --- | --- | --- | --- | --- | --- | --- | --- |
| A×1 | Top-1 | 56·3 | 58·6 | 57·5 | 56·3 | 60·9 | 57·9 ± 1·9 |
| A×1 | Top-3 | 72·4 | 80·5 | 71·3 | 77·0 | 79·3 | 76·1 ± 4·1 |
| A×1 | Top-5 | 77·0 | 83·9 | 80·5 | 81·6 | 82·8 | 81·1 ± 2·6 |
| P | Top-1 | 51·7 | 51·7 | 60·9 | 52·9 | 51·7 | 53·8 ± 4·0 |
| P | Top-3 | 69·0 | 72·4 | 74·7 | 70·1 | 74·7 | 72·2 ± 2·6 |
| P | Top-5 | 74·7 | 77·0 | 79·3 | 74·7 | 78·2 | 76·8 ± 2·1 |
| MDT | Top-1 | 63·2 | 64·4 | 58·6 | 58·6 | 57·5 | 60·5 ± 3·1 |
| MDT | Top-3 | 83·9 | 83·9 | 78·2 | 80·5 | 81·6 | 81·6 ± 2·4 |
| MDT | Top-5 | 87·4 | 87·4 | 82·8 | 83·9 | 86·2 | 85·5 ± 2·1 |

**MCR (n = 406), five seeds**

| Strategy | Metric | Seed 1 | Seed 2 | Seed 3 | Seed 4 | Seed 5 | Mean ± SD |
| --- | --- | --- | --- | --- | --- | --- | --- |
| A×1 | Top-1 | 56·2 | 56·4 | 57·1 | 58·4 | 57·1 | 57·0 ± 0·9 |
| A×1 | Top-3 | 70·4 | 69·5 | 70·4 | 70·0 | 69·2 | 69·9 ± 0·6 |
| A×1 | Top-5 | 75·4 | 75·1 | 76·4 | 76·6 | 74·6 | 75·6 ± 0·8 |
| P | Top-1 | 56·7 | 56·4 | 56·4 | 55·9 | 56·7 | 56·4 ± 0·3 |
| P | Top-3 | 68·0 | 70·7 | 70·0 | 68·5 | 68·7 | 69·2 ± 1·1 |
| P | Top-5 | 73·6 | 74·9 | 74·9 | 76·4 | 75·4 | 75·0 ± 1·0 |
| MDT | Top-1 | 55·9 | 55·7 | 54·9 | 57·1 | 55·9 | 55·9 ± 0·8 |
| MDT | Top-3 | 72·9 | 72·4 | 73·4 | 73·9 | 71·9 | 72·9 ± 0·8 |
| MDT | Top-5 | 78·3 | 79·8 | 80·3 | 80·0 | 79·3 | 79·6 ± 0·8 |

**ER-Reason (n = 364), five seeds**

| Strategy | Metric | Seed 1 | Seed 2 | Seed 3 | Seed 4 | Seed 5 | Mean ± SD |
| --- | --- | --- | --- | --- | --- | --- | --- |
| A×1 | Top-1 | 40·7 | 40·7 | 40·7 | 40·7 | 40·7 | 40·7 ± 0·0 |
| A×1 | Top-3 | 54·4 | 54·4 | 54·4 | 54·4 | 54·4 | 54·4 ± 0·0 |
| A×1 | Top-5 | 61·3 | 61·3 | 61·3 | 61·3 | 61·3 | 61·3 ± 0·0 |
| P | Top-1 | 38·7 | 40·7 | 38·7 | 41·5 | 40·1 | 39·9 ± 1·2 |
| P | Top-3 | 51·4 | 48·4 | 50·3 | 51·4 | 49·5 | 50·2 ± 1·3 |
| P | Top-5 | 57·1 | 54·1 | 56·6 | 56·9 | 56·3 | 56·2 ± 1·2 |
| MDT | Top-1 | 38·2 | 36·8 | 36·8 | 35·2 | 36·8 | 36·8 ± 1·1 |
| MDT | Top-3 | 49·7 | 50·0 | 47·8 | 46·4 | 45·9 | 48·0 ± 1·9 |
| MDT | Top-5 | 55·2 | 56·6 | 53·8 | 54·7 | 54·1 | 54·9 ± 1·1 |

The ER-Reason panel uses the self-hosted rerun with byte-identical prompts. Its A×1 rows have SD = 0 because exact greedy decoding produced identical outputs across the five seed directories. The MCR MDT rows use the repaired runs described in S14.

### Supplementary Table S2. Main results under the sensitivity judge (deepseek-v4.1-flash × v3 rules)

Values are means ± SD across five seeds. MCR uses the repaired MDT outputs and ER-Reason uses the self-hosted rerun, scored against the frozen sensitivity-judge cache (routing_study/results/judge_cache_dsflash_v3.json) under the identical v3 rules. Judge inputs contain diagnosis strings only, not case content. McNemar entries give discordant-pair counts (first-named strategy only correct : second only correct) with two-sided exact p values from the seed-pooled legacy analysis; S2b-2 gives the current case-level framework.

**S2a. Accuracy / recall (%)**

| Dataset | Strategy | Top-1 | Top-3 | Top-5 |
| --- | --- | --- | --- | --- |
| CPC (n = 87) | A×1 | 51·7 ± 1·8 | 72·6 ± 3·8 | 77·7 ± 2·6 |
|  | P | 50·1 ± 3·8 | 69·7 ± 2·6 | 75·2 ± 1·9 |
|  | MDT | 57·0 ± 2·1 | 78·4 ± 1·5 | 82·3 ± 1·3 |
| MCR (n = 406, five seeds) | A×1 | 53·1 ± 0·8 | 66·1 ± 0·6 | 72·0 ± 1·1 |
|  | P | 53·1 ± 1·0 | 65·0 ± 1·2 | 70·0 ± 1·2 |
|  | MDT | 52·7 ± 0·4 | 68·5 ± 0·9 | 74·4 ± 1·2 |
| ER-Reason (n = 364, five seeds) | A×1 | 33·2 ± 0·0 | 44·5 ± 0·0 | 48·9 ± 0·0 |
|  | P | 33·4 ± 1·2 | 41·6 ± 1·3 | 46·2 ± 1·0 |
|  | MDT | 31·1 ± 0·5 | 39·5 ± 1·5 | 44·9 ± 1·9 |

**S2b. Paired comparisons (sensitivity judge)**

| Dataset | Comparison | Top-1 | Top-3 | Top-5 |
| --- | --- | --- | --- | --- |
| CPC (n = 87) | MDT vs A×1 | 44:21, p = 0·0059 | p = 0·0022 | p = 0·0055 |
| CPC held-out (n = 46) | MDT vs A×1 | 30:10, p = 0·0022 | — | — |
| MCR (n = 406) | MDT vs A×1 | p = 0·62 | 127:79, p = 0·0010 | 118:68, p = 0·0003 |
| ER-Reason (n = 364, five seeds, self-hosted rerun) | A×1 vs MDT | 328:250, p = 0·0013 | 437:253, p = 2·4 × 10⁻¹² | 469:323, p = 2·4 × 10⁻⁷ |

**S2b-2. The same contrasts under the case-level framework (paired two-sided Wilcoxon on per-case five-seed hit rates; sensitivity judge).**

| Dataset | MDT − A×1 | Top-1 | Top-3 | Top-5 |
| --- | --- | --- | --- | --- |
| CPC (n = 87 at every endpoint) | diff, p | +5·3, p = 0·098 | +5·7, p = 0·020 | +4·6, p = 0·011 |
| MCR (n = 405 / 401 / 399 at top-1/3/5) | diff, p | −0·4, p = 0·70 | +2·3, p = 0·034 | +2·5, p = 0·022 |
| ER-Reason (n = 364 at every endpoint) | diff, p | −2·1, p = 0·22 | −5·1, p = 0·0073 | −4·0, p = 0·064 |

The pooled-McNemar cells of S2b follow the legacy framework retired in Table S3 and are retained for continuity; the case-level readings agree in direction with every pooled cell. Under the case-level framework the MCR and ER top-3 contrasts remain significant, the ER top-5 contrast sits at p = 0·064 (the primary judge gives p = 0·0039 there), and CPC top-1 is not significant. Seven MCR cases have some incomplete sensitivity-judge adjudication in the frozen second cache; complete-case denominators therefore vary by endpoint (405, 401, and 399). Computed offline by routing_study/scripts/s2b_caselevel_ds.py (routing_study/results/s2b_caselevel_ds.json). **Serving-stack note.** The primary-judge emergency top-3 deficit was −9·0 points on the cloud-stack runs and −6·4 on the self-hosted 4-bit stack, while the second family’s same-cohort replication showed little change across its provider paths (43·4 versus 43·5; S11c). Quantisation is a candidate explanation for the qwen magnitude shift, not an isolated cause; ordering was preserved across stacks.

The two judges give concordant directions for the MDT recall contrasts on CPC and MCR, the original P-arm deficits in the tested prompt/model context, and the ER-Reason reversal. This is a same-rule sensitivity analysis, not independent clinical validation. The MDT top-1 advantage on CPC (including the held-out subset) is significant under the sensitivity judge but not under the primary judge, so no top-1 superiority claim is made. The MCR rows use the repaired MDT runs described in S14.

### Supplementary Table S3. Legacy pooled-seed McNemar statistics versus the primary case-level analysis

The legacy analysis pooled case–seed pairs into a single McNemar test (435 pairs for CPC, 2,030 for MCR), treating repeated seeds of the same case as independent. Those figures are superseded by the case-level analysis (per-case five-seed hit rates, paired Wilcoxon signed-rank test; main text, Statistical analysis) and are reproduced here for reference only. a:b = discordant pairs (first-named strategy only correct : second only correct), two-sided exact p.

**CPC (n = 87)**

| Comparison | Metric | Legacy pooled McNemar (a:b, p) | Primary: Wilcoxon p | Conclusion change |
| --- | --- | --- | --- | --- |
| MDT vs A×1 | Top-1 | 34:23, p = 0·1849 | 0·4283 | None (both n.s.) |
| MDT vs A×1 | Top-3 | 37:13, p = 0·0009 | 0·0079 | None (both sig.) |
| MDT vs A×1 | Top-5 | 27:8, p = 0·0019 | 0·0151 | None (both sig.) |
| MDT vs P | Top-1 | 55:26, p = 0·0017 | 0·0744 | Sig. → n.s. |
| MDT vs P | Top-3 | 61:20, p < 0·0001 | 0·0043 | None (both sig.) |
| MDT vs P | Top-5 | 52:14, p < 0·0001 | 0·0036 | None (both sig.) |
| P vs A×1 | Top-1 | 25:43, p = 0·0385 | 0·2350 | Sig. → n.s. |
| P vs A×1 | Top-3 | 26:43, p = 0·0533 | 0·1981 | None (both n.s.) |
| P vs A×1 | Top-5 | 24:43, p = 0·0271 | 0·1225 | Sig. → n.s. |

**MCR (n = 406)**

| Comparison | Metric | Legacy pooled McNemar (a:b, p) | Primary: Wilcoxon p | Conclusion change |
| --- | --- | --- | --- | --- |
| MDT vs A×1 | Top-1 | 91:114, p = 0·1242 | 0·2528 | None (both n.s.) |
| MDT vs A×1 | Top-3 | 129:68, p < 0·0001 | 0·0079 | None (both sig.) |
| MDT vs A×1 | Top-5 | 127:47, p < 0·0001 | < 0·0001 | None (both sig.) |
| MDT vs P | Top-1 | 110:120, p = 0·5530 | 0·5472 | None (both n.s.) |
| MDT vs P | Top-3 | 159:83, p < 0·0001 | 0·0027 | None (both sig.) |
| MDT vs P | Top-5 | 174:82, p < 0·0001 | 0·0001 | None (both sig.) |
| P vs A×1 | Top-1 | 110:123, p = 0·4318 | 0·7593 | None (both n.s.) |
| P vs A×1 | Top-3 | 126:141, p = 0·3916 | 0·7005 | None (both n.s.) |
| P vs A×1 | Top-5 | 123:135, p = 0·4935 | 0·8364 | None (both n.s.) |

Under the case-level analysis the CPC persona penalty (P vs A×1) is no longer significant, and that claim is carried by the ER-Reason results instead; all MDT recall advantages are unchanged. The MedCaseReasoning rows carry the re-synthesised MDT runs of the data-integrity repair (Supplementary Note S14), which is why its discordant-pair counts differ by one or two pairs from the pre-repair values; the CPC and ER-Reason rows are unaffected, and the primary case-level conclusions of this table are the same before and after the repair. ER-Reason formerly used a single-seed exact McNemar analysis; it now uses the same case-level framework as the other two datasets, so no pooled-seed figures enter the main text for any dataset and the case-level results of Table 1 carry its tests.

### Supplementary Table S4. Benjamini–Hochberg correction across the 27 tests of the primary analysis framework

Recomputed by routing_study/scripts/make_supplement_tables_local.py (rerun 2026-10-05). p values are paired Wilcoxon signed-rank (two-sided) values on per-case five-seed hit rates for all three datasets: CPC and MCR from routing_study/results/stats_caselevel.json (unchanged), ER-Reason from the self-hosted rerun (routing_study/results/local_rerun/topn_erreason/seed_summary.json, written 2026-10-05). q values are Benjamini–Hochberg adjusted; rows sorted by p.

Rank | Dataset | Comparison | Endpoint | p | q (BH) |
1 | MCR | MDT vs A×1 | Top-5 | 3·75 × 10⁻⁵ | 0·001 |
2 | MCR | MDT vs P | Top-5 | 1·03 × 10⁻⁴ | 0·0014 |
3 | ER-Reason | MDT vs A×1 | Top-3 | 0·0022 | 0·0166 |
4 | MCR | MDT vs P | Top-3 | 0·0027 | 0·0166 |
5 | CPC | MDT vs P | Top-5 | 0·0036 | 0·0166 |
6 | ER-Reason | MDT vs A×1 | Top-5 | 0·0039 | 0·0166 |
7 | CPC | MDT vs P | Top-3 | 0·0043 | 0·0166 |
8 | CPC | MDT vs A×1 | Top-3 | 0·0079 | 0·0237 |
9 | MCR | MDT vs A×1 | Top-3 | 0·0079 | 0·0237 |
10 | ER-Reason | P vs A×1 | Top-5 | 0·0104 | 0·028 |
11 | CPC | MDT vs A×1 | Top-5 | 0·0151 | 0·0371 |
12 | ER-Reason | MDT vs A×1 | Top-1 | 0·0215 | 0·0484 |
13 | ER-Reason | P vs A×1 | Top-3 | 0·0257 | 0·0534 |
14 | ER-Reason | MDT vs P | Top-1 | 0·0383 | 0·0738 |
15 | CPC | MDT vs P | Top-1 | 0·0744 | 0·134 |
16 | CPC | P vs A×1 | Top-5 | 0·1225 | 0·2068 |
17 | CPC | P vs A×1 | Top-3 | 0·1981 | 0·3146 |
18 | CPC | P vs A×1 | Top-1 | 0·235 | 0·3525 |
19 | MCR | MDT vs A×1 | Top-1 | 0·2528 | 0·3592 |
20 | ER-Reason | MDT vs P | Top-3 | 0·3067 | 0·4141 |
21 | CPC | MDT vs A×1 | Top-1 | 0·4283 | 0·5507 |
22 | MCR | MDT vs P | Top-1 | 0·5472 | 0·6716 |
23 | ER-Reason | MDT vs P | Top-5 | 0·7005 | 0·7881 |
24 | MCR | P vs A×1 | Top-3 | 0·7005 | 0·7881 |
25 | MCR | P vs A×1 | Top-1 | 0·7593 | 0·8201 |
26 | ER-Reason | P vs A×1 | Top-1 | 0·8124 | 0·8364 |
27 | MCR | P vs A×1 | Top-5 | 0·8364 | 0·8364 |

Fourteen of the 27 tests have unadjusted p < 0·05; twelve of them remain significant after correction. The two exceptions are both emergency-corpus contrasts at endpoints that carry no primary claim in the main text: MDT-versus-P top-1 (p = 0·0383, q = 0·0738) and P-versus-A×1 top-3 (p = 0·0257, q = 0·0534), both reported as nominal throughout. The pre-specified team-versus-single-call contrast (MDT versus A×1) survives at every endpoint on every dataset, including ER-Reason top-1 at q = 0·0484. Four rows carry the re-synthesised MedCaseReasoning MDT runs of the data-integrity repair (Supplementary Note S14): the MDT-versus-A×1 top-3 contrast moves from p = 0·0055 to 0·0079 and MDT-versus-P top-3 from p = 0·0018 to 0·0027, the corresponding top-1 contrasts move from 0·2675 to 0·2528 and from 0·5605 to 0·5472, and the rank order of rows 11–12, 22 and 24 changes accordingly; all four remain on the same side of significance as before the repair.

### Supplementary Table S5. Judge validation: Cohen’s κ against two blinded clinician raters

Values from the locked evidence bundle (paper/evidence_bundle.md, §3). Each cell gives κ versus rater A / κ versus rater B. Batch 1 (n = 100 pairs) was stratified to include 66 boundary cases, so absolute κ values on batch 1 understate agreement on a natural case mix; batch 2 (n = 100 pairs) is the unbiased held-out validation set that no rule or prompt version touched during development.

| Judge × rule version | Batch 1 (inter-rater κ = 0·688) | Batch 2 (inter-rater κ = 0·812) |
| --- | --- | --- |
| deepseek-v4.1-flash × v2 | 0·540 / 0·440 | — (not run) |
| deepseek-v4.1-flash × v3 | 0·681 / 0·595 † | 0·660 / 0·760 |
| GLM-5.3-flash × v2 | 0·762 / 0·629 | — (not run) |
| GLM-5.3-flash × v3 | 0·786 / 0·612 † | **0·814 / 0·878** |

† Tuning-contaminated: the v3 rules were revised in response to error analysis on batch 1, so batch-1 v3 figures are reported for reference only and were not used to claim validity. The unbiased estimate is batch 2.

The primary and sensitivity judges agreed with each other at κ = 0·80 on batch 2. Across the 72,380 pairs held in both current caches, they disagreed on 4,122 (5·69%): 3,375 NO→YES and 747 YES→NO under GLM relative to deepseek. The v2→v3 revision flipped 10 of the 100 batch-1 pairs, all NO→YES. Dataset and label-granularity breakdowns are given in S8. These are judge–judge consistency measures, not clinician-referenced misclassification rates; the following panels evaluate judge–clinician agreement directly.

**S5-2. Dataset-specific judge validation: the same protocol on MedCaseReasoning and ER-Reason**

The raters are senior clinicians who are not authors of this study (a chief physician of respiratory medicine and an associate chief physician of hematology); the same two raters served all three corpora. The batch-2 protocol was repeated verbatim on the other two corpora, 100 pairs each: stratified 50 YES / 50 NO by the primary judge’s cached verdicts, half of each stratum boundary pairs (judge versus string-heuristic disagreement), items excluded from the CPC batches, both raters blinded to the judge’s verdicts and to each other, same four-clause rubric extended by a worked-examples clarification (the main-diagnosis test, below), diagnoses displayed in Chinese translation with the English original collapsible (translation performed per string, independent of the pairing, by the judge model — translation only, no equivalence judgment; translations_er_mcr.json), and the same blinded web instrument. MCR reference labels are disease-level but often descriptive case-report phrases; 46% of ER-Reason reference labels are symptom-level (168 of 364) (e.g., “Seizure”, “Lethargy”), the rest disease-level.

| Corpus | Inter-rater κ | Judge vs rater A | Judge vs rater B | Majority vs judge |
| --- | --- | --- | --- | --- |
| CPC (batch 2) | 0·812 | 0·814 | 0·878 | 0·772 |
| MedCaseReasoning | 0·633 | 0·620 | 0·460 | 0·520 |
| ER-Reason (primary, post-clarification) | 0·612 | **0·640** | 0·580 | 0·560 |

Two findings follow. First, the equivalence criterion itself degrades with reference-label character: the inter-rater κ falls from 0·812 (CPC disease-level) through 0·633 (MCR descriptive disease-level) to 0·612 (ER-Reason, 46% symptom-level, both raters under the clarified criterion) — and lower still before clarification (first-pass readings below) — so the human ground truth for adjudication is measurably softer on symptom-level labels than on disease-level ones. Second, the primary judge applies the naming criterion that the hit-rate endpoint is defined on (does the candidate name the reference diagnosis), and under that criterion it agrees with both trained raters at or near their mutual level on every corpus: CPC 0·814/0·878 against 0·812; ER-Reason 0·640/0·580 against 0·612; MCR 0·620/0·460 against 0·633.

The criterion clarification is part of the record. On ER-Reason, rater A’s first pass applied a clinical-relevance criterion — candidates that cause or mimic the referenced presentation were marked equivalent (YES rate 78% against the judge’s 50% by construction; κ versus the judge 0·32) — and was voided and re-done after the rubric was extended with the main-diagnosis test (score the candidate’s principal diagnosis, usually the head noun before “causing/secondary to/presenting with”: a candidate whose principal diagnosis is the reference label with an appended aetiology is equivalent; a candidate whose principal diagnosis merely causes or mimics the reference presentation is not). Rater B, annotating independently and without the clarification discussion, reproduced rater A’s pre-clarification pattern (YES rate 69%, κ versus the judge 0·38, κ 0·526 against A’s post-clarification pass): the two readings of symptom-level equivalence are both natural to clinicians, and the first-pass annotations of both raters are retained as documentation of that contest (backups/er_annotations_{a,b}_pass1_*.json). Rater B then re-annotated the same 100 pairs under the clarified rubric, in a reshuffled order, after a wash-out interval; one intervening same-day attempt showed overcorrection (YES rate 27%, κ 0·18 against the rater’s own first pass) and is excluded. The re-annotation (reported above) is self-consistent with the rater’s own first reading (κ = 0·645) while moving to the naming criterion (κ versus the judge 0·580), giving the corpus two post-clarification raters and the 0·612 inter-rater figure of the table. Judge–human disagreement on MCR is concentrated in genuinely contestable descriptive-label pairs (e.g., “starvation-induced liver enzyme elevation” versus “starvation hepatitis”; “diffuse alveolar haemorrhage” versus “pulmonary capillaritis”), with no directional drift. Source: routing_study/results/judge_validity/kappa_report_er_primary.json, kappa_report_{er,mcr}.json, sample_er{,_b2,_b3}.json, and translate_validity_samples.py / build_er_mcr_validity_samples.py; the CPC batch-2 row is kappa_report_glm_v3_batch2.json.

**S5-3. Judge validation on the self-hosted rerun’s candidate distribution: the same protocol and raters on ER-Reason**

Because the self-hosted ER-Reason rerun shifted the candidate distribution, the same two raters repeated the batch-2 protocol on 100 new pairs: 50 YES / 50 NO stratified by the primary judge’s cached verdicts, 61 boundary pairs deliberately over-sampled, and every previously rated pair excluded (rng seed 77). Both raters were blinded to the judge and to each other. Diagnoses were displayed in Chinese translation with the English original available; κ was computed after both raters had finished.

| Corpus | Inter-rater κ | Judge vs rater A | Judge vs rater B | Majority vs judge |
| --- | --- | --- | --- | --- |
| ER-Reason (self-hosted rerun distribution) | **0·877** | 0·580 | 0·540 | 0·580 |

Agreement rates were 94% between raters, 79% for judge–A, 77% for judge–B, and 79% for majority–judge; rater YES rates were 43% and 41% against the judge’s 50% by construction. Judge–clinician κ was below the raters’ mutual κ on this boundary-enriched sample. The second judge and word-coverage analyses (S8, S34) assess consistency and differential misclassification risk; they do not bound clinician-referenced error. This stratified sample also does not support unbiased arm-specific false-accept or false-reject estimates. Source: routing_study/scripts/build_er_local_validity_sample.py and routing_study/results/judge_validity/{sample_er_local.json,erloc_annotations_{a,b}.json,kappa_report_erloc.json}.

### Supplementary Table S6. Workup-informed versus presentation-only input on ER-Reason (n = 364, primary judge, five seeds)

Values are recomputed from routing_study/results/workup_vs_baseline.json, the machine-readable output of routing_study/scripts/eval_workup_vs_baseline.py (the same script that produced the now-supplementary per-seed arm levels formerly shown as main-text Table 2, and the interaction statistics). The two conditions share case identifiers, reference labels, prompts, model, decoding settings, seeds, and judge; only the input differs, the presentation text being reused byte-for-byte and the objective results of that encounter appended under a short heading. Cells in which a candidate–reference pair was left unadjudicated are excluded from the hit rate and reported through the denominator, never scored as misses. The hit rates in S6a are per-seed means whereas S6b reports case-level differences (each case contributing its five-seed hit-rate difference), so the two conventions differ slightly in the last digit on some cells.

**Temperature-matched check.** With all three arms at 0·3 (presentation-only), the single call stays ahead of MDT at every endpoint (top-3 +7·7, p = 2·8 × 10⁻⁸; top-5 +7·6, p = 5·6 × 10⁻⁷) and of P on recall, so the reversal is not a decoding-temperature artefact.

*The main-text granularity-stratified comparisons of the temperature-unmatched arms (n = 168 symptom-level / 196 disease-level) are exploratory and reported in the main text only. After the self-hosted rerun, the symptom-stratum recall contrasts survive Benjamini–Hochberg correction within the six-test family (top-3 −7·4 points, q = 0·049; top-5 −9·6 points, q = 0·022) while the disease-stratum contrasts — directionally concordant at −4·2, −5·6, and −3·6 points — are not individually significant (p = 0·064, 0·072, and 0·36). The archived cloud-era readings (Note S28 of the separate archive file, supplement_archive_separate.md), in which all six contrasts survived, predate the rerun.*

**S6a. Hit rates per seed (%)**

| Strategy | Input | Metric | Seed 1 | Seed 2 | Seed 3 | Seed 4 | Seed 5 | Mean ± SD |
| --- | --- | --- | --- | --- | --- | --- | --- | --- |
| A×1 | Presentation only | Top-1 | 40·7 | 40·7 | 40·7 | 40·7 | 40·7 | 40·7 ± 0·0 |
| A×1 | Presentation only | Top-3 | 54·4 | 54·4 | 54·4 | 54·4 | 54·4 | 54·4 ± 0·0 |
| A×1 | Presentation only | Top-5 | 61·3 | 61·3 | 61·3 | 61·3 | 61·3 | 61·3 ± 0·0 |
| A×1 | Plus objective results | Top-1 | 46·2 | 46·2 | 46·2 | 46·2 | 46·2 | 46·2 ± 0·0 |
| A×1 | Plus objective results | Top-3 | 62·4 | 62·4 | 62·4 | 62·4 | 62·4 | 62·4 ± 0·0 |
| A×1 | Plus objective results | Top-5 | 68·4 | 68·4 | 68·4 | 68·4 | 68·4 | 68·4 ± 0·0 |
| P | Presentation only | Top-1 | 38·7 | 40·7 | 38·7 | 41·5 | 40·1 | 39·9 ± 1·2 |
| P | Presentation only | Top-3 | 51·4 | 48·4 | 50·3 | 51·4 | 49·5 | 50·2 ± 1·3 |
| P | Presentation only | Top-5 | 57·1 | 54·1 | 56·6 | 56·9 | 56·3 | 56·2 ± 1·2 |
| P | Plus objective results | Top-1 | 44·2 | 42·9 | 43·7 | 42·6 | 45·9 | 43·8 ± 1·3 |
| P | Plus objective results | Top-3 | 58·0 | 53·8 | 57·1 | 56·3 | 59·1 | 56·9 ± 2·0 |
| P | Plus objective results | Top-5 | 64·8 | 60·4 | 64·6 | 63·7 | 65·4 | 63·8 ± 2·0 |
| MDT | Presentation only | Top-1 | 38·2 | 36·8 | 36·8 | 35·2 | 36·8 | 36·8 ± 1·1 |
| MDT | Presentation only | Top-3 | 49·7 | 50·0 | 47·8 | 46·4 | 45·9 | 48·0 ± 1·9 |
| MDT | Presentation only | Top-5 | 55·2 | 56·6 | 53·8 | 54·7 | 54·1 | 54·9 ± 1·1 |
| MDT | Plus objective results | Top-1 | 38·7 | 39·8 | 39·8 | 39·0 | 40·4 | 39·6 ± 0·7 |
| MDT | Plus objective results | Top-3 | 52·7 | 51·1 | 50·8 | 51·1 | 51·6 | 51·5 ± 0·8 |
| MDT | Plus objective results | Top-5 | 61·0 | 59·1 | 57·1 | 58·5 | 61·0 | 59·3 ± 1·7 |

All cells use n = 364; every candidate–reference pair in both conditions was adjudicated (no exclusions). The A×1 rows carry SD = 0 because temperature 0 on the self-hosted stack is exact greedy decoding (Methods).

**S6b. Workup effect by strategy and interaction tests (per-case five-seed hit rates)**

d_strategy = mean per-case difference (workup − presentation-only); differences between strategies are paired across cases (paired Wilcoxon two-sided, case-level cluster bootstrap 10,000 resamples). Positive values mean the strategy gained more from the added objective text.

| Endpoint | n | d(A×1) | d(P) | d(MDT) | d_MDT − d_A×1 [95% CI] | p | d_P − d_A×1 [95% CI] | p | d_MDT − d_P [95% CI] | p | Friedman χ² | p |
| --- | --- | --- | --- | --- | --- | --- | --- | --- | --- | --- | --- | --- |
| Top-1 | 364 | +5·5pp | +3·9pp | +2·8pp | −2·7pp [−5·5, +0·1] | 0·0521 | −1·6pp [−4·8, +1·7] | 0·3710 | −1·1pp [−4·0, +1·8] | 0·5554 | 2·200 | 0·3328 |
| Top-3 | 364 | +8·0pp | +6·7pp | +3·5pp | −4·5pp [−8·4, −0·7] | 0·0271 | −1·3pp [−5·4, +2·8] | 0·5982 | −3·2pp [−6·3, −0·1] | 0·0295 | 5·253 | 0·0723 |
| Top-5 | 364 | +7·1pp | +7·6pp | +4·5pp | −2·7pp [−7·0, +1·5] | 0·2493 | +0·4pp [−3·5, +4·5] | 0·6117 | −3·1pp [−6·3, +0·1] | 0·0176 | 5·073 | 0·0791 |

**S6c. Interaction by reference-label granularity (exploratory, uncorrected for multiplicity)**

| Stratum | Endpoint | n | d_MDT − d_A×1 [95% CI] | p | Friedman χ² | p |
| --- | --- | --- | --- | --- | --- | --- |
| Symptom-level label (n = 168) | Top-1 | 168 | −3·0pp [−7·4, +1·3] | 0·3171 | 0·329 | 0·8483 |
| Symptom-level label (n = 168) | Top-3 | 168 | −4·4pp [−10·4, +1·5] | 0·1879 | 2·669 | 0·2633 |
| Symptom-level label (n = 168) | Top-5 | 168 | −1·8pp [−8·5, +4·9] | 0·6285 | 6·938 | 0·0311 |
| Disease-level label (n = 196) | Top-1 | 196 | −2·4pp [−6·1, +1·1] | 0·0688 | 2·353 | 0·3083 |
| Disease-level label (n = 196) | Top-3 | 196 | −4·5pp [−9·4, +0·4] | 0·0409 | 4·861 | 0·0880 |
| Disease-level label (n = 196) | Top-5 | 196 | −3·5pp [−8·9, +1·8] | 0·2261 | 1·252 | 0·5346 |

At top-3 the interaction is negative in both strata (symptom-level −4·4 points, p = 0·19; disease-level −4·5 points, p = 0·041; neither survives correction within the interaction family), and at top-3/top-5 the joint Friedman test across the three strategies’ gains approaches significance — the added objective text helps the single call more than the role-based team, the opposite of what an information-scarcity account requires. In the declared leakage sensitivity analysis, excluding the 20 cases whose reference label was recoverable from the objective text left n = 344; the top-3 MDT-versus-A×1 interaction remained negative (−4·360pp [−8·256, −0·464], p = 0·03218; routing_study/results/local_rerun/extras.json, workup_exclusions).

### Supplementary Table S7. Held-out (46) and development (41) subsets, primary case-level analysis

Recomputed by routing_study/scripts/holdout_primary.py (2026-09-19) and written to routing_study/results/holdout46_primary.{json,md}. The 46 held-out cases were never exposed to prompt tuning or judge refinement; the 41 development cases are the subset used for prompt design and for rule development, and the two subsets are disjoint and sum to the 87 CPC cases of main-text Table 1 (split audited in the script). Statistics follow the main-text case-level framework, reusing the functions of stats_caselevel.py verbatim: per-case five-seed hit rates, paired Wilcoxon signed-rank test (two-sided, cases with a zero difference discarded), and 95% percentile confidence intervals from a case-level cluster bootstrap (10,000 resamples). Judge-cache coverage is complete in every cell: 0 of 3,450 candidate–reference pairs missing on the held-out subset and 0 of 3,075 on the development subset, so no case is excluded, and no candidate list contains fewer than five items.

**S7a. Hit rates (%), primary judge (GLM-5.3-flash × v3)**

| Subset | Strategy | Top-1 | Top-3 | Top-5 |
| --- | --- | --- | --- | --- |
| Held-out (n = 46) | A×1 | 55·7 ± 1·2 | 77·4 ± 4·5 | 82·2 ± 2·8 |
|  | P | 56·1 ± 3·2 | 73·0 ± 2·9 | 78·7 ± 2·4 |
|  | MDT | 60·0 ± 3·3 | 80·4 ± 1·5 | 84·8 ± 1·5 |
| Development (n = 41) | A×1 | 60·5 ± 3·2 | 74·6 ± 5·3 | 80·0 ± 4·0 |
|  | P | 51·2 ± 5·7 | 71·2 ± 4·0 | 74·6 ± 4·1 |
|  | MDT | 61·0 ± 3·9 | 82·9 ± 5·2 | 86·3 ± 4·1 |

Values are mean ± SD across the five seeds; the held-out rows correspond to the percentages reported in the main text. The full 87-case rows of the same computation equal main-text Table 1 exactly.

**S7b. Paired comparisons, primary judge (mean difference [95% CI], Wilcoxon p)**

| Subset | Comparison | Top-1 | Top-3 | Top-5 |
| --- | --- | --- | --- | --- |
| Held-out (n = 46) | MDT vs A×1 | +4·3pp [−1·7, +11·3], p = 0·277 (15/46 non-tied) | +3·0pp [−1·3, +7·4], p = 0·096 (12/46) | +2·6pp [+0·0, +6·1], p = 0·074 (5/46) |
|  | P vs A×1 | +0·4pp [−8·3, +9·1], p = 0·950 | −4·3pp [−13·0, +3·9], p = 0·373 | −3·5pp [−12·2, +4·8], p = 0·453 |
|  | MDT vs P | +3·9pp [−5·2, +13·9], p = 0·400 | +7·4pp [−0·9, +16·5], p = 0·104 | +6·1pp [−0·9, +13·9], p = 0·132 |
| Development (n = 41) | MDT vs A×1 | +0·5pp [−5·4, +7·3], p = 0·964 | +8·3pp [+0·0, +16·6], p = 0·055 | +6·3pp [+0·0, +13·7], p = 0·066 |
|  | P vs A×1 | −9·3pp [−18·0, −2·0], p = 0·076 | −3·4pp [−11·7, +3·4], p = 0·307 | −5·4pp [−12·7, +0·5], p = 0·102 |
|  | MDT vs P | +9·8pp [+1·5, +19·0], p = 0·107 | +11·7pp [+2·9, +21·5], p = 0·0085 | +11·7pp [+3·9, +20·5], p = 0·0086 |

The “non-tied” counts are the cases on which the paired test actually runs, since the test discards cases whose five-seed mean hit rates are equal. No comparison on the held-out subset reaches significance at any endpoint under the primary judge, and the held-out MDT-versus-A×1 margins are about half the development-set margins, consistent with the optimistic bias expected when prompt material is tuned on the development cases.

**S7c. Sensitivity judge (deepseek-v4.1-flash × v3) on the same subsets**

| Subset | Strategy | Top-1 | Top-3 | Top-5 |
| --- | --- | --- | --- | --- |
| Held-out (n = 46) | A×1 | 47·0 ± 1·2 | 72·6 ± 3·9 | 79·1 ± 2·9 |
|  | P | 50·4 ± 2·8 | 70·4 ± 3·3 | 77·8 ± 1·8 |
|  | MDT | 55·7 ± 2·5 | 78·3 ± 0·0 | 83·0 ± 1·0 |
| Development (n = 41) | A×1 | 57·1 ± 2·8 | 72·7 ± 5·0 | 76·1 ± 4·7 |
|  | P | 49·8 ± 5·6 | 68·8 ± 2·7 | 72·2 ± 2·8 |
|  | MDT | 58·5 ± 3·0 | 78·5 ± 3·2 | 81·5 ± 2·8 |

| Subset | Comparison | Top-1 | Top-3 | Top-5 |
| --- | --- | --- | --- | --- |
| Held-out (n = 46) | MDT vs A×1 | +8·7pp [+1·3, +17·0], p = 0·064 | +5·7pp [−0·9, +12·6], p = 0·101 | +3·9pp [−0·9, +10·0], p = 0·091 |
|  | P vs A×1 | +3·5pp [−3·9, +11·3], p = 0·469 | −2·2pp [−10·0, +5·7], p = 0·682 | −1·3pp [−9·1, +6·5], p = 0·916 |
| Development (n = 41) | MDT vs A×1 | +1·5pp [−4·9, +8·8], p = 0·722 | +5·9pp [−2·0, +13·7], p = 0·119 | +5·4pp [−2·0, +12·7], p = 0·065 |

The direction of every held-out and development contrast agrees between the two judges, and under the case-level framework neither judge reaches significance on the held-out subset at any endpoint. The framework matters for the sensitivity judge only: under the legacy pooled case–seed McNemar framework, the held-out MDT-versus-A×1 top-1 contrast was significant for the sensitivity judge (30:10, p = 0·0022; Supplementary Table S2) and not for the primary judge (22:12, p = 0·12), which is why the main text attributes the apparent held-out top-1 advantage to the retired pooled framework rather than to either judge.

### Supplementary Table S8. Judging agreement between the two LLM judges, by dataset and by ER label granularity

Recomputed offline by routing_study/scripts/judge_agreement_local.py (routing_study/results/local_rerun/judge_agreement_by_stratum_local.{json,md}). Agreement is measured on candidate–reference pairs present in both frozen caches (GLM-5.3-flash × v3 and deepseek-v4.1-flash × v3), with keys gold[:150] + "||" + cand[:150] attributed to dataset, strategy, and case from the per-case outputs. All three datasets use five seeds. “GLM YES / DS NO” means the primary judge accepted a pair that the sensitivity judge rejected.

| Stratum | Discordant / total pairs | Discordance rate | GLM YES / DS NO | GLM NO / DS YES |
| --- | --- | --- | --- | --- |
| Intersection of both caches | 4,122 / 72,380 | 5·69% | 3,375 | 747 |
| CPC (five seeds) | 141 / 3,316 | 4·25% | 129 | 12 |
| MedCaseReasoning (five seeds) | 576 / 16,091 | 3·58% | 481 | 95 |
| ER-Reason (five seeds, self-hosted rerun) | 738 / 12,973 | 5·69% | 629 | 109 |
| ER-Reason, symptom-level labels (five seeds) | 329 / 5,837 | 5·64% | 283 | 46 |
| ER-Reason, disease-level labels (five seeds) | 409 / 7,136 | 5·73% | 346 | 63 |

The dataset rows are independent key sets and do not add to the intersection total because a key can recur across seeds or strategies but is counted once per stratum. Denominators use the repaired MCR outputs (S14). This table measures agreement between two LLM judges applying the same rules, not agreement with clinicians and not a clinical misclassification rate. Concordant directions under the second judge reduce concern about a single judge’s idiosyncrasies, but neither judge–judge agreement nor paired scoring eliminates shared or arm-dependent misclassification.

### Supplementary Table S9. Workup-informed input excluding cases with no objective content (n = 330)

Recomputed from routing_study/results/workup_vs_baseline.json, the same machine-readable artifact as Supplementary Table S6, but restricted to the 330 encounters whose objective segment is non-empty. The 34 excluded encounters have an empty objective segment, so nothing is appended to their presentation text and the two conditions are identical by construction; their true gain is exactly zero, which mechanically dilutes every strategy’s mean gain and any interaction between strategies. Gains are per-case differences in five-seed hit rates (workup − presentation-only); interactions are differences between strategies’ per-case gains, paired across cases (paired Wilcoxon two-sided, case-level cluster bootstrap 10,000 resamples, seed 20260917); the Friedman test blocks on case across the three strategies’ gains jointly.

**S9a. Gain from adding the objective results, by strategy (percentage points)**

| Endpoint | n | A×1 | P | MDT |
| --- | --- | --- | --- | --- |
| Top-1 | 330 | +6·1 | +4·2 | +3·1 |
| Top-3 | 330 | +8·8 | +7·5 | +3·9 |
| Top-5 | 330 | +7·9 | +8·2 | +4·4 |

**S9b. MDT-versus-A×1 interaction and joint test**

| Endpoint | n | d_MDT − d_A×1 [95% CI] | p | Friedman p |
| --- | --- | --- | --- | --- |
| Top-1 | 330 | −3·0pp [−6·1, +0·1] | 0·051 | 0·30 |
| Top-3 | 330 | −4·8pp [−9·0, −0·8] | 0·027 | 0·11 |
| Top-5 | 330 | −3·5pp [−8·2, +1·2] | 0·16 | 0·03 |

Every candidate–reference pair was adjudicated, so all cells use n = 330. The same qualitative picture as the full-364 analysis (Supplementary Table S6): all three strategies gain from the added objective text, the ordering is untouched, and the interaction runs against the multi-perspective arms — the single call gains most, the role-based team least — with the top-3 interaction contrast nominal at p = 0·027 (not corrected within the interaction family). Excluding the zero-information cases moves the MDT-versus-A×1 interaction by less than 0·4 points at every endpoint relative to the full-364 estimates, so their diluting effect is negligible.

### Supplementary Note. ER-Reason encounter sampling and input construction

The analysis set was drawn from the 3,984 emergency department encounters of the credentialed ER-Reason corpus [main-text ref 21] in a single scripted step: encounters were deduplicated at the patient level, partitioned by chief complaint, and a stratified random sample of 364 encounters was drawn under a fixed sampling seed, so the sample is reproducible from the source table. Input construction ran on every sampled encounter without exclusions: the emergency department provider note was truncated immediately before the “Medical Decision Making” section, with fallback section markers (including “Final Disposition”) where the primary marker was absent — 3,957 of the 3,984 source encounters carry an explicit primary marker and the remainder were truncated at fallbacks, and no encounter was lost to missing or unparseable text. Reference labels exist at two granularities (symptom-level, n = 168; disease-level, n = 196); this mixture is a property of the source record and was retained as-is. All generation, moderation, and adjudication reported for ER-Reason in this paper used these 364 encounters and no others.

### Supplementary Note. Temperature-matched analyses

Every A×1-versus-multi-perspective comparison in the main text confounds strategy with decoding temperature, because A×1 requested temperature 0 while P and MDT requested 0·3. This note reports the temperature-matched comparisons on all three datasets; every dataset was re-run with all three arms at temperature 0·3 and five seeds each. All analyses are offline and read only the frozen judge cache.

**CPC (n = 87).** All three arms were run at temperature 0·3 for the full 87 cases and all five seeds; A×1 at temperature 0·3 therefore covers the complete set, not a subset of seeds. Statistics follow the main-text case-level framework (per-case five-seed hit rates, paired Wilcoxon two-sided, case-level cluster bootstrap 10,000 resamples). No candidate–reference pair was left unadjudicated in any arm.

| Comparison (temperature 0·3) | Top-1 | Top-3 | Top-5 |
| --- | --- | --- | --- |
| MDT − A×1 | +5·5pp [+0·7, +11·0], p = 0·109 | +6·9pp [+1·8, +12·2], p = 0·010 | +5·1pp [+0·9, +9·9], p = 0·027 |
| MDT − P | +6·7pp [+0·2, +13·3], p = 0·074 | +9·4pp [+3·2, +15·9], p = 0·0043 | +8·7pp [+3·4, +14·7], p = 0·0036 |
| P − A×1 | −1·1pp [−7·8, +5·3], p = 0·778 | −2·5pp [−8·5, +3·2], p = 0·398 | −3·7pp [−9·7, +1·8], p = 0·180 |
| A×1 at 0·3 − A×1 at 0 | −3·0pp [−6·0, −0·2], p = 0·114 | −1·4pp [−3·4, +0·7], p = 0·528 | −0·7pp [−3·9, +2·1], p = 0·916 |

For reference, A×1 at temperature 0·3 reached 54·9 ± 2·2 / 74·7 ± 2·4 / 80·5 ± 1·8% at top-1/3/5, against 57·9 ± 1·9 / 76·1 ± 4·1 / 81·1 ± 2·6% at temperature 0 (Table S1). Two readings follow. The MDT recall advantage survives temperature matching (significant at both recall endpoints, not at top-1), so the main conclusion does not depend on the asymmetry. And the P-versus-A×1 gap, which is not significant at any endpoint in the temperature-matched comparison, is at least partly a temperature effect: A×1 does better at temperature 0, which is the temperature the main analysis gives it, so the main-text CPC contrasts are conservative rather than inflated.

**MedCaseReasoning (n = 406).** The external set was temperature-matched in this revision: A×1 was re-run at temperature 0·3 for all 406 cases and all five seeds (2,030 calls), so the three arms are compared at a common temperature of 0·3 exactly as on CPC. Statistics follow the same case-level framework (per-case five-seed hit rates, paired two-sided Wilcoxon, case-level cluster bootstrap 10,000 resamples, majority-vote exact McNemar), and all 406 cases are adjudicated in every arm.

| Arm | Top-1 | Top-3 | Top-5 |
| --- | --- | --- | --- |
| A×1 (temperature 0·3) | 56·6 ± 0·9 | 69·9 ± 0·7 | 74·9 ± 1·1 |
| P (temperature 0·3) | 56·4 ± 0·3 | 69·2 ± 1·1 | 75·0 ± 1·0 |
| MDT (temperature 0·3) | 55·9 ± 0·8 | 72·9 ± 0·8 | 79·6 ± 0·8 |
| A×1 (temperature 0) | 57·0 ± 0·9 | 69·9 ± 0·6 | 75·6 ± 0·8 |

The P, MDT and temperature-0 A×1 rows are the MedCaseReasoning panel of Supplementary Table S1; the temperature-0.3 A×1 row is new here.

| Comparison | Top-1 | Top-3 | Top-5 |
| --- | --- | --- | --- |
| MDT − A×1 at 0·3 | −0·7pp [−2·9, +1·3], p = 0·4101 (16:17, p = 1·0000) | +3·0pp [+0·8, +5·1], p = 0·0066 (26:10, p = 0·0113) | +4·6pp [+2·8, +6·6], p < 0·0001 (28:8, p = 0·0012) |
| P − A×1 at 0·3 | −0·2pp [−2·6, +2·1], p = 0·9702 (18:23, p = 0·5327) | −0·7pp [−3·2, +1·7], p = 0·8224 (21:26, p = 0·5601) | +0·1pp [−2·2, +2·4], p = 0·7362 (25:24, p = 1·0000) |
| A×1 at 0·3 − A×1 at 0 | −0·4pp [−1·7, +0·9], p = 0·5670 (9:8, p = 1·0000) | +0·0pp [−1·2, +1·2], p = 0·9969 (13:10, p = 0·6776) | −0·7pp [−2·0, +0·6], p = 0·4550 (11:11, p = 1·0000) |

Entries are the first-named arm minus the second, in percentage points, with 95% percentile bootstrap confidence intervals, the paired two-sided Wilcoxon p value, and, in parentheses, majority-vote discordant-pair counts (first-named arm only correct : second only correct) with the exact two-sided McNemar p; every cell has n = 406. Both readings match the CPC panel. **The MDT recall advantage survives temperature matching**: with all three arms at 0·3, MDT remains significantly ahead of the temperature-matched single call at both recall endpoints (+3·0 points, p = 0·0066; +4·6 points, p < 0·0001) and is indistinguishable from it at top-1 (−0·7 points, p = 0·4101). The matched gap is no smaller than under the paper’s original temperature-asymmetric contrast on the same dataset (MDT at 0·3 − A×1 at 0: +3·0 points, p = 0·0079 at top-3 and +3·9 points, p < 0·0001 at top-5), so the asymmetry did not manufacture the advantage; at top-5 it widens from +3·9 to +4·6 points. And **temperature by itself barely moves the single call**: A×1 at 0·3 against A×1 at 0 differs by −0·4, +0·0 and −0·7 points at top-1/3/5, none significant (p ≥ 0·46), which places MedCaseReasoning with ER-Reason rather than with CPC, where the same change costs A×1 3·0, 1·4 and 0·7 points. The **persona arm still gives no gain**: P minus the temperature-matched single call is −0·2, −0·7 and +0·1 points (all p > 0·73), the same null as the unmatched comparison. No candidate–reference pair was left unadjudicated, so the primary figures and the pessimistic (unadjudicated = miss) and optimistic (unadjudicated = hit) bounds coincide. Source: routing_study/results/ax1_t03_mcr.{json,md}, produced by routing_study/scripts/ax1_t03_mcr.py from routing_study/results/topn_seeds_mcr_ax1t03/Ax1t03_s{1..5}.jsonl; the arm differs from the main-experiment A×1 arm in decoding temperature alone, which the script verifies by comparing the two prompts case by case against the source of the main arm’s call sites.

**ER-Reason (n = 364, five seeds).** A×1 at temperature 0·3 covers all five seeds (1,820 case-seed runs), regenerated with the rest of the ER-Reason arms on the self-hosted stack on 2026-10-02..04, so all three arms are compared at a common temperature of 0·3 exactly as on CPC and MedCaseReasoning. Statistics follow the same case-level framework as the rest of this note and of Table 1 (per-case five-seed hit rates, paired two-sided Wilcoxon test, case-level cluster bootstrap 10,000 resamples, majority-vote exact McNemar); all 364 cases are adjudicated in all four arms with 0 undecided candidate–reference pairs, so the primary figures and the pessimistic and optimistic bounds coincide. Source: routing_study/results/local_rerun/ — the temperature-0.3 arm from local_rerun/topn_erreason_ax1t03/Ax1t03_s{1..5}.jsonl, the P, MDT, and temperature-0 A×1 arms from local_rerun/topn_erreason/ — analysed offline against the frozen judge cache; the temperature-0.3 arm differs from the main-experiment A×1 arm in decoding temperature alone.

| Arm | Top-1 | Top-3 | Top-5 |
| --- | --- | --- | --- |
| A×1 (temperature 0·3) | 40·4 ± 0·7 | 55·7 ± 0·9 | 62·5 ± 1·0 |
| P (temperature 0·3) | 39·9 ± 1·2 | 50·2 ± 1·3 | 56·2 ± 1·2 |
| MDT (temperature 0·3) | 36·8 ± 1·1 | 48·0 ± 1·9 | 54·9 ± 1·1 |
| A×1 (temperature 0) | 40·7 ± 0·0 | 54·4 ± 0·0 | 61·3 ± 0·0 |

The P, MDT and temperature-0 A×1 rows are the ER-Reason panel of Supplementary Table S1; the temperature-0.3 A×1 row is new here.

| Comparison | Top-1 | Top-3 | Top-5 |
| --- | --- | --- | --- |
| A×1 at 0·3 − P | +0·5pp [−2·1, +3·1], p = 0·711 (20:19, p = 1·000) | +5·5pp [+2·9, +8·3], p = 4·4 × 10⁻⁵ (31:11, p = 0·0029) | +6·3pp [+3·8, +8·8], p = 1·8 × 10⁻⁶ (35:8, p < 0·0001) |
| A×1 at 0·3 − MDT | +3·7pp [+1·3, +6·0], p = 0·0011 (20:11, p = 0·150) | +7·7pp [+5·1, +10·4], p = 2·8 × 10⁻⁸ (37:12, p = 0·0005) | +7·6pp [+4·8, +10·5], p = 5·6 × 10⁻⁷ (39:13, p = 0·0004) |
| A×1 at 0·3 − A×1 at 0 | −0·2pp [−1·9, +1·5], p = 0·269 (6:9, p = 0·61) | +1·3pp [−0·7, +3·4], p = 0·019 (11:10, p = 1·0) | +1·3pp [−1·2, +3·7], p = 0·054 (16:12, p = 0·57) |

Entries are the first-named arm minus the second, in percentage points, with 95% percentile bootstrap confidence intervals, the paired two-sided Wilcoxon p value, and, in parentheses, majority-vote discordant-pair counts (first-named arm only correct : second only correct) with the exact two-sided McNemar p; every cell has n = 364. Two readings follow. **The matched ordering A×1 > P > MDT is preserved**: with all three arms at 0·3 the single call stays ahead of P at both recall endpoints (+5·5 points at top-3, p = 4·4 × 10⁻⁵; +6·3 at top-5, p = 1·8 × 10⁻⁶; top-1 +0·5, p = 0·71) and ahead of MDT at every endpoint (+3·7, +7·7, and +7·6 points, p = 0·0011, 2·8 × 10⁻⁸, and 5·6 × 10⁻⁷), so neither the emergency reversal nor the persona penalty on this dataset is an artefact of decoding temperature. And **temperature by itself moves A×1 by at most 1·3 points** (−0·2, +1·3, and +1·3 points at top-1/3/5; the top-3 difference is nominally significant, p = 0·019, while every majority-vote test is not), in contrast to CPC, where the same change costs A×1 3·0 points at top-1. Stratified by reference-label granularity the ordering is unchanged in both strata: symptom-level labels (n = 168) give A×1 at 0·3 29·6 / 46·9 / 55·6% against P 29·2 / 39·6 / 48·0% and MDT 26·8 / 39·0 / 46·9%, and disease-level labels (n = 196) give 49·7 / 63·3 / 68·5% against 49·2 / 59·2 / 63·3% and 45·3 / 55·6 / 61·7%, with A×1 − MDT significant at both recall endpoints in both strata (all p ≤ 4·3 × 10⁻⁴) and A×1 − P indistinguishable from it at top-1 in both (p ≥ 0·71).

**Held-out CPC subset.** The same three-arm temperature-0.3 comparison on the 46 held-out cases preserved the direction of the primary analysis: MDT − A×1 was +5·2 points at top-3 (p = 0·073) and +5·2 points at top-5 (p = 0·062), i.e. the same direction as the main analysis and again short of significance, consistent with the limited power of a 46-case set rather than with any inconsistency in direction.

### Supplementary Table S10. Matched control arms on the CPC set (n = 87, five seeds, primary judge)

Values are recomputed from the raw per-case outputs against the primary judge cache (GLM-5.3-flash × v3 rules) by the script named in each panel, and each panel reports 0 missing verdicts, so no case is excluded. Every arm uses qwen3.8-flash, the same 87 cases, five seeds, and the case-level statistics of the main framework (per-case five-seed hit rates, paired two-sided Wilcoxon signed-rank test, case-level cluster bootstrap 10,000 resamples). Differences are given as the first-named arm minus the second, in percentage points, with 95% percentile bootstrap confidence intervals. These are the arms behind Supplementary Figure S3, with panels S10d and S10e carrying the moderator ablation and the completed factorial added in this revision.

**S10a. Reasoning elicitation: the single call asked for a step-by-step justification (A×1+CoT)**

Source: routing_study/results/ax1_cot_cpc.{json,md}, produced by routing_study/scripts/ax1_cot_cpc.py from routing_study/results/topn_seeds_cot/. A×1+CoT and A×1 share model, provider, thinking mode (disabled), requested temperature (0), case text, and call count (one call per case-seed); the only changes are an added request for 5–10 short sentences of step-by-step reasoning before the JSON block and a raised token ceiling (4,096 against 2,048), so that the added justification cannot truncate the ranked list. A×1+CoT therefore isolates the effect of eliciting reasoning within a single call, and its contrast with P and MDT is the structural effect that remains once elicitation is accounted for.

| Arm | Top-1 | Top-3 | Top-5 |
| --- | --- | --- | --- |
| A×1+CoT (T = 0) | 58·9 ± 2·1 | 75·6 ± 2·9 | 82·3 ± 2·9 |
| A×1 (T = 0) | 57·9 ± 1·9 | 76·1 ± 4·1 | 81·1 ± 2·6 |
| P (T = 0·3) | 53·8 ± 4·0 | 72·2 ± 2·6 | 76·8 ± 2·1 |
| MDT (T = 0·3) | 60·5 ± 3·1 | 81·6 ± 2·4 | 85·5 ± 2·1 |

| Comparison | Top-1 | Top-3 | Top-5 |
| --- | --- | --- | --- |
| A×1+CoT − A×1 | +0·9pp [−5·7, +7·4], p = 0·71 | −0·5pp [−6·0, +5·1], p = 0·95 | +1·1pp [−3·0, +5·5], p = 0·61 |
| A×1+CoT − P | +5·1pp [+0·5, +9·9], p = 0·054 | +3·4pp [−1·1, +8·1], p = 0·17 | +5·5pp [+1·1, +10·3], p = 0·020 |
| A×1+CoT − MDT | −1·6pp [−8·7, +5·3], p = 0·59 | −6·0pp [−11·3, −0·9], p = 0·027 | −3·2pp [−6·9, +0·5], p = 0·067 |

Eliciting reasoning moved no endpoint beyond noise against A×1 itself, so the presence of intermediate reasoning text does not account for the A×1-versus-P gap; against the same arm, MDT retains a significant top-3 advantage and a directional top-5 advantage. The reasoning trace was produced in every call of the arm: across the 435 calls, the justification averaged 2,713 characters (minimum 982, no call with an empty justification) and the arm averaged 3,499 total tokens per call, against 2,846 for A×1 over the same cases and seeds.

**S10b. Call-budget control: five independent samples per case (A×5)**

Source: routing_study/results/ax5_full87.{json,md}, produced by routing_study/scripts/ax5_full87.py from routing_study/results/topn_seeds_ax5/ (87 cases × 5 seeds × 5 samples at temperature 0·7 = 2,175 calls). Two aggregations are reported. *Borda* merges the five ranked lists by rank position (weight 1·0 at rank 1 down to 0·2 at rank 5, candidates of the same disease merged, reusing aggregate_top5); *self-consistency* (SC) takes its top-1 from a majority vote over the rank-1 disease cluster and fills the remaining positions in Borda order. Each aggregation sees the same five samples per case, against MDT’s six calls per case (five isolated role calls plus the moderator).

| Arm | Top-1 | Top-3 | Top-5 |
| --- | --- | --- | --- |
| A×5, Borda | 51·3 ± 2·5 | 75·4 ± 1·0 | 81·8 ± 2·1 |
| A×5, self-consistency | 55·4 ± 1·3 | 75·4 ± 1·7 | 81·6 ± 2·4 |
| A×1 (T = 0), reference | 57·9 ± 1·9 | 76·1 ± 4·1 | 81·1 ± 2·6 |
| MDT (T = 0·3), reference | 60·5 ± 3·1 | 81·6 ± 2·4 | 85·5 ± 2·1 |

| Comparison | Top-1 | Top-3 | Top-5 |
| --- | --- | --- | --- |
| A×5 (Borda) − A×1 | −6·7pp [−12·2, −1·1], p = 0·017 | −0·7pp [−3·9, +2·3], p = 0·82 | +0·7pp [−2·5, +3·9], p = 0·59 |
| A×5 (self-consistency) − A×1 | −2·5pp [−6·2, +1·1], p = 0·30 | −0·7pp [−4·1, +2·5], p = 0·89 | +0·5pp [−2·8, +3·7], p = 0·65 |
| A×5 (Borda) − MDT | −9·2pp [−14·5, −4·4], p = 0·0029 | −6·2pp [−11·0, −1·8], p = 0·011 | −3·7pp [−7·4, −0·0], p = 0·059 |
| A×5 (self-consistency) − MDT | −5·1pp [−9·7, −0·9], p = 0·035 | −6·2pp [−11·0, −1·8], p = 0·0095 | −3·9pp [−7·6, −0·2], p = 0·045 |

Spending five calls per case does not recover the team’s recall: neither aggregation beats A×1 at top-3 or top-5 (Borda is 6·7 points *below* A×1 at top-1, p = 0·017), and both are beaten by MDT at every recall endpoint, significantly so at top-3 and top-5 under self-consistency. Call count alone therefore does not explain the MDT advantage.

**S10c. Prompt-transcription control: the P arm with the doubled braces folded (Pfixed)**

Source: routing_study/results/p_fixed_cpc.{json,md}, produced by routing_study/scripts/p_fixed_cpc.py from routing_study/results/topn_seeds_pfixed/. Pfixed is the main P arm with exactly one change, made in a process-local copy of the prompt constant: every {{/}} of the output-format suffix is folded to {/} (the shared constant is untouched, so the main-experiment P arm and every number reported from it are unchanged). Prompt concatenation, the perspective prompt, the case text, model, provider, thinking mode, requested temperature (0·3), token ceiling, and case/seed coverage are otherwise byte-identical. The “only difference” claim is audited case by case rather than asserted: for all 87 cases the two complete prompts have the same number of lines, differ on exactly one line, and differ there only by brace folding (87/87 cases consistent, 0 anomalies; the per-case character difference is −12, the six {{/}} pairs of that line). The defect itself and its effect on the second model family are documented in the Supplementary Note on the prompt-transcription defect (S13).

| Arm | Top-1 | Top-3 | Top-5 |
| --- | --- | --- | --- |
| Pfixed (braces folded) | 52·4 ± 1·3 | 75·6 ± 3·0 | 80·9 ± 1·9 |
| P (main experiment, literal braces) | 53·8 ± 4·0 | 72·2 ± 2·6 | 76·8 ± 2·1 |
| A×1 (T = 0), reference | 57·9 ± 1·9 | 76·1 ± 4·1 | 81·1 ± 2·6 |
| MDT (T = 0·3), reference | 60·5 ± 3·1 | 81·6 ± 2·4 | 85·5 ± 2·1 |

| Comparison | Top-1 | Top-3 | Top-5 |
| --- | --- | --- | --- |
| Pfixed − P | −1·4pp [−4·6, +1·6], p = 0·76 | +3·4pp [+0·2, +6·7], p = 0·052 | +4·1pp [+1·1, +7·4], p = 0·014 |
| Pfixed − A×1 | −5·5pp [−11·5, +0·5], p = 0·094 | −0·5pp [−5·5, +4·6], p = 0·99 | −0·2pp [−4·6, +4·1], p = 0·76 |
| Pfixed − MDT | −8·0pp [−14·9, −1·4], p = 0·045 | −6·0pp [−11·7, −0·5], p = 0·060 | −4·6pp [−9·4, −0·0], p = 0·068 |

Correcting the transcription defect raises the P arm’s recall by 3–4 points and does not lower its top-1. Persona still does not beat the single call (−5·5 points at top-1, p = 0·094; −0·5 and −0·2 at top-3 and top-5), and the team’s recall advantage over persona shrinks to −6·0 and −4·6 points with the corrected prompt, no longer individually significant at either recall endpoint although the point estimates still favour the team. Every P-arm number in the main text and in Table 1 comes from the original, uncorrected prompt; Pfixed appears only as this control.

**S10d. Moderator ablation: the five role lists aggregated by Borda without the moderator (MDT-Borda)**

Source: routing_study/results/mdt_nomoderator.{json,md}, produced by routing_study/scripts/mdt_nomoderator_cpc.py. The arm re-aggregates the five role lists of every MDT case-seed — the role-level outputs already deposited with the MDT run files (topn_mdt/roles.jsonl, seed 1; topn_mdt/s{2..5}/roles.jsonl) — by the same Borda rule used for A×5 (topn_cpc.aggregate_top5, rank-1 weight 1·0 → rank-5 0·2, same_disease cluster merging), with no moderator call; it therefore requires no new model calls and differs from MDT only in how the five opinions are combined. All 435 role files were complete (5 roles × 87 cases × 5 seeds, no empty lists); 0 candidate–reference pairs were left unadjudicated (344 missing pairs were adjudicated and merged into the shared primary cache, 69,283 → 69,627).

| Arm | Top-1 | Top-3 | Top-5 |
| --- | --- | --- | --- |
| MDT-Borda (roles, mechanical aggregation, no moderator) | 52·4 ± 2·9 | 76·3 ± 2·1 | 82·8 ± 1·6 |
| MDT (5 roles + moderator), reference | 60·5 ± 3·1 | 81·6 ± 2·4 | 85·5 ± 2·1 |
| A×1 (T = 0), reference | 57·9 ± 1·9 | 76·1 ± 4·1 | 81·1 ± 2·6 |

| Comparison | Top-1 | Top-3 | Top-5 |
| --- | --- | --- | --- |
| MDT-Borda − MDT | −8·0pp [−13·3, −2·8], p = 0·0044 (2:12, p = 0·0129) | −5·3pp [−9·0, −2·3], p = 0·0030 (0:6, p = 0·0312) | −2·8pp [−5·7, −0·2], p = 0·0557 (2:4, p = 0·6875) |
| MDT-Borda − A×1 | −5·5pp [−12·2, +1·1], p = 0·0860 (5:10, p = 0·3018) | +0·2pp [−4·4, +4·6], p = 0·7642 (3:4, p = 1·0000) | +1·6pp [−1·6, +4·8], p = 0·2810 (3:3, p = 1·0000) |

Mechanical aggregation of the isolated role lists therefore sits at the level of the single call at every endpoint and below the full team at every endpoint, so the moderator’s synthesis — not the aggregated lists alone — carries the team’s advantage. The moderator changed the rank-1 entry in 24–33 of 87 cases per seed relative to the Borda list and reproduced its full top-5 ordering in only 2–4 of 87. On the 46 held-out cases the same ablation gives MDT-Borda 48·3 ± 3·6 / 76·5 ± 3·6 / 83·0 ± 2·4% against the full team’s 60·0 ± 3·3 / 80·4 ± 1·5 / 84·8 ± 1·5%: −11·7pp at top-1 [−19·6, −3·9], p = 0·0135 (majority-vote 1:10, p = 0·0117), so the held-out data, which prompt development cannot have flattered, agree with the full-set reading.

**S10e. Completed factorial: the moderator over plain repeated samples (Ax5Mod)**

Source: routing_study/results/ax5_mod_cpc.{json,md}, produced by routing_study/scripts/ax5_mod_cpc.py. The arm feeds the same moderator with the five plain samples of each A×5 case-seed — the archived samples of topn_seeds_ax5/, generated independently at temperature 0·7 — instead of the five role opinions. The moderator prompt is verbatim that of mdt_cpc.py except its first sentence (“Five specialists independently reviewed the MGH CPC case below, each through their own lens, without seeing each other’s opinions.” → “Five independent assessments of the MGH CPC case below were produced without seeing one another.”), and the panel-opinion block carries no per-candidate rationale because the archived samples have none; both differences are disclosed here and in main-text Methods. Inference is 435 moderator calls on qwen3.8-flash (temperature 0·3, max_tokens 4,096, thinking disabled — the MDT moderator’s settings); 585 new candidate–reference pairs were adjudicated into the shared cache (70,892 → 71,477, 0 unresolved), and the case-level framework is that of the main analysis.

| Arm | Top-1 | Top-3 | Top-5 |
| --- | --- | --- | --- |
| A×5+Mod (samples + moderator) | 58·9 ± 2·2 | 81·4 ± 1·1 | 84·1 ± 1·0 |
| A×5 Borda (same samples, mechanical) | 51·3 ± 2·5 | 75·4 ± 1·0 | 81·8 ± 2·1 |
| MDT (roles + moderator), reference | 60·5 ± 3·1 | 81·6 ± 2·4 | 85·5 ± 2·1 |
| A×1 (T = 0), reference | 57·9 ± 1·9 | 76·1 ± 4·1 | 81·1 ± 2·6 |

| Comparison | Top-1 | Top-3 | Top-5 |
| --- | --- | --- | --- |
| A×5+Mod − A×1 | +0·9pp [−3·9, +6·0], p = 0·668 | +5·3pp [+1·8, +9·4], p = 0·0064 | +3·0pp [−0·5, +7·1], p = 0·091 |
| A×5+Mod − A×5 Borda | +7·6pp [+1·8, +13·3], p = 0·034 | +6·0pp [+2·5, +10·1], p = 0·0036 | +2·3pp [−0·7, +5·7], p = 0·095 |
| A×5+Mod − MDT | −1·6pp [−6·7, +3·4], p = 0·537 | −0·2pp [−3·7, +3·4], p = 0·758 | −1·4pp [−4·8, +2·1], p = 0·374 |

The completed factorial therefore reads: mechanical aggregation leaves both list sources at or below the single-call level (A×5 Borda here; MDT-Borda in S10d), while moderated synthesis lifts both to the team’s level and the two moderated arms are statistically indistinguishable from each other at every endpoint (p ≥ 0·37). The specialist framing of the lists adds nothing measurable to recall on this corpus; what is necessary is independent generation of the lists together with moderated synthesis. A test–retest control quantifies the moderator’s own sampling noise: five moderator calls on the identical Ax5Mod seed-1 input of each case (435 calls) produce worded-different lists (full top-5 identical in 12/87 cases; pairwise top-1 agreement 83·2%; mean pairwise Jaccard 0·67) but identical adjudicated accuracy — per-run top-1 hit rate 62·1% in all five runs, range 0·0 points — so moderator noise cannot account for any between-arm difference. Source: routing_study/results/moderator_retest.{json,md} (routing_study/scripts/moderator_retest.py); 162 new candidate–reference pairs were adjudicated into the shared cache (71,477 → 71,639, 0 unresolved). On the 46 held-out cases the direction matches (A×5+Mod 54·8/81·3/84·8% against A×1 55·7/77·4/82·2% and the team 60·0/80·4/84·8%: top-3 +3·9pp over A×1, p = 0·120; top-5 equal to the team). The emergency counterpart of this arm is panel S10f.

**S10f. The completed factorial on the emergency corpus: the moderator over plain samples does not reverse (ER Ax5Mod)**

The ER-Reason counterpart of panel S10e was regenerated together with the main experiment on the self-hosted stack on 2026-10-02..04 (364 encounters × 5 seeds): fresh sampling of the single-call prompt at temperature 0·7 (five samples per case-seed; 9,100 calls) followed by the identical moderator over the five samples (temperature 0·3, max_tokens 4,096, thinking disabled; prompt byte-identical to S10e’s; 1,820 calls). Generation was driven by the model-box pipeline run_all.sh; the analysis is the offline PHASE=analyze recomputation of routing_study/scripts/ax5_mod_er.py over routing_study/results/local_rerun/topn_ax5_mod_er/ and routing_study/results/local_rerun/topn_erreason/seed_summary.json, scored against the frozen primary judge cache with 0 unresolved pairs, under the case-level framework of the main analysis. The pre-rerun cloud outputs are retained as historical artifacts and are no longer the source of any number in this panel.

| Arm | top-1 | top-3 | top-5 |
| --- | --- | --- | --- |
| A×5+Mod (samples + moderator) | 39·8 ± 1·1 | 54·8 ± 1·7 | 61·9 ± 2·3 |
| A×5 Borda (samples + mechanical aggregation) | 30·2 ± 2·6 | 52·9 ± 1·5 | 61·0 ± 1·0 |
| A×1 (T = 0) | 40·7 ± 0·0 | 54·4 ± 0·0 | 61·3 ± 0·0 |
| P (T = 0·3) | 39·9 ± 1·2 | 50·2 ± 1·3 | 56·2 ± 1·2 |
| MDT (five roles + moderator) | 36·8 ± 1·1 | 48·0 ± 1·9 | 54·9 ± 1·1 |

| Contrast | top-k | mean difference [95% CI] | Wilcoxon p | majority-vote McNemar (a:b) p |
| --- | --- | --- | --- | --- |
| A×5+Mod vs A×1 | top-1 | −0·8pp [−3·3, +1·6] | 0·829 | 11:19 p=0·20 |
| A×5+Mod vs A×1 | top-3 | +0·4pp [−2·3, +3·2] | 0·422 | 19:17 p=0·87 |
| A×5+Mod vs A×1 | top-5 | +0·7pp [−2·3, +3·6] | 0·192 | 21:16 p=0·51 |
| A×5+Mod vs MDT | top-1 | +3·1pp [+0·7, +5·4] | 0·020* | 19:15 p=0·61 |
| A×5+Mod vs MDT | top-3 | +6·8pp [+4·3, +9·4] | 1·2 × 10⁻⁶* | 36:10 p=0·0002* |
| A×5+Mod vs MDT | top-5 | +7·0pp [+4·6, +9·6] | 5·2 × 10⁻⁷* | 35:8 p<0·0001* |
| A×5+Mod vs A×5 Borda | top-1 | +9·7pp [+7·0, +12·4] | 6·9 × 10⁻⁹* | 41:10 p<0·0001* |
| A×5+Mod vs A×5 Borda | top-3 | +1·9pp [−0·2, +4·1] | 0·271 | 21:15 p=0·41 |
| A×5+Mod vs A×5 Borda | top-5 | +0·9pp [−1·3, +3·1] | 0·388 | 18:15 p=0·73 |
| A×5 Borda vs A×1 | top-1 | −10·5pp [−13·8, −7·3] | 2·0 × 10⁻⁷* | 8:47 p<0·0001* |
| A×5 Borda vs A×1 | top-3 | −1·5pp [−4·4, +1·3] | 0·958 | 16:20 p=0·62 |
| A×5 Borda vs A×1 | top-5 | −0·2pp [−3·4, +2·9] | 0·48 | 22:20 p=0·88 |

The moderator-over-mechanical contrast therefore replicates its CPC direction on the emergency corpus at larger magnitude (+9·7pp at top-1 here versus +7·6pp in S10e), while the moderator arm is indistinguishable from the single call at every endpoint (|difference| ≤ 0·8pp, all p ≥ 0·19): plain samples under moderated synthesis neither gain nor lose against A×1 on this distribution, so the emergency reversal is specific to the specialist role lists and not to multi-call inference or to the moderator.

**S10f-2. The same factorial on the second model family (deepseek-v4.1-flash, ER Ax5Mod)**

*Withdrawn.* The three factorial arms of this panel (deepseek-v4.1-flash, ER Ax5Mod) were generated on 2026-09-23 through the provider’s direct API, predating the zero-data-retention routing adopted for the study’s other ER-Reason case-text pathways; following the routing audit of 2026-10-02 the panel is withdrawn and is no longer cited as evidence. The second model family’s emergency replication is carried by the main-text three-arm rerun, whose case text goes through OpenRouter’s zero-retention fixed endpoint with per-request provider auditing (see Methods, “Models and decoding”); the replication’s A×1 baseline, which like this panel predated the routing (generated 2026-09-20 over the direct API), was regenerated through the identical endpoint on 2026-10-05 and its pairs re-adjudicated (S11c), so every tabulated arm of the emergency replication now rests on zero-retention-routed generation. The panel’s former numbers appear in no table of this supplement; the panel number is retained so that cross-references remain stable.

**S10g. P-split: the persona failure located at generation time (fifth factorial cell)**

P-split keeps P’s sequential, mutually conditioning generation and replaces only its adjudication with a fresh moderator call, separating two candidate explanations of the P result: (a) generation-time conditioning collapses the candidate pool, or (b) the non-independent synthesizer discards material that independent adjudication would keep. Generation re-runs the five-perspective stage of P with the perspective prompt truncated immediately before its synthesis task plus an explicit no-synthesis instruction (435 calls); the moderator prompt is byte-identical to S10e’s except its first sentence, which truthfully discloses that the five analyses were written sequentially in one context with later perspectives seeing earlier ones (435 calls). Same model, temperature 0·3, max_tokens 4,096, thinking disabled (routing_study/scripts/p_split_cpc.py). One candidate–reference pair remained unresolved after judging and is counted as a miss.

| Arm | top-1 | top-3 | top-5 |
| --- | --- | --- | --- |
| P-split (conditioned perspectives + independent moderator) | 51·0 ± 2·4 | 75·2 ± 4·3 | 83·2 ± 1·9 |
| P (conditioned perspectives + self-synthesis) | 53·8 ± 4·0 | 72·2 ± 2·6 | 76·8 ± 2·1 |
| A×1 (T = 0) | 57·9 ± 1·9 | 76·1 ± 4·1 | 81·1 ± 2·6 |
| MDT (isolated roles + moderator) | 60·5 ± 3·1 | 81·6 ± 2·4 | 85·5 ± 2·1 |
| A×5+Mod (samples + moderator) | 58·9 ± 2·2 | 81·4 ± 1·1 | 84·1 ± 1·0 |

| Contrast | top-k | mean difference [95% CI] | Wilcoxon p | majority-vote McNemar (a:b) p |
| --- | --- | --- | --- | --- |
| P-split vs P | top-1 | −2·8pp [−7·7, +1·9] | 0·2743 | 3:6 p=0·5078 |
| P-split vs P | top-3 | +3·0pp [−1·8, +8·0] | 0·2140 | 7:5 p=0·7744 |
| P-split vs P | top-5 | +6·4pp [+2·3, +11·0] | 0·0034* | 6:3 p=0·5078 |
| P-split vs A×1 | top-1 | −6·5pp [−13·0, −0·2] | 0·0416* | 3:8 p=0·2266 |
| P-split vs MDT | top-1 | −9·5pp [−16·7, −2·6] | 0·0128* | 4:14 p=0·0309* |
| P-split vs A×5+Mod | top-1 | −7·9pp [−14·0, −2·1] | 0·0107* | 2:9 p=0·0654 |

P-split sits with P, not with the moderator arms, at top-1 while its top-5 exceeds P by 6·4 points: an independent adjudicator recovers tail material that P’s self-synthesis dropped, but cannot restore diversity that was never generated. The persona failure on this corpus is therefore a generation-time failure, and independence of generation is an input-level requirement. Source: routing_study/results/p_split_cpc.{json,md}.

**S10h. Moderator anatomy: input ablation, opinion-count sweep, and order permutation (seed-1 panel; five-seed powered replication on CPC and MedCaseReasoning)**

Three single-seed ablations take the Ax5Mod seed-1 inputs (87 cases) and vary one property of the moderator’s input at a time (routing_study/scripts/moderator_anatomy.py); each is paired against the unmodified Ax5Mod seed-1 outputs. The lists-only arm removes the case text and replaces the guideline “resolve conflicts by re-checking against the case text” with “resolve conflicts by weighing the evidence across the lists”, prompt otherwise verbatim. The k-sweep arms truncate the opinion list to its first k entries, k ∈ {1,2,3,4}, with the prompt’s count word adjusted truthfully. The order arms present the opinions in a fixed random permutation (seed 20260920) and in full reversal. Because each arm is a single moderator run on one seed, the confidence intervals (±6–7 points at top-1) bound only large effects, and these comparisons are exploratory.

The same three ablations were then run under identical prompt variants, decoding, and frozen judge at five seeds on both the CPC (n = 87) and MedCaseReasoning (n = 406) corpora (routing_study/scripts/moderator_anatomy_full.py; deposited as routing_study/results/moderator_anatomy_full.{json,md}), replacing the single-seed caveat with the following readings. **Input ablation**: removing the case text left the recall endpoints statistically unchanged on both corpora (CPC top-3 −2·3 points, p = 0·11, top-5 −0·0, p = 1·0; MedCaseReasoning top-3 −0·4, p = 0·27, top-5 −1·1, p = 0·074) while costing a small, significant top-1 decrement on the larger corpus (MedCaseReasoning −1·8 points, p = 0·0006; CPC −2·1, p = 0·12) — the moderator’s recall synthesis is list-driven, and the case text contributes a small head-of-list benefit rather than the recall gain. **Opinion-count sweep**: a one-opinion floor appears at the breadth endpoint on MedCaseReasoning (k = 1 top-5 −3·6 points, p < 0·0001, recovering monotonically to within noise by k = 3; k = 2 −1·7, p = 0·032), while top-3 recall shows no dose-response pattern on MedCaseReasoning (k = 2–4 all |effects| ≤ 0·7 points, all p ≥ 0·13) and the CPC k-sweep retains its seed-1 scatter (k = 1 −4·8, p = 0·026; k = 4 −2·3, p = 0·040; k = 2 and k = 3 null) rather than any monotone trend; plurality therefore matters only at the list’s breadth and only below two-to-three lists, refining — not reversing — the seed-1 “no dose-response” reading. **Order**: a fixed permutation changed nothing at any endpoint on either corpus (all p ≥ 0·16), and a full reversal remained within noise at both recall endpoints on both corpora (largest |effect| 0·7 points, all p ≥ 0·072), the lone nominal effect being a CPC top-1 advantage under reversal (+2·8 points, p = 0·015) that does not survive correction within the ablation family. At five seeds the moderator is therefore an order-invariant synthesis whose recall output needs at least two independent lists but no particular subset of them.

| Arm | top-1 | top-3 | top-5 |
| --- | --- | --- | --- |
| Ax5Mod_s1_ref (full input, k = 5) | 62·1 | 82·8 | 85·1 |
| lists_only (no case text) | 60·9 | 80·5 | 83·9 |
| k=1 | 62·1 | 79·3 | 83·9 |
| k=2 | 59·8 | 79·3 | 82·8 |
| k=3 | 57·5 | 81·6 | 83·9 |
| k=4 | 60·9 | 82·8 | 85·1 |
| order_reverse (reversed order) | 63·2 | 81·6 | 85·1 |
| order_perm20260920 (fixed random order) | 62·1 | 82·8 | 86·2 |

No ablation reaches significance against the reference at any endpoint except the isolated k=3 top-1 dip (−4·6pp, p = 0·046), which has no monotone pattern around it and is read as noise. The moderator’s adjudicated output is invariant to removing the case text, to reducing five opinions to one (top-1 identical at 62·1%), and to permuting presentation order: it behaves as a synthesis function over the lists themselves rather than as a re-reader of the case or an aggregator of volume. Source: routing_study/results/moderator_anatomy.{json,md}.

**S10i. The completed factorial on MedCaseReasoning: the moderator-over-mechanical contrast at its largest (MCR Ax5Mod)**

The external counterpart of panels S10e/S10f repeats the full factorial recipe on MedCaseReasoning: fresh sampling of the single-call prompt at temperature 0·7 (five samples per case-seed; 10,150 calls) followed by the identical moderator (temperature 0·3, max_tokens 4,096, thinking disabled; prompt byte-identical to S10e’s; 2,030 calls), five seeds, n = 406 cases (routing_study/scripts/ax5_mod_mcr.py). Judging crossed a multi-hour provider degradation on 2026-09-21; a bounded-batch retry loop (250-pair batches, per-call throttling, hard request deadlines, and an exclusive file lock on the shared cache) drove unresolved pairs from 3,745 to zero before the analysis was produced, superseding an interim analysis computed on the incomplete adjudication, which is retained only as a voided artifact.

| Arm | top-1 | top-3 | top-5 |
| --- | --- | --- | --- |
| A×5+Mod | 58·2 ± 0·9 | 71·5 ± 0·8 | 79·0 ± 0·8 |
| A×5 Borda | 47·5 ± 2·1 | 69·3 ± 1·2 | 75·6 ± 1·3 |
| A×1 | 57·0 ± 0·9 | 69·9 ± 0·6 | 75·6 ± 0·8 |
| P | 56·4 ± 0·3 | 69·2 ± 1·1 | 75·0 ± 1·0 |
| MDT | 55·9 ± 0·8 | 72·9 ± 0·8 | 79·6 ± 0·8 |

| Contrast | top-k | mean difference [95% CI] | Wilcoxon p |
| --- | --- | --- | --- |
| A×5+Mod vs A×1 | top-1 | +1·1pp [−0·2, +2·6] | 0·0871 |
| A×5+Mod vs A×1 | top-3 | +1·6pp [+0·2, +3·0] | 0·0183* |
| A×5+Mod vs A×1 | top-5 | +3·4pp [+1·6, +5·2] | <0·0001* |
| A×5+Mod vs MDT | top-1 | +2·3pp [+0·3, +4·3] | 0·0166* |
| A×5+Mod vs MDT | top-3 | −1·4pp [−3·4, +0·6] | 0·2241 |
| A×5+Mod vs MDT | top-5 | −0·5pp [−2·3, +1·3] | 0·5165 |
| A×5+Mod vs P | top-1 | +1·8pp [−0·4, +4·0] | 0·2152 |
| A×5+Mod vs P | top-3 | +2·3pp [+0·0, +4·6] | 0·0941 |
| A×5+Mod vs P | top-5 | +4·0pp [+1·7, +6·4] | 0·0023* |
| A×5+Mod vs A×5 Borda | top-1 | +10·6pp [+8·1, +13·2] | <0·0001* |
| A×5+Mod vs A×5 Borda | top-3 | +2·2pp [+0·7, +3·6] | 0·0031* |
| A×5+Mod vs A×5 Borda | top-5 | +3·4pp [+1·9, +5·0] | 0·0002* |

The factorial geometry replicates on the external corpus with the moderator-over-mechanical contrast at its largest of the three corpora: +10·6pp top-1 here versus +9·7pp on ER-Reason (S10f) and +7·6pp on CPC (S10e). Mechanical aggregation again sits below the single call at top-1 (A×5 Borda 47·5% versus A×1 57·0%), the moderator arm matches the role-based team at both recall endpoints (top-5 −0·5pp, p = 0·52) while exceeding it at top-1 (+2·3pp, p = 0·017), and the recall advantage over A×1 holds for the moderator arm (+1·6pp top-3, p = 0·018; +3·4pp top-5, p < 0·0001). Source: routing_study/results/ax5_mod_mcr.{json,md}.

### Supplementary Table S11. Second model family (deepseek-v4.1-flash) on the CPC and ER-Reason sets (five seeds, primary judge)

Source: routing_study/results/dsflash_family_cpc.{json,md}. A×1 and P were run by routing_study/scripts/seeds_87_dsflash.py; MDT ran the existing isolated-role implementation (routing_study/scripts/mdt_cpc.py) on the same provider. The outputs are routing_study/results/topn_seeds_dsflash/ (A×1, P) and routing_study/results/topn_mdt_dsflash/ (roles, synthesis). Provider deepseek-v4.1-flash, model id deepseek-v4.1-flash, endpoint https://api.deepseek.com/v1; A×1 requested temperature 0, P and MDT 0·3; thinking disabled; max_tokens 2,048 for A×1 and 4,096 for P and the MDT role and moderator calls. Scoring is one call per case-seed for A×1 and P (870 calls over five seeds) and six for MDT. Prompts and case text are byte-identical to the main experiment (A_TOPN_PROMPT; PERSPECTIVE_PROMPT + P_TOPN_SUFFIX; the role and moderator prompts of S-M3), which is what makes the two families comparable. The qwen rows are the main qwen3.8-flash arms recomputed over the same five seeds (Table 1).

| Arm | Top-1 | Top-3 | Top-5 |
| --- | --- | --- | --- |
| deepseek-v4.1-flash A×1 | 46·7 ± 0·6 | 72·9 ± 1·0 | 79·5 ± 0·5 |
| deepseek-v4.1-flash P | 47·4 ± 1·7 | 66·9 ± 3·2 | 75·6 ± 3·3 |
| deepseek-v4.1-flash MDT | 51·5 ± 1·3 | 75·9 ± 0·8 | 83·2 ± 2·9 |
| qwen3.8-flash A×1 (five seeds) | 57·9 ± 1·9 | 76·1 ± 4·1 | 81·1 ± 2·6 |
| qwen3.8-flash P (five seeds) | 53·8 ± 4·0 | 72·2 ± 2·6 | 76·8 ± 2·1 |
| qwen3.8-flash MDT (five seeds) | 60·5 ± 3·1 | 81·6 ± 2·4 | 85·5 ± 2·1 |

**S11a. Paired comparisons within the second family (deepseek-v4.1-flash)**

| Comparison | Top-1 | Top-3 | Top-5 |
| --- | --- | --- | --- |
| MDT − A×1 | +4·8pp [+0·2, +9·7], p = 0·067 | +3·0pp [−0·7, +6·7], p = 0·171 | +3·7pp [−0·0, +7·6], p = 0·115 |
| MDT − P | +4·1pp [−3·0, +11·3], p = 0·359 | +9·0pp [+1·6, +16·6], p = 0·017 | +7·6pp [+1·8, +13·3], p = 0·023 |
| P − A×1 | +0·7pp [−7·4, +8·5], p = 0·673 | −6·0pp [−13·1, +1·1], p = 0·096 | −3·9pp [−9·4, +1·6], p = 0·327 |

**S11b. Cross-family contrast on the same cases and seeds (qwen3.8-flash minus deepseek-v4.1-flash)**

| Comparison | Top-1 | Top-3 | Top-5 |
| --- | --- | --- | --- |
| MDT, qwen − deepseek-v4.1-flash | +9·0pp [−0·2, +18·6], p = 0·044 | +5·7pp [−0·7, +12·4], p = 0·091 | +2·3pp [−3·2, +8·1], p = 0·431 |
| P, qwen − deepseek-v4.1-flash | +6·4pp [−0·2, +13·8], p = 0·140 | +5·3pp [−1·6, +12·4], p = 0·325 | +1·1pp [−4·4, +6·9], p = 0·844 |
| A×1, qwen − deepseek-v4.1-flash | +11·3pp [+3·0, +19·8], p = 0·012 | +3·2pp [−3·2, +9·9], p = 0·422 | +1·6pp [−5·1, +8·3], p = 0·682 |

On CPC with deepseek-v4.1-flash, MDT was numerically above A×1 at every endpoint but no contrast was individually significant (top-1 +4·8pp, p = 0·067; top-3 +3·0pp, p = 0·171; top-5 +3·7pp, p = 0·115). MDT exceeded the original P arm at top-3 and top-5 (+9·0 and +7·6 points), while P did not exceed A×1 in this tested prompt/model context. All five seeds were complete and adjudicated.

The P arm’s brace-repair counts for the five-seed arm: 56 of the 435 final P rows (12·9%) were recovered by the parser-level brace normalisation and flagged brace_repaired in the run files — 8 of 261 rows (3·1%) in seeds 1–3 and 48 of 174 rows (27·6%) in seeds 4–5. One property of the deepseek-v4.1-flash A×1 arm is worth noting: at requested temperature 0 its decision-level output is close to deterministic, so the five seeds agree on almost every case and the per-seed SDs are correspondingly small (0·5–1·0 points); pairwise seed comparisons differ in wording on 70–85 of 87 cases while the adjudicated outcome flips on at most a few. This is a property of the serving endpoint rather than a duplication — the five files are five genuine calls (per-case token counts differ between seeds), and it is also why the deepseek-v4.1-flash A×1 arm contributes less seed variance than the temperature-0.3 arms. The normalisation is confined to the parser: the prompt text was not changed and no diagnostic content was edited, so the two families remain strictly comparable.

**S11c. Emergency-corpus replication (ER-Reason, n = 364, five seeds; primary and sensitivity judges)**

The three strategies were re-run with deepseek-v4.1-flash on the **same 364 ER-Reason cases**, five seeds, and prompts byte-identical to the qwen arms. This is model-family replication, not an independent emergency cohort or external clinical validation. Case-text generation used OpenRouter’s zero-retention route pinned to one provider, including the regenerated A×1 baseline (local_rerun/ds_ax1/). All candidate–reference pairs were adjudicated under both judges with zero unresolved. One P-arm row returning an empty list was counted as a miss. Source: routing_study/results/local_rerun/er_ds_3arm.{json,md} (er_ds_ax1_zdr.py and er_ds_3arm_stats.py).

| Arm (deepseek-v4.1-flash, ER-Reason) | Top-1 | Top-3 | Top-5 |
| --- | --- | --- | --- |
| **Primary judge (GLM-5.3-flash × v3)** |  |  |  |
| A×1 | 43·4 ± 0·7 | 62·5 ± 1·9 | 70·4 ± 1·1 |
| P | 46·7 ± 0·8 | 57·0 ± 0·7 | 63·0 ± 0·7 |
| MDT | 40·1 ± 1·0 | 54·9 ± 0·7 | 62·5 ± 1·2 |
| **Sensitivity judge (deepseek-v4.1-flash × v3)** |  |  |  |
| A×1 | 37·6 ± 0·7 | 53·6 ± 1·1 | 60·2 ± 1·2 |
| P | 40·2 ± 0·5 | 47·7 ± 1·0 | 51·7 ± 0·7 |
| MDT | 33·0 ± 1·2 | 46·5 ± 0·6 | 51·8 ± 0·9 |

| Comparison | Judge | Top-1 | Top-3 | Top-5 |
| --- | --- | --- | --- | --- |
| MDT − A×1 | primary | −3·4pp [−6·0, −0·7], p = 0·016 | −7·5pp [−10·5, −4·6], p = 1·8 × 10⁻⁶ | −7·9pp [−11·0, −5·0], p = 4·7 × 10⁻⁷ |
| MDT − P | primary | −6·6pp [−9·7, −3·6], p = 2·8 × 10⁻⁵ | −2·0pp [−4·9, +0·8], p = 0·19 | −0·4pp [−3·4, +2·5], p = 0·89 |
| P − A×1 | primary | +3·2pp [+0·3, +6·2], p = 0·030 | −5·5pp [−8·4, −2·6], p = 1·9 × 10⁻⁴ | −7·5pp [−10·4, −4·7], p = 7·4 × 10⁻⁷ |
| MDT − A×1 | sensitivity | −4·6pp [−7·2, −2·0], p = 1·8 × 10⁻⁴ | −7·1pp [−9·9, −4·3], p = 6·7 × 10⁻⁷ | −8·4pp [−11·4, −5·4], p = 2·9 × 10⁻⁸ |
| MDT − P | sensitivity | −7·2pp [−10·1, −4·3], p = 5·6 × 10⁻⁷ | −1·1pp [−3·8, +1·4], p = 0·48 | +0·2pp [−2·5, +2·9], p = 0·89 |
| P − A×1 | sensitivity | +2·6pp [−0·4, +5·7], p = 0·10 | −5·9pp [−8·7, −3·2], p = 1·9 × 10⁻⁵ | −8·6pp [−11·5, −5·6], p = 8·1 × 10⁻⁸ |

Both judges reproduced A×1 > MDT at every endpoint on these same emergency cases, with P intermediate. Under the sensitivity judge, which shares a family with the generator, the recall-endpoint deficits were at least as large (top-5 −8·4 points). This consistency across model family and judge does not create an independent clinical validation cohort or eliminate shared label/judge limitations.

### Supplementary Table S12. Backbone scale (176B versus 2400B) and the isolated team versus a single larger call

Source: routing_study/results/scale_comparison.{json,md}, produced by routing_study/scripts/scale_comparison.py. The script is purely offline: it reads the existing CPC runs and the two frozen judge caches (judge_cache_glm_v3.json and judge_cache_dsflash_v3.json) and makes no model calls, writing to neither cache. Cases, prompts (promptv2, frozen), seeds (five), and the case-level statistics are those of the main experiment; all values are percentages or percentage-point differences on per-case five-seed hit rates. There are no missing primary-judge verdicts in any cell; the sensitivity-judge cache is frozen and does not cover the five re-run 2400B P rows (ten candidate–reference pairs, in seeds 2, 4 and 5), so the sensitivity-judge comparison rows that involve P @2400B are reported on n = 85 at top-1 and n = 87 at top-3 and top-5, the top-1 cell being the one reduced by unadjudicated pairs.

**S12a. Per-seed accuracy under both judges (n = 87, five seeds)**

| Arm | Judge | Top-1 | Top-3 | Top-5 |
| --- | --- | --- | --- | --- |
| A×1 @176B | primary (GLM-5.3-flash × v3) | 57·9 ± 1·9 | 76·1 ± 4·1 | 81·1 ± 2·6 |
| A×1 @2400B | primary | 62·8 ± 1·0 | 79·3 ± 1·4 | 86·2 ± 1·1 |
| P @176B | primary | 53·8 ± 4·0 | 72·2 ± 2·6 | 76·8 ± 2·1 |
| P @2400B | primary | 62·3 ± 1·7 | 76·1 ± 2·5 | 83·9 ± 1·4 |
| MDT @176B (six calls) | primary | 60·5 ± 3·1 | 81·6 ± 2·4 | 85·5 ± 2·1 |
| A×1 @176B | sensitivity (deepseek-v4.1-flash × v3) | 51·7 ± 1·8 | 72·6 ± 3·8 | 77·7 ± 2·6 |
| A×1 @2400B | sensitivity | 59·1 ± 1·9 | 75·2 ± 1·7 | 82·5 ± 2·2 |
| P @176B | sensitivity | 50·1 ± 3·8 | 69·7 ± 2·6 | 75·2 ± 1·9 |
| P @2400B | sensitivity | 58·4 ± 2·5 | 72·2 ± 3·5 | 79·8 ± 2·5 |
| MDT @176B (six calls) | sensitivity | 57·0 ± 2·1 | 78·4 ± 1·5 | 82·3 ± 1·3 |

**S12b. Scale effect and scale-by-strategy interaction (2400B − 176B)**

| Contrast | Judge | Top-1 | Top-3 | Top-5 |
| --- | --- | --- | --- | --- |
| A×1 @2400B − A×1 @176B | primary | +4·8pp [−4·6, +14·3], p = 0·41 | +3·2pp [−3·9, +10·3], p = 0·37 | +5·1pp [−0·9, +11·0], p = 0·12 |
| P @2400B − P @176B | primary | +8·5pp [−0·2, +17·5], p = 0·066 | +3·9pp [−3·0, +10·8], p = 0·30 | +7·1pp [+1·6, +12·9], p = 0·016 |
| interaction (ΔA×1 − ΔP) | primary | −3·7pp [−14·3, +7·1], p = 0·51 | −0·7pp [−9·0, +7·6], p = 0·81 | −2·1pp [−8·3, +4·1], p = 0·43 |
| A×1 @2400B − A×1 @176B | sensitivity | +7·4pp [−3·0, +17·5], p = 0·20 | +2·5pp [−5·5, +10·8], p = 0·54 | +4·8pp [−1·8, +11·7], p = 0·20 |
| P @2400B − P @176B | sensitivity | +8·5pp [−0·5, +17·4], p = 0·081 | +2·5pp [−4·8, +9·9], p = 0·47 | +4·6pp [−1·6, +11·0], p = 0·10 |
| interaction (ΔA×1 − ΔP) | sensitivity | −0·9pp [−12·0, +9·9], p = 0·83 | +0·0pp [−9·0, +9·0], p = 0·99 | +0·2pp [−6·4, +7·1], p = 0·91 |

Under both judges the scale effect is real but smaller than the earlier draft claimed, and the scale-by-strategy interaction is null: the four-condition Friedman test over A×1/P at 176B/2400B is not significant at top-1 and top-3 under either judge (primary χ² = 7·13, p = 0·068 and χ² = 4·77, p = 0·19; sensitivity χ² = 5·96, p = 0·11 and χ² = 2·75, p = 0·43) and reaches significance only at top-5 under the primary judge (χ² = 12·63, p = 0·0055), which indicates a main effect of scale or strategy rather than an interaction.

**S12c. The isolated team at 176B versus a single 2400B call (MDT @176B − A×1 @2400B)**

| Judge | Top-1 | Top-3 | Top-5 |
| --- | --- | --- | --- |
| primary | −2·3pp [−12·0, +7·4], p = 0·72 | +2·3pp [−4·8, +9·7], p = 0·50 | −0·7pp [−6·2, +4·8], p = 0·90 |
| sensitivity | −2·1pp [−12·0, +8·0], p = 0·73 | +3·2pp [−4·6, +10·8], p = 0·37 | −0·2pp [−6·4, +5·7], p = 0·96 |

Six calls through five isolated roles at 176B therefore match a single call with an ≈14-fold larger backbone: the top-5 difference is −0·7 points under the primary judge and −0·2 under the sensitivity judge, neither significant, and at top-3 the team is numerically ahead (81·6% versus 79·3%) by a margin that is likewise not significant. The “matching” claim is a statement of statistical indistinguishability at these sample sizes rather than of a demonstrated equality.

**S12d. Completeness check for the auxiliary 2400B P arm**

The 2400B P arm contained five rows with an empty output, which were re-run before this analysis and written back into the affected files (the pre-repair files are retained as P_s*.jsonl.bak_prefill; the repair is logged in routing_study/results/refill_defects.log). Dropping the five affected cases altogether (n = 82) leaves every contrast above in the same direction and changes no conclusion: under the primary judge the A×1 scale effect becomes +2·7 / +1·2 / +3·2 points (p = 0·63 / 0·70 / 0·31), P @2400B − P @176B becomes +5·6 / +1·7 / +5·4 points (p = 0·22 / 0·70 / 0·047), and MDT @176B − A×1 @2400B becomes +0·5 / +3·9 / +0·7 points (p = 0·85 / 0·27 / 0·74) at top-1/3/5. The sensitivity judge shows the same pattern.

### Supplementary Note. Prompt transcription defect and the corrected-prompt control

One prompt constant contained a transcription defect, and we document it and its control here. P_TOPN_SUFFIX (routing_study/scripts/topn_cpc_promptv2.py) is a plain string that is never passed through Python’s .format(), so the JSON example it carries is sent literally with doubled braces ({{"top5": [{{"rank": 1, ...}}]}}), whereas A_TOPN_PROMPT, which *is* passed through .format(), sends the same example with single braces. Every caller concatenates the prompt as PERSPECTIVE_PROMPT.format(structured_case=...) + P_TOPN_SUFFIX, so the defect is confined to the P arm and is identical on all three datasets; the A×1 and MDT arms are unaffected. The byte-exact text actually sent, together with SHA-256 fingerprints of PERSPECTIVE_PROMPT, P_TOPN_SUFFIX, the concatenated prompt, and A_TOPN_PROMPT, is deposited as routing_study/results/prompt_actually_sent.md, which is generated from the constants themselves rather than transcribed by hand; the same file shows the line-by-line diff against the intent version, in which exactly one line differs and only in its braces. The version reproduced in Supplementary Methods (S-M2) is the intent version.

*Qwen3.8-flash side.* Across the 6,105 P-arm outputs of the main experiment — CPC (87 × 5 = 435), MedCaseReasoning (406 × 5 = 2,030), ER-Reason (364 × 5 = 1,820) and the ER-Reason workup condition (364 × 5 = 1,820) — no output was empty and none contained fewer than five candidates. qwen3.8-flash rewrites the doubled braces itself, so the defect left no trace in the main-experiment numbers. The one exception among the qwen-side arms lies outside these 6,105 outputs: the auxiliary 2400B P arm had five empty rows, which were re-run (Supplementary Table S12 and the data-integrity note below).

*Deepseek-flash side.* The same outputs are not self-correcting. deepseek-v4.1-flash echoes the doubled braces verbatim, so the parser first finds no valid JSON and the generator retries (up to three attempts, as in the main runs). In the family’s seeds 1–3, 57 of 261 P-arm rows (21·8%) returned an unparseable list on the first attempt; across the full five-seed arm, 56 of 435 final rows (12·9%; 8 of 261 in seeds 1–3 and 48 of 174 in seeds 4–5, the same five-seed counts as Supplementary Table S11’s note) are flagged brace_repaired: true — of the 57 unparseable first attempts in seeds 1–3, forty-nine were recovered on retry and eight by a parser pass that folded the echoed braces ({{/}}) back to single ones (the seeds 4–5 extension follows the main runs’ retry logic). The normalisation is confined to the parser: the prompt text was not changed and no diagnostic content was edited, so the two families remain strictly comparable and the affected rows are described in the table note of Supplementary Table S11.

*Control.* The effect of the defect on the qwen-side conclusions is quantified by the corrected-prompt arm (Pfixed) in Supplementary Table S10c, which differs from the main P arm only in the folded braces. Because the defect and its correction concern the prompt constant rather than the model, the same control was repeated on the external set (MedCaseReasoning); the arm, its prompt-equivalence audit, and its case-level statistics are reported in Tables S13a–S13b below.

*External replication of the control.* Because the defect and the correction both concern the prompt constant rather than the model or the corpus, the corrected-prompt arm was re-run on MedCaseReasoning, the external benchmark, under the same five-seed protocol and the same frozen primary judge (GLM-5.3-flash × v3), with the inference and judging pipeline of the main MCR P arm reused verbatim (topn_mcr.call_top5; temperature = 0·3, thinking disabled, max_tokens = 4096), so the prompt suffix is the only difference from that arm. All statistics are case-level: each of the 406 cases contributes its five-seed hit rate, arms are compared across cases with the paired two-sided Wilcoxon signed-rank test and case-level cluster-bootstrap 95% confidence intervals (10,000 resamples), and the secondary exact McNemar test uses majority-vote outcomes (a case hit when at least three of five seeds hit). No candidate–reference pair is missing in any arm and no case is excluded.

**Supplementary Table S13a. Prompt-equivalence audit for the corrected-prompt control, CPC and MedCaseReasoning**

| Quantity | CPC control (S10c) | External control (S13b) |
| --- | --- | --- |
| Cases audited | 87 | 406 |
| Cases with the same line count, exactly one line differing, and that line differing only in brace folding | 87/87 | 406/406 |
| Anomalies | 0 | 0 |
| Per-case character difference | −12 | −12 |

The audit is run by the analysis script itself (routing_study/scripts/p_fixed_mcr.py) on the two complete prompts of every case and is asserted case by case inside the script rather than checked by hand: on all 406 cases the two prompts have the same number of lines, differ on exactly one line, and differ there only by folding each {{/}} of the output-format suffix to {/} (six escaped pairs per case, hence the per-case character difference of −12; the differing line number varies with the case text, on the observed range 46–77). The unfixed suffix (P_TOPN_SUFFIX, sha256 39fe609b…) and the folded suffix (P_FIXED_SUFFIX, sha256 606d280f…) are both fingerprinted in the script, and the shared constant is untouched, so no main-experiment result moves.

**Supplementary Table S13b. Corrected-prompt control on the external set: the P arm on MedCaseReasoning with the doubled braces folded (Pfixed), n = 406, five seeds, primary judge**

Source: routing_study/results/p_fixed_mcr.{json,md}, produced by routing_study/scripts/p_fixed_mcr.py from routing_study/results/topn_seeds_mcr_pfixed/Pfixed_s{1..5}.jsonl (406 cases × 5 seeds = 2,030 calls). Every value is on the case-level, five-seed-hit-rate scale described above; differences are the first-named arm minus the second, in percentage points.

| Arm | Top-1 | Top-3 | Top-5 |
| --- | --- | --- | --- |
| Pfixed (braces folded) | 57·2 ± 0·9 | 70·5 ± 0·8 | 76·0 ± 1·2 |
| P (main experiment, literal braces) | 56·4 ± 0·3 | 69·2 ± 1·1 | 75·0 ± 1·0 |
| A×1 (T = 0), reference | 57·0 ± 0·9 | 69·9 ± 0·6 | 75·6 ± 0·8 |
| MDT (T = 0·3), reference | 55·9 ± 0·8 | 72·9 ± 0·8 | 79·6 ± 0·8 |

| Comparison | Top-k | Hit rates (A vs B) | Mean difference [95% CI] | Wilcoxon p | Majority-vote McNemar (A-only : B-only, p) |
| --- | --- | --- | --- | --- | --- |
| Pfixed − P | top-1 | 57·2% vs 56·4% | +0·8pp [−0·4, +2·1] | 0·165 | 11 : 4, p = 0·119 |
| Pfixed − P | top-3 | 70·5% vs 69·2% | +1·3pp [−0·0, +2·8] | 0·098 | 15 : 7, p = 0·134 |
| Pfixed − P | top-5 | 76·0% vs 75·0% | +0·9pp [−0·6, +2·5] | 0·242 | 15 : 9, p = 0·307 |
| Pfixed − A×1 | top-1 | 57·2% vs 57·0% | +0·2pp [−2·2, +2·6] | 0·668 | 19 : 16, p = 0·736 |
| Pfixed − A×1 | top-3 | 70·5% vs 69·9% | +0·6pp [−1·8, +3·1] | 0·452 | 24 : 18, p = 0·441 |
| Pfixed − A×1 | top-5 | 76·0% vs 75·6% | +0·3pp [−2·0, +2·7] | 0·563 | 27 : 20, p = 0·382 |
| Pfixed − MDT | top-1 | 57·2% vs 55·9% | +1·3pp [−0·9, +3·7] | 0·242 | 19 : 16, p = 0·736 |
| Pfixed − MDT | top-3 | 70·5% vs 72·9% | −2·4pp [−4·5, −0·3] | 0·0501 | 12 : 25, p = 0·047 |
| Pfixed − MDT | top-5 | 76·0% vs 79·6% | −3·6pp [−5·7, −1·5] | 0·0007 | 13 : 26, p = 0·053 |

Two consequences follow, and both differ from CPC. First, the defect has no measurable effect on this corpus: folding the braces moves the P arm by less than 1·5 points at every endpoint and none of the three contrasts reaches significance (all p ≥ 0·098), so the three-to-four-point CPC effect of the same correction does not generalise to the external set. Second, the team’s recall advantage over persona survives the correction: MDT is ahead of Pfixed at top-5 (−3·6pp, p = 0·0007) with a borderline difference at top-3 (−2·4pp, p = 0·0501), while at top-1 Pfixed is numerically ahead (+1·3pp, p = 0·242). The CPC reading — that the corrected prompt leaves the team’s recall advantage over persona no longer individually significant at either recall endpoint — therefore does not reproduce on the 406-case external set. The corrected arm also still fails to beat the single call at every endpoint (+0·2 / +0·6 / +0·3 points, p = 0·668 / 0·452 / 0·563).

### Supplementary Note. Data integrity repair

Two data-integrity defects were found after the main runs and repaired before the analyses reported here. Neither is a difference in model behaviour: both were missing input at the level of the run files, and each entered the aggregate as if it were a genuine response.

*MedCaseReasoning MDT role calls.* In the MedCaseReasoning MDT arm, 16 role calls returned an empty candidate list — five in seed 1, five in seed 2, one each in seeds 3 and 4, and four in seed 5 — and the moderator synthesis treated each empty list as one of its five panel opinions, so the affected case-seeds were synthesised from four opinions plus a blank. We re-ran those 16 role calls, re-synthesised the 16 affected case-seeds with the moderator, and re-adjudicated the resulting candidate lists against the same frozen judge and cache (routing_study/results/judge_cache_glm_v3.json), with no other change to the judging setup. The pre-repair files are retained as *.bak_prefill beside their repaired counterparts, and the repair is logged in routing_study/results/refill_defects.log. The change to the reported numbers is at most 0·2 points: MDT top-1 56·0% → 55·9%, top-3 73·0% → 72·9%, top-5 79·6% (unchanged); the MDT-versus-A×1 top-3 contrast moves from p = 0·0055 to p = 0·0079 and the MDT-versus-P top-3 contrast from p = 0·0018 to p = 0·0027, both still significant, so no conclusion depends on the repair. The repaired values are the ones in main-text Table 1, in Supplementary Tables S1 and S4, and in the pooled-seed legacy table (S3), where several discordant-pair counts move by one or two pairs for the same reason.

*Auxiliary 2400B P arm.* The 2400B P arm contained five rows with an empty output; those rows were re-run and written back into the affected files (P_s*.jsonl.bak_prefill backups, same log). Their effect is reported in the note under Supplementary Table S12: excluding the five affected cases leaves every contrast unchanged in direction and in significance.

### Supplementary Table S15. GEE sensitivity analysis: the 27 primary tests re-fit as case-clustered observation-level models

Source: routing_study/results/local_rerun/gee_sensitivity_local.{json,md}, produced by routing_study/scripts/gee_sensitivity_local.py (offline; 2026-10-05 variant — the ER-Reason models read the self-hosted rerun’s run files, while the CPC and MedCaseReasoning models are unchanged from routing_study/results/gee_sensitivity.{json,md}; it reads the run files and the frozen primary judge cache and makes no model calls). The primary analysis takes the case as the unit of inference — each case contributing its five-seed hit rate — with the paired Wilcoxon signed-rank test, which discards tied cases and does not propagate seed-level variance. This table re-fits every test of the primary framework as an observation-level logistic generalized estimating equation (GEE; binomial family, exchangeable working correlation, clusters = cases), with one model per dataset × comparison × endpoint; the observation count is 2 × n × 5 per comparison (870 for CPC, 4,060 for MedCaseReasoning, 3,640 for ER-Reason). Entries are the arm effect of the first-named strategy: β (log-odds ratio), OR with robust 95% CI, and Wald p. Hit flags come from the same frozen cache as the primary analysis, so the two frameworks score identical data.

| Dataset | Comparison | Endpoint | β | OR [95% CI] | Wald p |
| --- | --- | --- | --- | --- | --- |
| CPC | MDT vs A×1 | top-1 | +0·105 | 1·11 [0·92, 1·35] | 0·288 |
|  |  | top-3 | +0·332 | 1·39 [1·07, 1·83] | 0·0155 |
|  |  | top-5 | +0·316 | 1·37 [1·06, 1·78] | 0·0181 |
| CPC | MDT vs P | top-1 | +0·273 | 1·31 [1·00, 1·72] | 0·0463 |
|  |  | top-3 | +0·536 | 1·71 [1·19, 2·45] | 0·0036 |
|  |  | top-5 | +0·580 | 1·79 [1·21, 2·64] | 0·0037 |
| CPC | P vs A×1 | top-1 | −0·168 | 0·85 [0·66, 1·08] | 0·176 |
|  |  | top-3 | −0·204 | 0·82 [0·60, 1·10] | 0·183 |
|  |  | top-5 | −0·264 | 0·77 [0·55, 1·08] | 0·126 |
| MedCaseReasoning | MDT vs A×1 | top-1 | −0·046 | 0·95 [0·87, 1·05] | 0·337 |
|  |  | top-3 | +0·147 | 1·16 [1·05, 1·28] | 0·0049 |
|  |  | top-5 | +0·227 | 1·25 [1·13, 1·40] | < 0·0001 |
| MedCaseReasoning | MDT vs P | top-1 | −0·020 | 0·98 [0·89, 1·08] | 0·675 |
|  |  | top-3 | +0·182 | 1·20 [1·08, 1·33] | 0·0008 |
|  |  | top-5 | +0·259 | 1·30 [1·14, 1·47] | < 0·0001 |
| MedCaseReasoning | P vs A×1 | top-1 | −0·026 | 0·97 [0·89, 1·07] | 0·584 |
|  |  | top-3 | −0·035 | 0·97 [0·86, 1·09] | 0·566 |
|  |  | top-5 | −0·032 | 0·97 [0·85, 1·10] | 0·634 |
| ER-Reason | MDT vs A×1 | top-1 | −0·247 | 0·78 [0·69, 0·88] | < 0·0001 |
|  |  | top-3 | −0·361 | 0·70 [0·62, 0·78] | < 0·0001 |
|  |  | top-5 | −0·400 | 0·67 [0·60, 0·75] | < 0·0001 |
| ER-Reason | MDT vs P | top-1 | −0·146 | 0·86 [0·77, 0·98] | 0·019 |
|  |  | top-3 | −0·117 | 0·89 [0·79, 1·00] | 0·043 |
|  |  | top-5 | −0·133 | 0·88 [0·78, 0·99] | 0·030 |
| ER-Reason | P vs A×1 | top-1 | −0·102 | 0·90 [0·80, 1·02] | 0·113 |
|  |  | top-3 | −0·244 | 0·78 [0·69, 0·89] | 0·0002 |
|  |  | top-5 | −0·267 | 0·77 [0·68, 0·86] | < 0·0001 |

All 27 models converged. The GEE reproduces every primary conclusion in direction and significance, with two details worth noting. First, it is *more* sensitive than the tied-discarding case-level test in tie-heavy cells: on the CPC top-1 contrast it moves MDT-versus-P to significance (OR 1·31, p = 0·046), consistent with the legacy pooled-seed McNemar table (S3), which had flagged that contrast before being retired. Second, it returns the single contrast that fell to the primary Benjamini–Hochberg correction — the ER-Reason MDT-versus-P top-3 comparison — to significance (OR 0·89, 95% CI 0·79–1·00, p = 0·043), so the one “no conclusion” carried in the main text is a property of the tied-discarding framework rather than of the data. No primary conclusion is weakened under the GEE.

### Supplementary Table S16. Majority-vote threshold sensitivity: the 27 primary tests at t = 2/3/4/5 of five seeds

Source: routing_study/results/majority_threshold_sensitivity.{json,md} (routing_study/scripts/majority_threshold_sensitivity.py, offline); the ER-Reason rows of the current table are recomputed from the self-hosted rerun (2026-10-05) by routing_study/scripts/er_local_extras.py (threshold section of routing_study/results/local_rerun/extras.json) under the same t-of-5 convention. The secondary analysis of the main text scores a case as a majority-vote hit at ≥3 of 5 seeds. This table recomputes the exact McNemar test for all 27 primary contrasts at thresholds t = 2, 3, 4 and 5 of five seeds. The pre-specified threshold t = 3 is row-highlighted by design; t = 2 maximises sensitivity, t = 5 (unanimous) minimises it.

| Dataset | Comparison | Endpoint | t=2 (a:b, p) | t=3 (a:b, p) | t=4 (a:b, p) | t=5 (a:b, p) |
| --- | --- | --- | --- | --- | --- | --- |
| CPC | MDT vs A×1 | top-1 | 5:0, 0·063 | 7:2, 0·180 | 7:5, 0·774 | 3:6, 0·508 |
| CPC | MDT vs A×1 | top-3 | 6:1, 0·125 | 8:3, 0·227 | 8:2, 0·109 | 7:4, 0·549 |
| CPC | MDT vs A×1 | top-5 | 4:0, 0·125 | 4:2, 0·688 | 7:1, 0·070 | 6:2, 0·289 |
| CPC | MDT vs P | top-1 | 12:3, 0·035 | 10:3, 0·092 | 9:4, 0·267 | 7:4, 0·549 |
| CPC | MDT vs P | top-3 | 9:3, 0·146 | 14:6, 0·115 | 14:3, 0·013 | 13:2, 0·007 |
| CPC | MDT vs P | top-5 | 7:1, 0·070 | 9:5, 0·424 | 15:3, 0·008 | 15:1, 0·0005 |
| CPC | P vs A×1 | top-1 | 4:8, 0·388 | 4:6, 0·754 | 4:7, 0·549 | 2:8, 0·109 |
| CPC | P vs A×1 | top-3 | 4:5, 1·000 | 4:7, 0·549 | 4:9, 0·267 | 1:9, 0·022 |
| CPC | P vs A×1 | top-5 | 3:5, 0·726 | 4:6, 0·754 | 5:11, 0·210 | 1:11, 0·006 |
| MCR | MDT vs A×1 | top-1 | 15:17, 0·860 | 16:16, 1·000 | 14:22, 0·243 | 20:24, 0·652 |
| MCR | MDT vs A×1 | top-3 | 24:11, 0·041 | 26:7, 0·0013 | 21:10, 0·071 | 21:18, 0·749 |
| MCR | MDT vs A×1 | top-5 | 26:7, 0·0013 | 25:5, 0·0003 | 21:6, 0·0059 | 22:14, 0·243 |
| MCR | MDT vs P | top-1 | 14:25, 0·108 | 21:17, 0·627 | 23:20, 0·761 | 29:18, 0·144 |
| MCR | MDT vs P | top-3 | 25:15, 0·154 | 31:10, 0·0015 | 29:10, 0·0034 | 36:11, 0·0003 |
| MCR | MDT vs P | top-5 | 25:12, 0·047 | 29:10, 0·0034 | 32:11, 0·0019 | 41:8, <0·0001 |
| MCR | P vs A×1 | top-1 | 23:14, 0·188 | 17:21, 0·627 | 15:26, 0·117 | 16:31, 0·040 |
| MCR | P vs A×1 | top-3 | 24:21, 0·766 | 19:21, 0·875 | 16:24, 0·268 | 17:39, 0·005 |
| MCR | P vs A×1 | top-5 | 23:17, 0·430 | 22:21, 1·000 | 19:25, 0·451 | 16:41, 0·0013 |
| ER-Reason | MDT vs A×1 | top-1 | 14:17, 0·72 | 11:23, 0·058 | 6:31, <0·0001 | 3:45, <0·0001 |
| ER-Reason | MDT vs A×1 | top-3 | 18:27, 0·23 | 12:36, 0·0007 | 8:45, <0·0001 | 4:70, <0·0001 |
| ER-Reason | MDT vs A×1 | top-5 | 26:32, 0·51 | 15:37, 0·0032 | 12:52, <0·0001 | 5:75, <0·0001 |
| ER-Reason | MDT vs P | top-1 | 17:23, 0·43 | 16:24, 0·27 | 19:30, 0·15 | 16:27, 0·13 |
| ER-Reason | MDT vs P | top-3 | 20:24, 0·65 | 22:27, 0·57 | 28:27, 1·0 | 19:34, 0·053 |
| ER-Reason | MDT vs P | top-5 | 23:27, 0·67 | 24:23, 1·0 | 28:25, 0·78 | 23:36, 0·12 |
| ER-Reason | P vs A×1 | top-1 | 21:18, 0·75 | 18:22, 0·64 | 13:27, 0·038 | 8:39, <0·0001 |
| ER-Reason | P vs A×1 | top-3 | 20:25, 0·55 | 13:32, 0·0066 | 11:49, <0·0001 | 7:58, <0·0001 |
| ER-Reason | P vs A×1 | top-5 | 23:25, 0·89 | 13:36, 0·0014 | 8:51, <0·0001 | 6:63, <0·0001 |

Reading: the three headline conclusions are threshold-robust. The emergency reversal (ER MDT-vs-A×1) is significant at the pre-specified threshold t = 3 and at both stricter thresholds, t = 4 and t = 5, on every endpoint, and A×1-vs-P is significant at t = 3/4/5 on the recall endpoints; under the loosest criterion, t = 2, no ER contrast separates — a 2-of-5 standard credits an arm that misses on three of the five seeds with a hit. The MCR and CPC recall advantages of MDT over A×1 and P are significant at t = 3 and at one or both neighbouring thresholds, weakening only at t = 5, where the unanimous criterion discards most of the evidence (per-seed hit rates of 4/5 fail it). The two contrasts that move with the threshold in the other direction (CPC MDT-vs-P, significant only at t ≥ 4; MCR MDT-vs-A×1 top-3, significant at t ≤ 3) are consistent with a small true effect measured with seed-level noise and carry no conclusion in the main text. No primary conclusion reverses sign at any threshold.

### Supplementary Figure S1

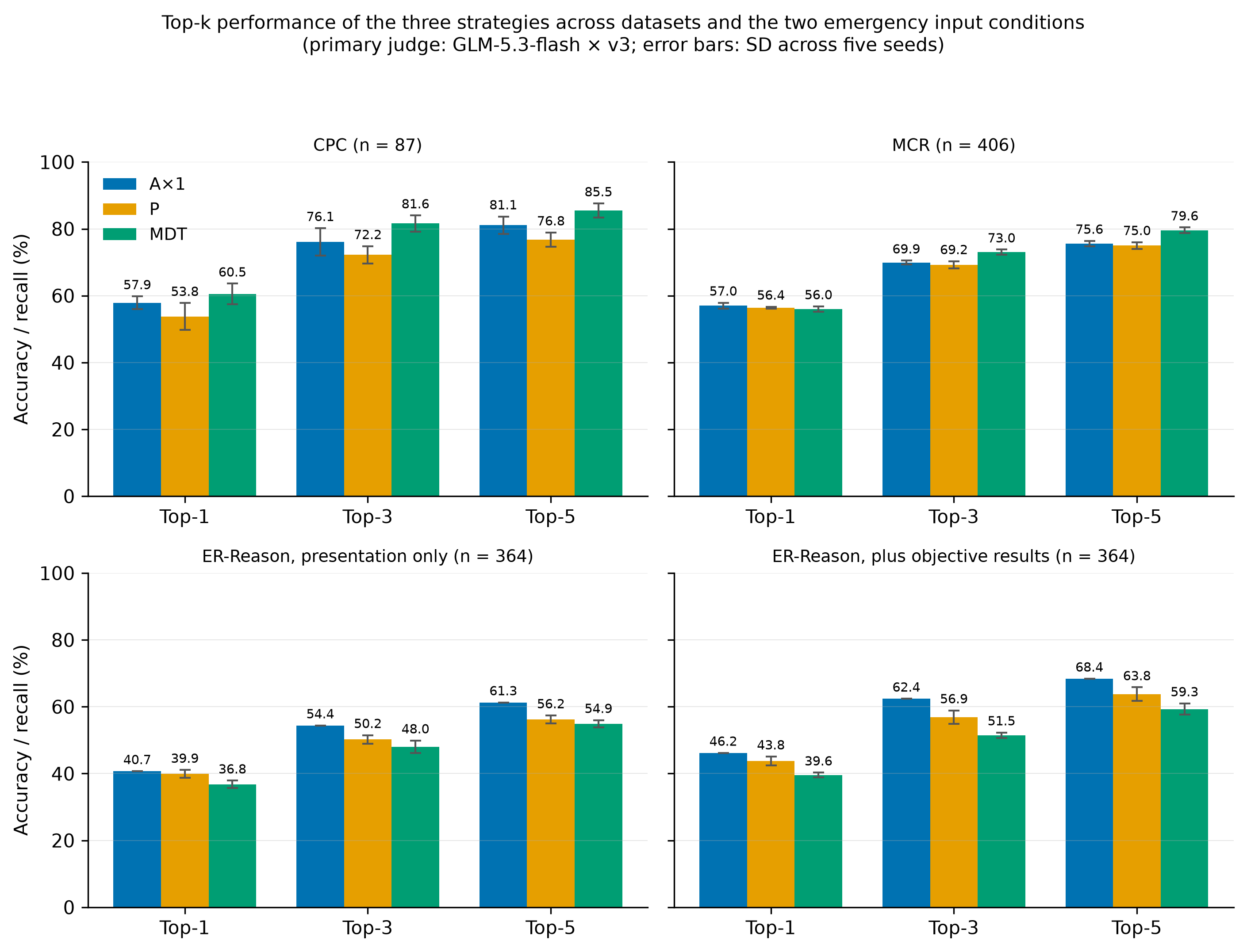

Top-k performance of the three strategies across datasets and across the two emergency input conditions

**Figure S1. Top-k performance of the three strategies across the datasets and across the two emergency input conditions (primary judge, GLM-5.3-flash × v3).** Bars give top-1 accuracy and top-3/top-5 recall (%); error bars are SD across the five seeds. Panels: CPC (n = 87); MCR (n = 406); ER-Reason under presentation-only input (n = 364); ER-Reason under presentation plus the objective results of the visit (n = 364). Colors follow the Okabe–Ito colorblind-safe palette (A×1 blue, P orange, MDT green). Values are the recomputed per-seed figures of Supplementary Table S1 (CPC, MCR, presentation-only ER-Reason) and of Supplementary Table S6 (workup-informed ER-Reason). Generated by routing_study/scripts/make_supplement_figure.py (matplotlib, 300 dpi); the self-hosted-rerun variant of 2026-10-05 is routing_study/scripts/make_supplement_figure_local.py, writing routing_study/results/local_rerun/supplementary_figure_s1_local.png.

#### Reproducibility notes

- **Primary tables (S1, S3, S4, S7):** make_supplement_tables.py, stats_caselevel.py, and holdout_primary.py; current ER-Reason values use make_supplement_tables_local.py and local_rerun/topn_erreason/seed_summary.json. Outputs include stats_caselevel.{json,md} and holdout46_primary.{json,md}.
- **Workup analyses (S6, S9):** eval_workup_local.py reads local_rerun/topn_erreason_workup/ and writes local_rerun/workup_vs_baseline_local.{json,md}. The no-objective and 20-case leakage exclusions are computed in er_local_extras.py and local_rerun/extras.json.
- **Sensitivity judge and clinician agreement (S2, S5):** frozen cache judge_cache_dsflash_v3.json; case-level contrasts s2b_caselevel_ds.py and s2b_caselevel_ds.json; annotation and κ artifacts in routing_study/results/judge_validity/. Batch-1 rule-development results are tuning-contaminated; batch 2 and the corpus-specific panels are the relevant held-out evaluations.
- **Legacy statistics (S3):** routing_study/results/stats_caselevel.{json,md}.
- **Temperature matching:** temp_matched_caselevel.py (CPC), ax1_t03_mcr.py (MCR), and the ER-Reason analysis over local_rerun/topn_erreason_ax1t03/ and local_rerun/topn_erreason/. Analysis is offline against the frozen cache.
- **Word coverage (S34):** word_coverage_local.py for ER-Reason and word_coverage_cpc_mcr.py for CPC/MCR, using five seeds and the repaired run files.
- **Judge agreement (S8):** judge_agreement_local.py reads the two frozen caches and per-case outputs and writes local_rerun/judge_agreement_by_stratum_local.{json,md}. Judge traffic contains reference labels and candidate strings only.
- **Prompt-transcription audit (S13):** byte-exact prompts and fingerprints are in prompt_actually_sent.md; CPC and MCR controls use p_fixed_cpc.py and p_fixed_mcr.py. The shared prompt constant is unchanged, and each corrected prompt is audited against the original case by case.
- **Matched controls (S10a–S10c):** ax1_cot_cpc.py, ax5_full87.py, and p_fixed_cpc.py; outputs ax1_cot_cpc.{json,md}, ax5_full87.{json,md}, and p_fixed_cpc.{json,md}.
- **Moderator and factorial controls (S10d–S10i):** mdt_nomoderator_cpc.py, ax5_mod_cpc.py, ax5_mod_er.py, p_split_cpc.py, moderator_anatomy.py, moderator_anatomy_full.py, and ax5_mod_mcr.py. These scripts preserve the role/sample inputs, prompt variants, and case-level analyses described in each panel.
- **GEE and threshold sensitivity (S15, S16):** gee_sensitivity.py with gee_sensitivity_local.py for ER-Reason; majority_threshold_sensitivity.py with the threshold section of local_rerun/extras.json for current ER-Reason rows.
- **Second family and scale (S11, S12):** seeds_87_dsflash.py, mdt_cpc.py, er_ds_3arm_stats.py, and scale_comparison.py; outputs include dsflash_family_cpc.{json,md}, local_rerun/er_ds_3arm.{json,md}, and scale_comparison.{json,md}.
- **Data-integrity repair (S14):** refill_defects.log records role-call refills, moderator re-syntheses, and the 2400B P refill; pre-repair files are retained as *.bak_prefill. Repaired outputs were re-adjudicated under the same frozen judge.
- **Emergency serving and routing:** current qwen ER-Reason outputs are under local_rerun/ from the self-hosted 4-bit stack. The deepseek same-cohort replication uses the zero-retention pinned route and regenerated A×1 baseline in local_rerun/ds_ax1/; the direct-API S10f-2 arm remains withdrawn.
- **Cache handling:** judge_shards.py and merge_judge_shards.py manage arm-specific shards, conflict checks, and consolidation. Reported analyses use the consolidated caches and complete adjudication unless a panel explicitly states otherwise.

**S17. The three strategies on a closed flagship (gpt-5.1, CPC A×1 / P / MDT)**

To test whether the family replications above depend on staying within the open-weight tier, the three strategies were re-run end to end on gpt-5.1 (deployment string gpt-5.1-2025-11-13, OpenAI-compatible endpoint), 87 CPC cases × 5 seeds, prompts byte-identical to the main experiment (A×1 T = 0, max_tokens 2,048; P / roles / moderator T = 0·3, max_tokens 4,096; a homogeneous gpt-5.1 team). Adjudication used the same frozen primary-judge cache; a completion-gated retry loop drove unresolved pairs to zero before the analysis was produced, superseding an interim analysis computed on an incomplete adjudication under a provider degradation window, which is retained only as a voided artifact.

| Arm | top-1 | top-3 | top-5 |
| --- | --- | --- | --- |
| A×1 (gpt-5.1) | 54·9 ± 1·9 | 71·0 ± 3·0 | 82·5 ± 3·0 |
| P (gpt-5.1) | 62·1 ± 1·6 | 77·9 ± 2·2 | 82·3 ± 1·3 |
| MDT (gpt-5.1) | 60·2 ± 1·7 | 81·6 ± 2·6 | 88·3 ± 2·6 |

Arm-level values are per-seed means ± SD across five seeds, the manuscript’s display convention; the corresponding case-level five-seed hit-rate means for MDT are 60·0/81·6/88·4% (differences ≤ 0·2 points; no contrast is affected). | A×1 (qwen3.8-flash, reference) | 57·9 ± 1·9 | 76·1 ± 4·1 | 81·1 ± 2·6 | | P (qwen3.8-flash, reference) | 53·8 ± 4·0 | 72·2 ± 2·6 | 76·8 ± 2·1 | | MDT (qwen3.8-flash, reference) | 60·5 ± 3·1 | 81·6 ± 2·4 | 85·5 ± 2·1 |

| Contrast | top-k | mean difference [95% CI] | Wilcoxon p |
| --- | --- | --- | --- |
| MDT (gpt-5.1) vs A×1 (gpt-5.1) | top-1 | +5·6pp [+1·4, +10·2] | 0·0161* |
| MDT (gpt-5.1) vs A×1 (gpt-5.1) | top-3 | +10·9pp [+5·8, +16·5] | 0·0003* |
| MDT (gpt-5.1) vs A×1 (gpt-5.1) | top-5 | +6·0pp [+2·6, +9·8] | 0·0053* |
| MDT (gpt-5.1) vs P (gpt-5.1) | top-1 | −1·6pp [−6·7, +3·5] | 0·2499 |
| MDT (gpt-5.1) vs P (gpt-5.1) | top-3 | +4·0pp [−0·2, +8·6] | 0·1828 |
| MDT (gpt-5.1) vs P (gpt-5.1) | top-5 | +6·3pp [+2·1, +10·7] | 0·0119* |
| P (gpt-5.1) vs A×1 (gpt-5.1) | top-1 | +7·1pp [+3·0, +11·5] | 0·0009* |
| P (gpt-5.1) vs A×1 (gpt-5.1) | top-3 | +6·9pp [+2·1, +12·0] | 0·0070* |
| P (gpt-5.1) vs A×1 (gpt-5.1) | top-5 | −0·2pp [−3·9, +3·7] | 0·7709 |
| MDT (qwen) vs MDT (gpt-5.1) | top-1 | +0·0pp [−9·5, +9·3] | 0·9344 |
| MDT (qwen) vs MDT (gpt-5.1) | top-3 | −0·2pp [−7·2, +7·0] | 0·9085 |
| MDT (qwen) vs MDT (gpt-5.1) | top-5 | −3·0pp [−8·1, +1·6] | 0·2506 |
| P (qwen) vs P (gpt-5.1) | top-1 | −8·3pp [−16·8, +0·2] | 0·0614（legacy pooled case–seed McNemar 36:72, p = 0·0007; see S3 caveat） |
| P (qwen) vs P (gpt-5.1) | top-3 | −5·7pp [−12·4, +0·9] | 0·1265 |
| P (qwen) vs P (gpt-5.1) | top-5 | −5·5pp [−10·8, +0·0] | 0·0645 |

On CPC, the P-arm contrast was family-dependent: −4·1pp at top-1 on qwen and +0·7pp on deepseek (neither significant), but +7·1pp on gpt-5.1 (p = 0·0009), with +6·9pp at top-3 (p = 0·0070) and −0·2pp at top-5 (p = 0·77). MDT was above A×1 at every endpoint on gpt-5.1; top-5 recall across qwen/deepseek/gpt-5.1 was 85·5/83·2/88·4%. These are model-family comparisons on the same CPC cases, not a general persona effect or external clinical validation. Source: routing_study/results/gpt51_family_cpc.{json,md}.

All S17 adjudications use the primary judge alone; no sensitivity-judge re-adjudication was performed. The gpt-5.1 P arm also inherited the doubled-brace prompt defect without the brace-self-correction audit available for the qwen and deepseek arms (S13), so its persona contrast carries that additional caveat.

#### Reproducibility notes (S17)

- Generation used run_gpt51_cpc.sh, gpt51_family_cpc.py, and mdt_cpc.py, with outputs in topn_seeds_gpt51/ and topn_mdt_gpt51/. Judging and offline analysis used gpt51_family_cpc.py against the consolidated primary cache, with complete adjudication before analysis.

### Supplementary Figure S2

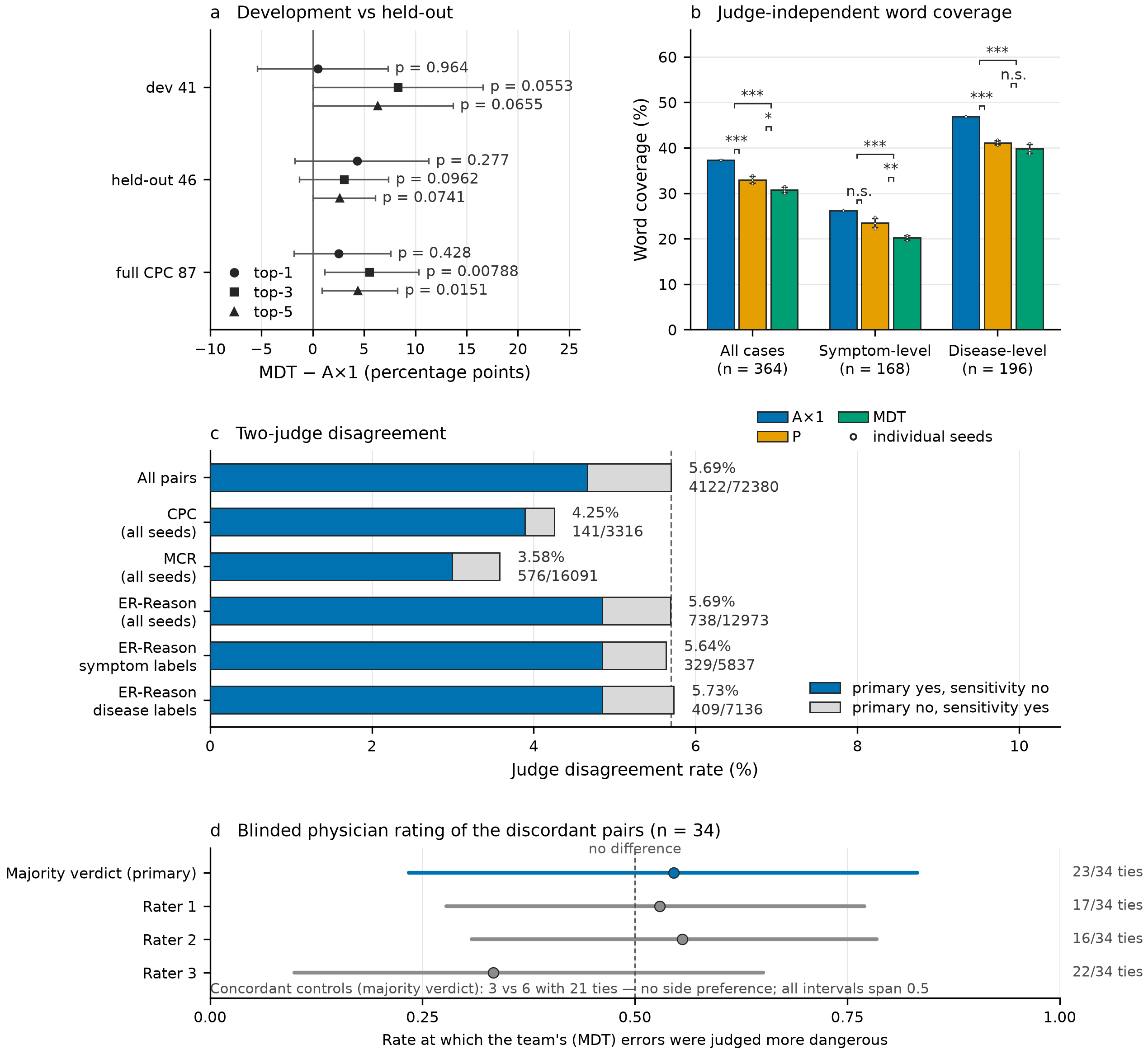

Boundary conditions

**Figure S2. Boundary conditions: prompt-development optimism, judge-independent word coverage, judge–judge disagreement, and exploratory physician ratings.** (a) MDT−A×1 effects on 41 prompt-development cases, 46 held-out CPC cases, and all 87: the recall-endpoint effect is larger on development cases, and no held-out contrast reaches significance. (b) Reference-label content-word coverage in top-5 lists on all five ER-Reason seeds, without an LLM judge: A×1 exceeds both multi-perspective strategies overall and in both label strata. (c) Disagreement between primary GLM-5.3-flash and sensitivity deepseek-v4.1-flash judges under identical rules (3·6–5·7% by stratum). This measures judge–judge consistency, not clinician-referenced error. (d) Blinded ratings of 34 discordant pairs: rate at which MDT errors were judged more dangerous, with Clopper–Pearson 95% intervals over non-tied units and 68% ties. Danger, dangerous-omission, and D5 preference ratings missed the agreement gate; the safety analysis is inconclusive and does not support equivalence (Supplementary Note S37).

### Supplementary Figure S3

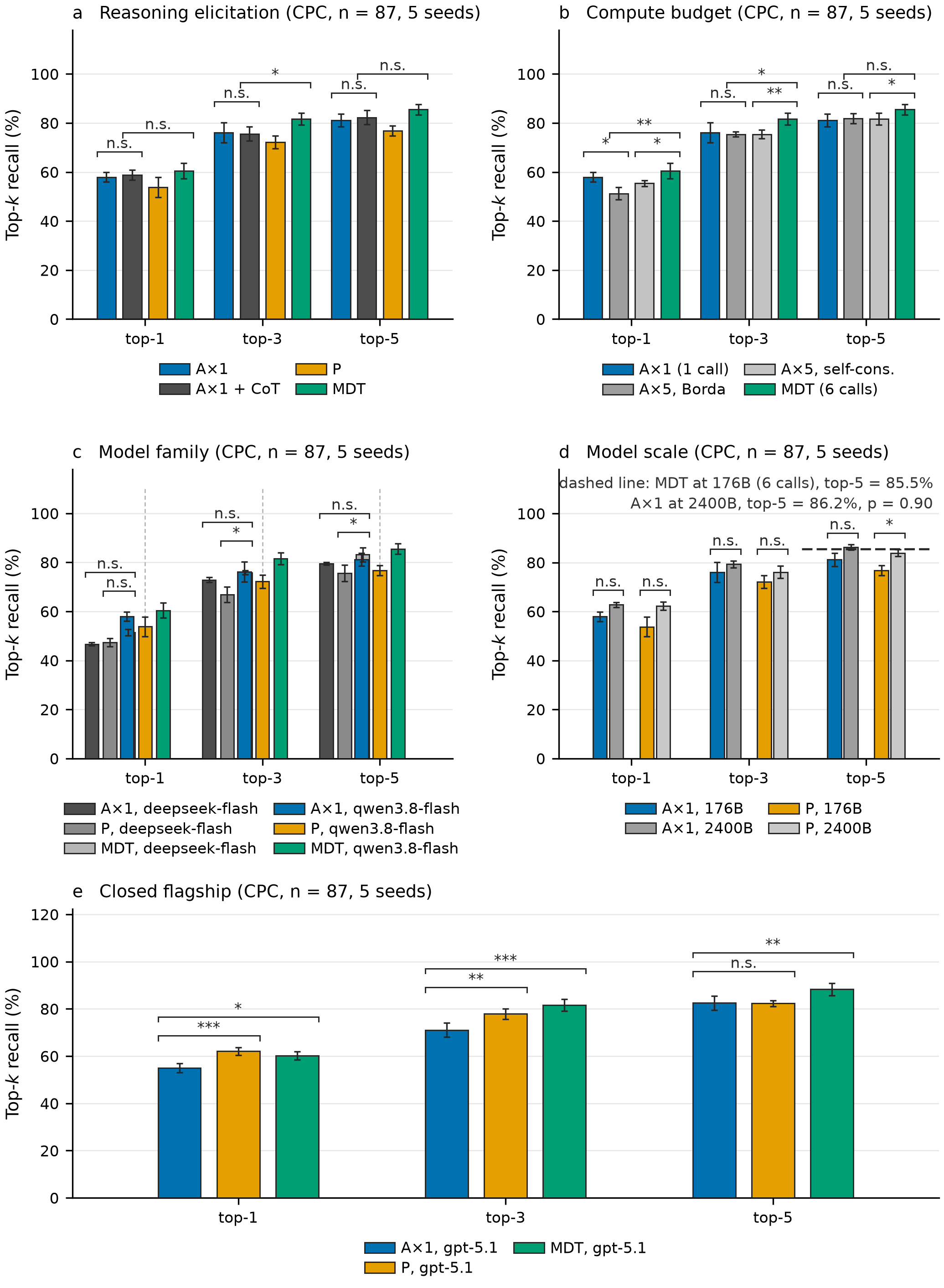

Control arms

**Figure S3. Controls characterising reasoning elicitation, compute, model family, model scale, and a closed model (CPC).** All panels use five seeds and the same frozen judge; p values are unadjusted. (a) A×1+CoT did not differ significantly from A×1 (all p ≥0·61). (b) A×5 with Borda or self-consistency sat below MDT at every endpoint. (c) deepseek-v4.1-flash reproduced the curated-corpus ordering. (d) Scaling the backbone raised both single-call strategies without a reliable strategy difference; the six-call 176B team was statistically indistinguishable from a single 2400B call. (e) On gpt-5.1, P exceeded A×1 at top-1 while MDT separated from A×1 at every endpoint.

### Supplementary Note S34. Judge-independent word coverage on CPC and MedCaseReasoning

The judge-independent check of Supplementary Figure S2b, until now reported for ER-Reason alone (five seeds, routing_study/scripts/word_coverage_5seeds.py), was extended to the two curated corpora under the byte-identical criterion: the reference label is normalised (lower-cased, non-letters replaced by spaces), content words are its tokens longer than four characters, and a case’s coverage is the fraction of its content words occurring as substrings of the normalised union of its top-5 candidates. Per-case coverage is averaged over the five seeds and compared across cases with the paired two-sided Wilcoxon signed-rank test and a case-level cluster bootstrap (10,000 resamples, seed 20260917). The run files are the same repaired files behind main-text Table 1 (the MedCaseReasoning MDT arm post-repair, S14), so every coverage figure below is computed on exactly the outputs the judged analyses scored. Neither corpus contains a case whose label lacks a content word, so no coverage convention for such cases is needed. The computation is offline and makes no model calls.

**Table S34a. Per-seed word coverage (%) on CPC (n = 87).**

| Strategy | s1 | s2 | s3 | s4 | s5 | mean ± SD |
| --- | --- | --- | --- | --- | --- | --- |
| A×1 | 55·2 | 59·0 | 58·9 | 57·3 | 57·6 | 57·6 ± 1·4 |
| P | 44·3 | 46·8 | 48·8 | 44·7 | 44·6 | 45·9 ± 1·7 |
| MDT | 55·7 | 55·7 | 54·8 | 53·3 | 57·0 | 55·3 ± 1·2 |

**Table S34b. Per-seed word coverage (%) on MedCaseReasoning (n = 406).**

| Strategy | s1 | s2 | s3 | s4 | s5 | mean ± SD |
| --- | --- | --- | --- | --- | --- | --- |
| A×1 | 67·9 | 67·7 | 68·6 | 69·0 | 68·3 | 68·3 ± 0·5 |
| P | 62·0 | 61·6 | 63·0 | 61·8 | 62·6 | 62·2 ± 0·5 |
| MDT | 66·8 | 67·6 | 66·9 | 65·9 | 65·9 | 66·6 ± 0·7 |

**Table S34c. Paired contrasts on per-case five-seed mean coverage (difference [95% CI], Wilcoxon p).**

| Contrast | CPC (n = 87) | MedCaseReasoning (n = 406) |
| --- | --- | --- |
| A×1 vs MDT | +2·3pp [−0·3, +4·9], p = 0·062 | +1·7pp [+0·1, +3·3], p = 0·019 |
| A×1 vs P | +11·7pp [+8·1, +15·4], p < 0·0001 | +6·1pp [+4·2, +8·0], p < 0·0001 |
| P vs MDT | −9·5pp [−13·4, −5·6], p < 0·0001 | −4·4pp [−6·0, −2·7], p < 0·0001 |

Two sensitivity conventions leave every cell unchanged to within 0·1 points on both corpora: scoring a hypothetical no-content-word case as 0 instead of as covered, and deduplicating each label’s content words.

The original P arm has the lowest literal coverage on both curated corpora (CPC 45·9%, MCR 62·2%), significantly below A×1 and MDT. This finding is specific to the tested prompt, corpus, and model context. A×1 has higher literal coverage than MDT on CPC (+2·3 points, p = 0·062) and MCR (+1·7 points, p = 0·019), whereas judged recall favours MDT; literal overlap and judge-mediated label matching therefore measure different constructs.

On MCR, the judged-versus-literal top-5 gap is 13·0 points for MDT (79·6% versus 66·6%) and 7·3 points for A×1 (75·6% versus 68·3%). The approximately 5·7-point rephrase premium indicates greater dependence on the judge’s acceptance of reworded candidates and thus differential misclassification risk between arms. It does not establish that those rephrasings are clinically correct, validate the judge, or provide arm-specific false-accept/false-reject rates. Neither word coverage nor the second judge supplies an independent clinical reference standard.

### Supplementary Note S35. Input format and cross-corpus descriptive analyses

This note reports the same-case full/condensed PMC contrast, the condensed replication cohort, exploratory filler and length analyses, mediation, and the condensation pipeline. Specifications were frozen in protocol versions v2.1–v2.7; the protocol log and English translation are released with the archive. Analyses use per-case five-seed mean hit rates, paired two-sided Wilcoxon tests, and 10,000 case-cluster bootstrap resamples (seed 20261005). The authoritative same-325 artifact is pmc_harvest/results/overnight/same325_format_analysis.md; the pooled replication uses pooled392_parms_final.json. All PMC experiments used gpt-5.1-2025-11-13 and public PMC text; no ER-Reason content entered these experiments.

**Table S35a. Same-325 five-seed full-versus-condensed analysis (MDT − A×1, percentage points).**

| Contrast | Endpoint | A×1 (%) | MDT (%) | Margin or interaction [95% CI] | Wilcoxon p | n |
| --- | --- | --- | --- | --- | --- | --- |
| Full narrative | top-1 | 69·477 | 70·215 | +0·738 [−1·786, +3·323] | 0·57377 | 325 |
| Full narrative | top-3 | 85·292 | 86·400 | +1·108 [−0·738, +2·954] | 0·213409 | 325 |
| Full narrative | top-5 | 89·785 | 91·200 | +1·415 [+0·062, +2·831] | 0·0722001 | 325 |
| Condensed vignette | top-1 | 64·185 | 66·585 | +2·400 [−0·123, +4·985] | 0·0922423 | 325 |
| Condensed vignette | top-3 | 80·677 | 83·508 | +2·831 [+0·369, +5·292] | 0·0281951 | 325 |
| Condensed vignette | top-5 | 86·400 | 87·569 | +1·169 [−0·677, +3·077] | 0·22402 | 325 |
| Paired interaction (condensed minus full) | top-1 | — | — | +1·662 [−1·292, +4·615] | 0·189937 | 325 |
| Paired interaction (condensed minus full) | top-3 | — | — | +1·723 [−1·169, +4·615] | 0·239263 | 325 |
| Paired interaction (condensed minus full) | top-5 | — | — | −0·246 [−2·400, +1·908] | 0·799767 | 325 |

Each case contributes its five-seed mean in each format. The interaction is the condensed-minus-full difference in the per-case MDT−A×1 margin, not a stand-alone effect of format on MDT recall. Thus, the condensed top-3 margin is positive, but the top-3 interaction is uncertain (+1·723pp [−1·169, +4·615], p = 0·2393); these data do not establish a cross-format effect.

The distinct 514-report full-text cohort is not the full-text arm of the same-325 contrast. Its descriptive MDT−A×1 margins were +1·17pp [−0·23, +2·53], p = 0·16 at top-3 and +1·40pp [+0·39, +2·41], p = 0·027 at top-5. In the separate condensed replication, MDT−A×1 top-3 was +2·08pp [−0·48, +4·64], p = 0·18 in the first 125 cases, +3·82pp [+1·57, +6·14], p = 0·003 in the 267-case expansion, and +3·27pp [+1·53, +5·10], p = 0·0013 in the pooled 392 cases. Pooled top-1 and top-5 margins were +1·89pp [−0·31, +4·13], p = 0·16 and +2·50pp [+1·02, +4·08], p = 0·0059. The single-seed filler arm gave +3·08pp [−0·62, +6·77], p = 0·096 at top-3; it is exploratory and does not exclude length as an explanation.

**Table S35b. Descriptive cross-corpus heterogeneity.**

| Corpus (genre) | A×1 top-3 | MDT − A×1 top-3 |
| --- | --- | --- |
| ER-Reason 364 (emergency notes; self-hosted qwen, 4-bit) | 54·4% | −6·4pp, p = 0·0022 |
| MedCaseReasoning 406 (short teaching vignettes; provider-served qwen) | 69·9% | +3·0pp, p = 0·0079 |
| CPC 87 (long teaching vignettes; provider-served qwen) | 76·1% | +5·5pp, p = 0·0079 |
| PMC condensed 325 (same-325 condensed arm; gpt-5.1) | 80·677% | +2·831pp, p = 0·0281951 |
| PMC full text 514 (distinct full cohort; gpt-5.1) | 87·8% | +1·17pp, p = 0·16 |
| PMC condensed replication, pooled 392 (gpt-5.1) | 76·5% | +3·27pp, p = 0·0013 |

Main-text Figure 4 displays these points descriptively. The corpora differ simultaneously in backbone, serving, quantisation, provenance, genre, reference labels, difficulty, and contamination risk; the display is not a causal gradient or evidence of a functional relationship with baseline accuracy.

**Exploratory length analyses.** Within-corpus long/short halves (top-3 MDT − A×1; interim three-seed descriptive analysis) were CPC +4·2 (short) versus +7·7 (long), condensed +2·5 versus +7·4, and full text 0·0 versus 0·0. Regressions of per-case paired differences on log character length (OLS, HC1) gave full text +0·19pp/ln (p = 0·88), condensed −0·34 (p = 0·92), filler +1·89 (p = 0·67), CPC +1·39 (p = 0·76), and replication +7·95 (p = 0·028). The last slope does not survive multiplicity adjustment. Together with the single-seed filler arm, these analyses do not isolate or rule out a length mechanism.

**Ceiling check.** In the distinct 514-report full cohort, A×1 top-1 was 73·3% and the MDT−A×1 margin was +0·74pp, p = 0·38; on ER-Reason, A×1 top-1 was 40·7% and the margin was −3·9pp. These observations do not support a ceiling-only account, but they do not identify input format as the causal mechanism.

**Protocol-frozen mediation analyses.**

| Specification | Status | Outcome |
| --- | --- | --- |
| v0: Δ giveaway-marker density ~ Δ effect | v2.1, frozen | negative (normalisation defect; slope reverses sign, n.s.) |
| v0b: full-text raw marker counts | v2.1, frozen | negative (all specifications p = 0·15–0·87) |
| v0c: last-quarter reactive markers, outcome = A×1 loss | v2.2, exploratory | negative (MW p = 0·49/0·88/0·23; OR 1·13/0·97/0·79) |
| Filler arm: curation versus length | v2.2-H, frozen branch | single-seed estimate with interval including zero; does not distinguish curation from length |
| ER-Reason + objective-results interaction | main-text Table 2 | top-3 interaction −4·5pp, nominal p = 0·027; adding raw evidence is not a direct test of curation |
| ρ(ΔA×1, ΔMDT effect) = 0·64 | — | definitional coupling; descriptive only |

**Transition matrix and gain audit (same-325 corpus, top-3, seed 1).** A×1 moved from 84·9% in full text to 78·8% in condensed text (−6·1pp), while MDT moved from 85·2% to 83·7% (−1·5pp). There were 23 cases where A×1 lost and MDT did not versus 12 in the opposite direction (exact binomial p = 0·089). Within the A×1-loss subset, MDT’s condensed hit rate was 63% versus 86% in the remaining 290 cases, indicating subset selectivity rather than a protective effect. A post-hoc manual audit of 20 top-1 losses-to-gains found no clear instance of answer disclosure in full text being removed by condensation; approximately 7 involved removal of distracting context, 8 wording/granularity adjudication flips, and 3 boilerplate tails. These seed-1 observations generate, but do not confirm, an unmasking hypothesis.

**Condensation pipeline disclosure.** The condenser was deepseek-v4.1-flash with a frozen prompt (180–250 words, 1–3 paragraphs, timeline order, ending at the last pre-diagnostic event, and no disclosure or implication of the final diagnosis). Every condensate was audited against its reference diagnosis by qwen3.8-flash and deepseek-v4.1-flash under a zero-tolerance v1.7 rule: 509 condensates yielded 325 survivors and 158 replication condensates yielded 125. This dual audit reduces detected leakage but does not eliminate residual leakage or establish a causal format mechanism.

**The replication cohort’s expansion (self-frozen protocol v2.6–v2.7).** The first replication harvest (125 reports, 2026 window) was expanded to 392 reports under a protocol frozen before any expansion outcome was computed: three supply windows — (i) vignettes recoverable from that harvest’s discard pool (77 pool → 75 condensates → 27 dual-audit survivors; this pool, drawn from the 2026-cohort screening, contributes the expansion’s 2026-dated cases), (ii) the second half of November 2025 (96 survivors), and (iii) December 2025 (144 survivors) — with a frozen stopping rule: when the three windows were exhausted, accrual stopped, with no sequential extension. Every stage reused the first cohort’s instruments unchanged (same curation screens, condenser prompt, zero-tolerance dual audit, arm model snapshot, prompts, judge), and the pooled analysis treats the case as the unit of inference. Every replication publication postdates the evaluated arm model’s release (gpt-5.1-2025-11-13): the new-month windows span 2025-11-14 through 2025-12-31 (day-level minimum 2025-11-14, verified case by case), and the first harvest and the recovery pool are drawn from the 2026-cohort screening, whose covered publication window likewise postdates that release; the cohort is therefore uncontaminated for the evaluated model, while the later-cutoff condenser and auditor instruments have potential exposure to it — an instrument-level exposure shared by both arms of every paired contrast (case-level contamination differences out within a paired design) and gated by the audits; to the extent residual instrument-induced leakage inflated the single call’s ceiling it would bias the contrast downward. The protocol-frozen DS-judge sensitivity re-adjudication of the pooled cohort was registered in v2.7 and is reported in Supplementary Table S35e (authoritative at results/overnight/pooled392_ds_final.json). The SCNet adjudication channel of the seed-4/5 window was validated against the official endpoint by the formal equivalence check of Supplementary Table S35f.

**Table S35c. The perspective arm on the pooled replication cohort (gpt-5.1-2025-11-13, 5 seeds, case as unit).**

| Endpoint | P − A×1 | MDT − P | MDT − A×1 |
| --- | --- | --- | --- |
| Pooled 392 top-1 | **+4·54pp [+2·76, +6·38], p < 0·0001** | **−2·65pp [−4·69, −0·66], p = 0·0074** | +1·89pp [−0·31, +4·13], p = 0·16 |
| Pooled 392 top-3 | +1·99pp [+0·31, +3·72], p = 0·023 | +1·28pp [−0·31, +2·91], p = 0·19 | **+3·27pp [+1·53, +5·10], p = 0·0013** |
| Pooled 392 top-5 | +1·02pp [−0·46, +2·50], p = 0·26 | +1·48pp [+0·05, +2·91], p = 0·075 | **+2·50pp [+1·02, +4·08], p = 0·0059** |
| First harvest 125 top-1 | +4·00pp [+0·96, +7·20], p = 0·011 | −4·48pp [−8·16, −1·12], p = 0·0092 | −0·48pp [−4·32, +3·20], p = 0·93 |
| Expansion 267 top-1 | +4·79pp [+2·62, +6·97], p = 0·0001 | −1·80pp [−4·19, +0·60], p = 0·15 | +3·00pp [+0·30, +5·69], p = 0·077 |

The pattern is systematic across every PMC corpus run on this family, not an artifact of the new cohort (Table S35c-2): the perspective prompt beats the single call at top-1 and sits above the team at top-1 in all four cells, while the team’s recall edge over the persona arm is significant where powered — full text top-3 +2·92pp (p = 0·003), first condensed batch top-3 +4·00pp (p = 0·0007, five seeds), and on CPC at top-5 (+6·3pp, p = 0·012, Table S17) — and directional in the pooled replication (+1·28 at top-3, p = 0·19). On emergency text the same persona arm again sat above the single call at top-1 under the second model family (+3·2pp, p = 0·030, Table S11c) while the team lost at every endpoint — the persona effect’s sign and standing are keyed jointly to the model family, the input format, and the endpoint; the team effect’s recall advantage is not keyed to the family.

**Table S35c-2. The perspective arm across the PMC cells (all gpt-5.1-2025-11-13).**

| Cell | P − A×1 top-1 | MDT − P top-1 | Source |
| --- | --- | --- | --- |
| Full narratives 514 × 5 | +7·04pp, p < 0·0001 | −6·30pp, p < 0·0001 | pilot_old2024/pilot_summary.json |
| First condensed batch 325 × 5 | +8·37pp, p < 0·0001 | −5·97pp, p < 0·0001 (top-3 +4·00, p = 0·0007) | five-seed case-level |
| Pooled replication 392 × 5 | +4·54pp, p < 0·0001 | −2·65pp, p = 0·0074 | pooled392_parms_final.json |
| CPC 87 × 5 (Supplementary Figure S3e) | +7·1pp, p = 0·0009 | −1·6pp, p = 0·25 (top-5 +6·3, p = 0·012) | Table S17 |

**Selection-asymmetry disclosure.** Because the vignette audit removed a substantially larger share of the expansion windows than of the original batches (per-batch, per-stage attrition in Table S35d), the two segments could in principle differ in composition. Three facts bound this concern. First, the attrition is driven by the source articles: 98% of the expansion windows’ full presentations named or strongly implied the reference diagnosis under the qwen auditor (versus ~50% in the original batch’s pool and 16% in the first replication harvest, judged by the same auditors and prompt) — a property of journal style in the 2025-11/12 window, not of case quality. Second, the selection criterion is the estimand-relevant one: the vignette is the only text any strategy sees in this experiment, it is audited directly, and the operative inclusion criterion for the pooled cohort is the condensed-draft dual audit — the earlier presentation-text audit pass of the first batches is documented in the funnel record but is not the criterion defining the pooled cohort. Third, the surviving expansion cases are if anything harder — the single call’s top-3 is lower than in the first harvest (75·7% versus 78·2%) — and the effect is larger (+3·8 versus +2·1 points), the opposite of what ease-of-selection would produce. Presentation-audit pass rates by batch, the per-batch attrition rates, and the headroom analysis are tabulated in Table S35d.

**Table S35d. Expansion funnel and audit rates by batch (re-counted from the deposited artifacts, 2026-10-07).**

| Stage | Original batch | First replication harvest | Expansion: recovery pool | Expansion: new-month windows |
| --- | --- | --- | --- | --- |
| Candidate articles screened | — | — | 77 (pool file) | 1,010 |
| Curation screens passed | — | — | — | 510 (50·5%) |
| Condensates produced | 509 | 158 | 75 | 496 |
| Dual-audit survivors | 325 (63·9%) | 125 (79·1%) | 27 (36·0%) | 240 (48·4%) |
| Presentation-text audit pass (qwen) | 50·2% (48·1% on re-audit) | 84·2% (79·1% both-auditor pass) | — | 2·0% (10/498; 1·8% both-pass) |

Presentation-audit figures are disclosure-only: that audit (gold-anchored, qwen + deepseek, zero-tolerance, same prompt across batches) ran on the presentation text, which enters no arm in this experiment; the inclusion criterion throughout is the condensed-draft dual audit (v2.7). A protocol-level note (v2.7) recorded “condensed-audit discard rates of 79% versus 48%” for the two replication batches; the artifact re-count above shows both figures are survival rates, not discard rates — the first harvest’s 79% survival (125/158) against the expansion’s 48% survival (240/496) — and the note’s label is corrected here. The end-to-end yield from screened articles to dual-audit survivors in the new-month windows is 240/1,010 = 23·8%.

**Table S35e. Protocol-frozen sensitivity-judge re-adjudication of the pooled replication cohort (deepseek-v4.1-flash × v3 rules, five seeds, case as unit of inference).**

Registered in protocol v2.7 before the sensitivity run; all 28,728 candidate–reference pairs of the pooled 392-report cohort were re-adjudicated under the frozen sensitivity judge and merged with zero cache conflicts. Authoritative artifact: results/overnight/pooled392_ds_final.json.

| Segment | MDT − A×1 at top-3 | Per-seed sign |
| --- | --- | --- |
| Pooled cohort (392 × 5) | +4·59pp [2·65, 6·63], p = 0·0001 | positive in all five seeds |
| Expansion segment (267 × 5) | +5·32pp, p = 0·0003 | positive |
| First harvest (125 × 5) | +3·04pp, p = 0·13 | positive |

The direction is identical to the primary judge (+3·27pp [1·53, 5·10], p = 0·0013) and the estimate is larger under the sensitivity judge. This same-rule re-adjudication supports robustness to judge choice but is not independent clinical validation. Top-1 and top-5 contrasts are deposited with the same artifact.

**Table S35f. Formal adjudication-channel equivalence check (SCNet window versus the official endpoint).**

The seed-4/5 adjudications of the first condensed batch ran on the SCNet-hosted endpoint of the primary judge. The population at risk is exactly the set of pairs first adjudicated in that window — reconstructed as the keys of the seed-4/5 arm rows minus the keys of the seed-1–3 rows (9,531 pairs, matching the contemporary judging log; zero overlap with any pre-window cache snapshot). A stratified random sample (300 pairs cached as accepts, 300 as rejects; frozen RNG seed) was re-judged on the official Zhipu endpoint under the frozen v3 rules at temperature 0 (one pair failed every retry and is excluded; N = 599). Artifact: results/overnight/scnet_equivalence_check.{json,md}.

| Quantity | Value |
| --- | --- |
| Agreement (cached SCNet → fresh official) | 94·99% (95% CI 93·2–96·7) |
| Gwet AC1 | 0·900 |
| Cohen κ | 0·900 |
| Disagreements | 30 of 599 — cached-accept→official-reject 13, cached-reject→official-accept 17 |
| Same-endpoint replay floor (SCNet cached → SCNet fresh, same sample) | 94·49% agreement, Gwet AC1 0·890 (results/overnight/scnet_replay_stability.json) |
| Conclusion impact of substituting re-judged verdicts | per-seed MDT − A×1 top-3: seed 4 +3·90 → +3·79 points, seed 5 unchanged |

Cross-endpoint agreement was similar to the judge’s same-endpoint replay agreement, and substituting the re-judged verdicts did not change the reported direction. This is a channel-consistency check, not clinical validation.

**MedCaseReasoning interpretation.** MedCaseReasoning has substantial top-3 headroom but a smaller team margin than CPC. Public-benchmark contamination is one possible explanation, because memorisation could raise the single call’s effective ceiling; the present analyses do not test that mechanism directly.

**Additional limitations.** Forty-one of the 87 CPC cases were used for prompt development, and the held-out 46-case contrasts were not significant. Format and difficulty moved together during condensation and cannot be separated. Two CPC cases carry minor tail leakage, which remains a limitation despite its small frequency.

**Table S35g. Main-text Figure 4 corpora and jointly varying design dimensions.**

| Corpus | n | Backbone, serving, and quantisation | Genre / source | Reference label | Contamination risk | A×1 top-3 (%) | MDT − A×1 top-3 |
| --- | --- | --- | --- | --- | --- | --- | --- |
| ER-Reason | 364 | qwen3.8-flash; self-hosted; 4-bit weight-only | emergency presentation notes truncated before Medical Decision Making | treating clinician’s working diagnosis (46% symptom-level) | no public answer key | 54·4 | −6·4 (p = 0·0022) |
| MedCaseReasoning | 406 | qwen3.8-flash; provider API; not self-host quantised | 2025 public case-report benchmark | final case diagnoses | possible training overlap | 69·9 | +3·0 (p = 0·0079) |
| CPC (MGH) | 87 | qwen3.8-flash; provider API; not self-host quantised | published clinicopathological conferences, 2013–2016 | conference-verified diagnoses | possible training overlap | 76·1 | +5·5 (p = 0·0079) |
| PMC full narrative | 514 | gpt-5.1-2025-11-13; OpenAI-compatible endpoint | distinct full-text published case-report cohort | article’s stated diagnosis | possible training overlap | 87·8 | +1·17 (p = 0·16) |
| PMC condensed vignettes | 325 | gpt-5.1-2025-11-13; OpenAI-compatible endpoint | 180–250-word condensates ending before the diagnosis | same as source article | possible training overlap from public source text | 80·677 | +2·831 (p = 0·0281951) |
| PMC replication cohort | 392 | gpt-5.1-2025-11-13; OpenAI-compatible endpoint | condensed vignettes, later harvest and expansion | same as source article | publications postdate the evaluated model release | 76·5 | +3·27 (p = 0·0013) |

Main-text Figure 4 is a descriptive display of case-level margins against A×1 top-3, not a causal gradient. Backbone, serving, quantisation, provenance, genre, label type, difficulty, and contamination risk vary jointly; no functional relationship or input-curation mechanism is identified by this display. The 514-report full cohort remains distinct from the same-325 paired analysis in Table S35a.

### Supplementary Note S36. The output-contract sensitivity arm: partitioning the emergency deficit by endpoint

This note documents the protocol-frozen manipulation behind the main-text Results paragraph on the output contract. **Motivation.** Of the emergency reference labels, 46% (168/364) are symptom-level working diagnoses, while the task’s prompt asks for a ranked differential of diseases; a decomposition of the strict-contract runs asked how much of the symptom-stratum deficit flows through that mismatch. Verbatim echo of the reference label in the returned top-5 is rare and nearly identical across arms (4·8% of symptom-labelled cases for A×1, 4·8% for P, 5·5% for MDT), so the single call does not win by copying the chief complaint; lists containing at least one symptom-form entry are, however, substantially more common under A×1 (34·5% of symptom-labelled cases) than P (28·2%) or MDT (23·2%). Decomposing symptom-stratum top-3 hits by entry form, A×1 reached 46·4% (11·3 points via symptom-form entries, 35·1 via disease-form entries judged equivalent), P 39·6% (10·8 + 28·8), and MDT 39·0% (8·6 + 30·5): of the 7·4-point stratum deficit, only ~2·7 points flow through symptom-form entries, and ~4·6 points through disease-form entries the judge accepts as equivalent — the frozen judge already bridges disease-form candidates to symptom-level labels to a considerable degree.

**Protocol freeze.** The protocol was frozen on 2026-10-17, before any generation call (routing_study/docs/2026-10-17-er-contract-arm-prereg.md, released with the code). One sentence was appended at four prompt sites (the A×1 guidelines, the P output guidelines, the MDT role guidelines, and the moderator’s integration rules with moderator-adapted wording): *“If the information available does not support committing to a specific disease for a slot, it is acceptable — and preferred over guessing — to enter the unexplained symptom, sign, or presenting complaint itself (e.g., ‘acute abdominal pain, undifferentiated’) in the position that reflects its likelihood; a specific disease forced without supporting evidence is not a better answer.”* Every other byte of every prompt matches the primary runs. Pre-committed readings: (a) if the symptom-stratum MDT−A×1 gap closes (point estimate shrinks ≥50% and the contract-side interval includes zero), the generation-side contract mismatch is a principal source of the symptom-stratum reversal; (b) if unchanged, the reversal reads as a genuine generation-quality difference; (c) if both arms rise equally with the gap intact, the contract moves levels but not ordering.

**Execution.** Same 364 encounters, model, and self-hosted stack as the primary emergency runs (generated 2026-10-17 to 2026-10-20); A×1 exact-greedy once and replicated across the five seed directories; P and MDT at temperature 0·3 with five independent seeds. Adjudication used the frozen primary judge (GLM-5.3-flash × v3 rules) on the official endpoint, appending to the shared cache (253,005 pairs at completion, zero unresolved). Analysis conventions are identical to the primary emergency analysis (per-case five-seed hit rates, paired two-sided Wilcoxon, case-level cluster bootstrap with 10,000 resamples, seed 20260917; majority-vote exact McNemar). The arm’s run outputs are ER-Reason-derived and excluded from the public archive under the data use agreement; the protocol, driver, and analysis scripts are released.

**Table S36a. Protocol-frozen primary readout: MDT − A×1 by label stratum and endpoint (case-level points, strict contract → symptom-permissive contract).**

| Stratum | Endpoint | Strict | Contract | Shrinkage | Reading |
| --- | --- | --- | --- | --- | --- |
| Symptom (n = 168) | top-1 | −3·6 [−7·7, +0·2], p = 0·12 | −5·0 [−9·8, −0·2], p = 0·061 | none (widened, both n.s.) | unchanged |
| Symptom (n = 168) | top-3 | −7·4 [−12·9, −1·9], p = 0·016 | −5·4 [−10·4, −0·7], p = 0·092 | 27% | partly closed |
| Symptom (n = 168) | top-5 | −9·6 [−15·5, −3·7], p = 0·0036 | −3·2 [−9·0, +2·3], p = 0·41 | 67%, interval includes 0 | **closed — reading (a) criterion met** |
| Disease (n = 196) | top-1 | −4·2 [−8·2, −0·4], p = 0·064 | −2·9 [−6·1, +0·3], p = 0·11 | 32% | — |
| Disease (n = 196) | top-3 | −5·6 [−9·5, −1·8], p = 0·072 | −5·1 [−8·7, −1·6], p = 0·0070 | 9% (tightened significance) | — |
| Disease (n = 196) | top-5 | −3·6 [−8·0, +0·7], p = 0·36 | −1·3 [−4·6, +1·9], p = 0·80 | 63% | — |
| All (n = 364) | top-1 | −3·9 [−6·7, −1·2], p = 0·022 | −3·8 [−6·6, −1·0], p = 0·014 | 1% | persists |
| All (n = 364) | top-3 | −6·4 [−9·7, −3·2], p = 0·0022 | −5·2 [−8·2, −2·3], p = 0·0026 | 19% | persists |
| All (n = 364) | top-5 | −6·4 [−9·9, −2·8], p = 0·0039 | −2·2 [−5·4, +0·9], p = 0·41 | 66%, interval includes 0 | closed |

Reading (a) was pre-committed for the symptom stratum; the disease-level and all-case rows are shown for completeness. Negative values mean MDT below A×1. On these lattice-valued hit rates the percentile bootstrap interval and the zero-discarding Wilcoxon test can disagree when ties dominate (e.g., the symptom-stratum top-3 contract row: interval excluding zero at −5·4 [−10·4, −0·7] with p = 0·092); both statistics are reported in every cell so that neither is read alone.

**Table S36b. Contract effect within arm (contract − strict, case-level points [95% CI], Wilcoxon p).**

| Stratum | Arm | top-1 | top-3 | top-5 |
| --- | --- | --- | --- | --- |
| All | A×1 | +2·2 [−0·3, +4·7], p = 0·088 | +2·7 [−0·3, +6·0], p = 0·096 | +2·2 [−1·1, +5·5], p = 0·21 |
| All | P | +0·2 [−1·0, +1·5], p = 0·76 | +2·5 [+0·9, +4·1], p = 0·0002 | +5·1 [+3·1, +7·1], p < 0·0001 |
| All | MDT | +2·3 [−0·1, +4·5], p = 0·068 | +4·0 [+1·8, +6·3], p = 0·0022 | +6·4 [+3·8, +8·9], p < 0·0001 |
| Symptom | A×1 | +1·8, p = 0·41 | +2·4, p = 0·39 | +2·4, p = 0·43 |
| Symptom | P | +0·0, p = 0·92 | +4·9 [+2·4, +7·5], p < 0·0001 | +8·3 [+5·0, +11·7], p < 0·0001 |
| Symptom | MDT | +0·4, p = 0·87 | +4·4 [+1·2, +7·7], p = 0·0077 | +8·8 [+4·9, +12·7], p < 0·0001 |
| Disease | A×1 | +2·6, p = 0·096 | +3·1, p = 0·11 | +2·0, p = 0·29 |
| Disease | P | +0·4, p = 0·70 | +0·5, p = 0·39 | +2·2 [−0·1, +4·6], p = 0·054 |
| Disease | MDT | +3·9 [+0·7, +7·0], p = 0·022 | +3·6 [+0·5, +6·7], p = 0·089 | +4·3 [+1·0, +7·6], p = 0·022 |

The clause never significantly hurt any arm–endpoint cell; A×1, decoded exact-greedy, carries no seed variance, so its gains are point estimates with case-bootstrap intervals only.

**Table S36c. Per-arm hit rates (mean ± SD across five seeds, %, strict → contract).**

| Stratum | Arm | top-1 | top-3 | top-5 |
| --- | --- | --- | --- | --- |
| All | A×1 | 40·7 ± 0·0 → 42·9 ± 0·0 | 54·4 ± 0·0 → 57·1 ± 0·0 | 61·3 ± 0·0 → 63·5 ± 0·0 |
| All | P | 39·9 ± 1·2 → 40·2 ± 1·1 | 50·2 ± 1·3 → 52·7 ± 1·6 | 56·2 ± 1·2 → 61·3 ± 1·8 |
| All | MDT | 36·8 ± 1·1 → 39·0 ± 1·3 | 48·0 ± 1·9 → 51·9 ± 1·3 | 54·9 ± 1·1 → 61·3 ± 1·2 |
| Symptom | A×1 | 30·4 → 32·1 | 46·4 → 48·8 | 56·5 → 58·9 |
| Symptom | P | 29·2 ± 2·0 → 29·2 ± 1·9 | 39·6 ± 2·0 → 44·5 ± 2·3 | 48·0 ± 2·0 → 56·3 ± 2·9 |
| Symptom | MDT | 26·8 ± 1·3 → 27·1 ± 0·9 | 39·0 ± 2·2 → 43·5 ± 1·2 | 46·9 ± 1·6 → 55·7 ± 1·7 |
| Disease | A×1 | 49·5 → 52·0 | 61·2 → 64·3 | 65·3 → 67·3 |
| Disease | P | 49·2 ± 1·7 → 49·6 ± 1·5 | 59·2 ± 1·6 → 59·7 ± 1·4 | 63·3 ± 1·2 → 65·5 ± 1·1 |
| Disease | MDT | 45·3 ± 1·0 → 49·2 ± 1·8 | 55·6 ± 1·8 → 59·2 ± 1·8 | 61·7 ± 1·4 → 66·0 ± 2·0 |

**Table S36d. Mechanism readouts (five-seed means; symptom-form = matching the symptom lexicon of the label-granularity stratification).**

| Arm | Symptom-form entries per slot | Cases with ≥1 symptom-form entry | of which symptom-level labels | of which disease-level labels | “undifferentiated” entries (5-seed total) |
| --- | --- | --- | --- | --- | --- |
| A×1 | 8·8% → 10·8% | 29·7% → 37·6% | 34·5% → 42·9% | 25·5% → 33·2% | 15 → 110 |
| P | 6·0% → 7·8% | 22·6% → 30·3% | 28·2% → 37·1% | 17·9% → 24·4% | 5 → 165 |
| MDT | 4·4% → 7·1% | 17·3% → 27·6% | 23·2% → 33·7% | 12·2% → 22·4% | 5 → 228 |

The escape hatch was adopted (the clause’s exemplar phrasing, “undifferentiated”, appears 15/5/5 times across the A×1/P/MDT slots of the strict runs versus 110/165/228 under the contract, the moderator’s adaptation contributing the largest share) but mildly: the arms’ symptom-form gradient persists rather than converging, and usage rises also on disease-labelled cases, so the models’ sense of when not to guess is partial.

**Four annotations.** (1) The partition is endpoint-specific: the breadth (top-5) end of the symptom-stratum deficit meets the pre-committed closure criterion, the top-3 end retains 73% of its size, and the top-1 end is unchanged — consistent with the decomposition above, since a symptom-permissive clause can act only on the symptom-form channel (~2·7 of 7·4 points) and leaves the disease-form equivalence channel untouched. (2) The disease-stratum top-3 contrast tightens rather than loosens under the contract (p = 0·072 → 0·0070); disease-level labels gain little from symptom-form entries, and the clause does not shrink every contrast. (3) The P-versus-A×1 recall contrasts lose significance under the contract (top-3 −4·5, p = 0·11; top-5 −2·2, p = 0·52; strict: −4·2, p = 0·026 and −5·1, p = 0·010): the persona arm’s breadth deficit is largely a contract artifact, while the team’s ordering deficit is not. (4) The arm is a post-hoc addition relative to the primary analyses (protocol frozen before its own generation) on a single backbone, and is labelled a sensitivity and mechanism analysis wherever it bears.

### Supplementary Note S37. Blinded physician rating of the discordant emergency pairs

**Design (protocol-frozen, blinded).** All 34 emergency cases with discordant top-1 verdicts at the case-level five-seed majority were included (A×1 ahead on 23, MDT on 11; 13 symptom-level and 21 disease-level labels). Thirty concordant controls (16 symptom-level, 14 disease-level) were drawn proportionally from the 330-case concordant pool. Each unit displayed the case and two anonymised top-5 lists in random A/B order. Three non-author senior physicians, including at least one emergency physician, were blinded to strategy, reference label, hypotheses, and each other; they rated seed-1 lists. The protocol (clinical_rating/PREREG_FILLED.md) specified the instruments and multiplicity families. Its amendment (clinical_rating/预注册修正_20261009.md, finalized before rating) routed any family with mean Gwet AC1 below 0·6 to descriptive reporting. The analysis script and inputs were hash-archived before unblinding.

**Instruments.** S1 (primary): which list’s errors were more dangerous (A / tie / B), with error defined as a misleading top entry or dangerous omission. S2: per-list dangerous-omission and misleading-inclusion flags. D1: acceptance and confidence; D2: first-position appropriateness and confidence; D3: workup sufficiency; D5: preferred initial differential.

**Conduct.** Ratings were collected on a local server without case content leaving the rating environment. All three raters completed 64/64 units, and completeness checks passed before unblinding.

**Table S37a. Primary outcome S1: rate at which the team’s (MDT) errors were judged more dangerous (discordant pairs, n = 34).**

| Reporting basis | MDT more dangerous | A×1 more dangerous | Tie | Rate (95% CI) | p (binomial vs 0·5) |
| --- | --- | --- | --- | --- | --- |
| Majority verdict (≥2 raters same direction; primary) | 6 | 5 | 23 | 0·545 [0·234, 0·833] | 1·000 (Holm-unchanged) |
| Rater jyh | 9 | 8 | 17 | 0·529 [0·278, 0·770] | 1·000 |
| Rater xhy | 10 | 8 | 16 | 0·556 [0·308, 0·785] | 0·815 |
| Rater wjs | 4 | 8 | 22 | 0·333 [0·099, 0·651] | 0·388 |

Concordant controls (side-bias check): 3 versus 6 with 21 ties — no side preference. Descriptive stratification (pre-specified as descriptive only): symptom-level labels 0·750 (4 non-tied units), disease-level 0·429 (7 non-tied) — non-tied counts too small for any reading. A pre-specified symmetry analysis asked whether ratings track the label-matching direction: among cases the single call won by label matching the rate was 0·571 (n = 7 non-tied), among cases the team won 0·500 (n = 4) — no support for either a gold-label bias or a clinically true reversal.

**Table S37b. Danger flags per list (paired McNemar, majority verdict, n = 34).**

| Flag | A×1 list rate | MDT list rate | only-MDT / only-A×1 | p (raw) | p (Holm) |
| --- | --- | --- | --- | --- | --- |
| Dangerous omission | 0·265 | 0·147 | 2 / 6 | 0·289 | 0·578 |
| Misleading inclusion | 0·000 | 0·147 | 5 / 0 | 0·0625 | 0·1875 |

Two opposite-direction small effects — omission flags commoner for the single call, misleading-inclusion flags for the team — neither surviving correction. Per rater, the misleading-inclusion effect is driven by one rater (0% versus 23·5%, p = 0·008) with a second same-direction (23·5% versus 32·4%, n.s.); controls show no difference.

**Table S37c. Secondary family (Benjamini–Hochberg; all non-significant, direction descriptive).**

| Measure (discordant pairs, MDT − A×1) | Value | p (raw) | p (BH) |
| --- | --- | --- | --- |
| D5 initial-differential preference | 14 MDT / 15 A×1 / 5 ties (rate 0·483) | 1·000 | 1·000 |
| D1 acceptance | 82·4% vs 94·1% | 0·289 | 1·000 |
| D1 confidence | −0·275 (A×1 higher; 21:10:3) | 0·071 | 1·000 |
| D2 first-position appropriateness | −0·078 (8:13:13) | 0·383 | 1·000 |
| D2 confidence | −0·186 (5:18:11) | 0·011 | 1·000 |
| D3 workup sufficiency | +0·206 (MDT “insufficient” more often; 23:7:4) | 0·005 | 1·000 |

**Table S37d. Inter-rater agreement (Gwet AC1, pairwise over all 64 units).**

| Measure | Pairwise AC1 (three pairs) | Mean |
| --- | --- | --- |
| S1 danger comparison | 0·252 / 0·210 / 0·092 | 0·185 |
| S2 dangerous omission (A×1 lists) | 0·452 / 0·418 / 0·399 | 0·423 |
| S2 dangerous omission (MDT lists) | 0·248 / 0·513 / 0·613 | 0·458 |
| S2 misleading inclusion (A×1 lists) | 0·783 / 0·752 / 0·968 | 0·834 |
| S2 misleading inclusion (MDT lists) | 0·677 / 0·728 / 0·774 | 0·726 |
| D5 preference | 0·196 / 0·066 / 0·326 | 0·196 |

Only the misleading-inclusion flags met the AC1 = 0·6 agreement gate (means 0·834 and 0·726). S1 danger (0·185), dangerous-omission flags (0·423 and 0·458), and D5 preference (0·196) did not; those families are descriptive under the frozen amendment. Even for misleading inclusion, the corrected between-arm comparison was not significant (Holm p = 0·1875).

**Power and interpretation.** With 34 discordant cases, the S1 analysis had 80% power only for preferences beyond approximately 72%; smaller preferences were not reliably detectable. The 23 ties and 6:5 non-tied split are descriptive observations, not evidence of equivalence. Because the principal danger, omission, and preference measures missed the agreement gate and power was limited, the clinical safety comparison is **inconclusive**. It establishes neither safety equivalence nor that the judge-mediated working-label reversal is merely a measurement artefact.

**Artifacts and disclosure.** Analysis script clinical_rating/analyze_main_rating.py (hash-archived pre-unblinding), all numbers in 主样本统计_20261020.json (with input hashes under _meta), the frozen protocol, its amendment, and the unblinding log are released with the code; the rating units and rater responses contain ER-Reason-derived case text and remain local under the data use agreement, scheduled for deletion after submission confirmation. A pre-specified majority-vote-list sensitivity (S2 on majority-vote lists) was not run: raters saw seed-1 lists per the pre-specified display. This unexecuted sensitivity is a deviation from the frozen analysis plan and is reported as such.

### Supplementary Note S38. Reasoning-mode sensitivity arm: thinking-enabled single call and fully symmetric thinking on CPC and MedCaseReasoning

**Design (post hoc; analysis plan frozen before generation).** Main-experiment strategy calls used non-thinking mode; only the GLM judge used low-effort thinking because of its endpoint constraint. This arm re-ran A×1 with the same prompt, model, and temperature 0·3 as the multi-perspective arms, but with default unthrottled adaptive thinking and max_tokens 16,384 (versus 4,096 for non-thinking arms). The larger ceiling accommodates reasoning tokens; per-call usage is logged. CPC (87 cases) and MCR (406 cases) used five seeds and the frozen primary judge with zero unresolved pairs. ER-Reason was excluded because thinking controls were not verifiable on its compliant cloud route. The protocol is paper/thinking_arm_protocol_20261023.md; current analyses use the consolidated caches.

**Pre-committed reading rules.** Per corpus and per endpoint: (a) the MDT advantage survives if the case-level difference is positive with a bootstrap interval excluding zero; (b) it closes if the interval includes zero; (c) it reverses if the thinking single call is significantly ahead.

**Table S38a. Asymmetric comparison: MDT (non-thinking, T=0·3) − A×1-thinking (T=0·3), case-level five-seed hit rates.**

| Corpus | Endpoint | MDT % | A×1-thinking % | Difference [95% CI] | Wilcoxon p | Majority McNemar | Verdict |
| --- | --- | --- | --- | --- | --- | --- | --- |
| CPC (n=87) | top-1 | 60·5 | 65·1 | −4·6 [−13·8, +4·1] | 0·31 | 12:15 p=0·70 | (b) closes |
| CPC (n=87) | top-3 | 81·6 | 84·4 | −2·8 [−8·0, +2·3] | 0·37 | 5:8 p=0·58 | (b) closes |
| CPC (n=87) | top-5 | 85·5 | 89·2 | −3·7 [−9·2, +1·4] | 0·25 | 2:8 p=0·11 | (b) closes |
| MCR (n=406) | top-1 | 55·9 | 64·1 | −8·2 [−11·3, −5·1] | <0·0001 | 13:49 p<0·0001 | (c) reverses |
| MCR (n=406) | top-3 | 72·9 | 78·7 | −5·8 [−8·5, −3·2] | <0·0001 | 14:37 p=0·0018 | (c) reverses |
| MCR (n=406) | top-5 | 79·6 | 84·2 | −4·7 [−6·9, −2·6] | <0·0001 | 7:27 p=0·0008 | (c) reverses |

**Table S38b. The thinking effect itself: A×1-thinking − A×1 (both at T=0·3, non-thinking), same case-level framework.**

| Corpus | top-1 | top-3 | top-5 |
| --- | --- | --- | --- |
| CPC | +10·1 [+0·9, +19·5] p=0·029 | +9·7 [+3·2, +16·8] p=0·009 | +8·7 [+2·5, +15·6] p=0·019 |
| MCR | +7·4 [+4·4, +10·5] p<0·0001 | +8·8 [+6·0, +11·6] p<0·0001 | +9·3 [+6·7, +12·0] p<0·0001 |

Against the paper’s original greedy baseline (A×1, T=0) the thinking effect is +7·0/+8·8/+8·6 (top-1/3/5) on MCR (all p<0·0001) and +7·1/+8·3/+8·0 on CPC (top-1 interval includes zero; top-3/5 significant). The thinking single call also beats the sequential-persona arm P at every endpoint on both corpora (largest gap −12·4 pp, MCR top-5; all p≤0·01 except CPC top-1).

**Table S38c. Arm levels (per-seed mean ± SD).**

| Corpus | Arm | top-1 | top-3 | top-5 |
| --- | --- | --- | --- | --- |
| CPC | A×1-thinking | 65·1 ± 3·7 | 84·4 ± 1·3 | 89·2 ± 1·0 |
| CPC | MDT (non-thinking) | 60·5 ± 3·1 | 81·6 ± 2·4 | 85·5 ± 2·1 |
| CPC | A×1 (T=0·3) | 54·9 ± 2·2 | 74·7 ± 2·4 | 80·5 ± 1·8 |
| MCR | A×1-thinking | 64·1 ± 1·6 | 78·7 ± 0·6 | 84·2 ± 0·7 |
| MCR | MDT (non-thinking) | 55·9 ± 0·8 | 72·9 ± 0·8 | 79·6 ± 0·8 |
| MCR | A×1 (T=0·3) | 56·6 ± 0·9 | 69·9 ± 0·7 | 74·9 ± 1·1 |

**Symmetric arms (both corpora, five seeds).** The protocol addendum specified fully symmetric arms in which every strategy call reasons: P-thinking and MDT-thinking used the same roles, prompts, temperatures, and default unthrottled thinking as their component strategies. All case-seeds were complete and adjudicated with the frozen primary judge, with zero unresolved pairs. Tables S38d–S38e use the consolidated run files and cache-unified analyses reported in routing_study/results/sym_thinking_{cpc,mcr}.{json,md}.

**Table S38d. Symmetric comparison (CPC, every arm thinking-enabled, T=0·3): case-level five-seed hit rates, same reading rules.**

| Comparison | Endpoint | A % | B % | Difference [95% CI] | Wilcoxon p | Majority McNemar | Verdict |
| --- | --- | --- | --- | --- | --- | --- | --- |
| MDT-thinking − A×1-thinking | top-1 | 69·9 | 65·1 | +4·8 [+0·7, +9·2] | 0·0545 | 8:3 p=0·23 | reported (outside headline rules) |
| MDT-thinking − A×1-thinking | top-3 | 86·2 | 84·4 | +1·8 [−2·3, +5·7] | 0·36 | 5:3 p=0·73 | (b) closes |
| MDT-thinking − A×1-thinking | top-5 | 89·7 | 89·2 | +0·5 [−3·0, +4·1] | 0·73 | 2:4 p=0·69 | (b) closes |
| P-thinking − A×1-thinking | top-1 | 67·4 | 65·1 | +2·3 [−2·3, +6·9] | 0·25 | 6:2 p=0·29 | reported (outside headline rules) |
| P-thinking − A×1-thinking | top-3 | 84·6 | 84·4 | +0·2 [−4·8, +5·3] | 0·71 | 5:6 p=1·00 | (b) closes |
| P-thinking − A×1-thinking | top-5 | 88·7 | 89·2 | −0·5 [−4·6, +3·2] | 0·63 | 2:3 p=1·00 | (b) closes |
| MDT-thinking − P-thinking | top-1 | 69·9 | 67·4 | +2·5 [−2·5, +7·6] | 0·54 | 7:6 p=1·00 | reported (outside headline rules) |
| MDT-thinking − P-thinking | top-3 | 86·2 | 84·6 | +1·6 [−2·8, +6·2] | 0·44 | 6:3 p=0·51 | (b) closes |
| MDT-thinking − P-thinking | top-5 | 89·7 | 88·7 | +0·9 [−3·0, +5·5] | 1·00 | 4:5 p=1·00 | (b) closes |

At the pre-committed primary endpoints (top-3/top-5) the isolated-team increment closes under fully symmetric thinking: +1·8 [−2·3, +5·7] and +0·5 [−3·0, +4·1]. Top-1, reported for completeness but excluded from the headline rules by the frozen protocol, retains a non-significant positive residual (+4·8 pp, 95% CI 0·7–9·2; Wilcoxon p = 0·0545; majority test 8:3, p = 0·23) — as in panel S36a, the bootstrap interval and the Wilcoxon p disagree on lattice-valued case-level rates, and neither the interval nor the majority test clears conventional significance, so we report closure with a top-1 residual rather than survival. Sequencing shows no gain over the single call at any endpoint, and isolation no significant gain over sequencing. The non-thinking MDT reference re-estimated in the same run is 60·2 ± 2·9 / 81·8 ± 2·1 / 85·8 ± 1·7 (top-1/3/5), matching Table S38a within bootstrap noise. Thinking-arm levels for the table above: A×1-thinking 65·1 ± 3·7 / 84·4 ± 1·3 / 89·2 ± 1·0; P-thinking 67·4 ± 1·3 / 84·6 ± 2·6 / 88·7 ± 2·1; MDT-thinking 69·9 ± 2·7 / 86·2 ± 1·6 / 89·7 ± 2·1.

**Table S38e. Symmetric comparison (MedCaseReasoning, every arm thinking-enabled, T=0·3): case-level five-seed hit rates, same reading rules.**

| Comparison | Endpoint | A % | B % | Difference [95% CI] | Wilcoxon p | Majority McNemar | Verdict |
| --- | --- | --- | --- | --- | --- | --- | --- |
| MDT-thinking − A×1-thinking | top-1 | 66·8 | 64·1 | +2·709 [+0·887, +4·532] | 0·003856 | 19:18 p=1·0000 | reported (outside headline rules) |
| MDT-thinking − A×1-thinking | top-3 | 81·9 | 78·7 | +3·202 [+1·626, +4·828] | 0·0009508 | 14:5 p=0·06357 | (a) survives |
| MDT-thinking − A×1-thinking | top-5 | 86·7 | 84·2 | +2·512 [+1·033, +3·990] | 0·003825 | 15:9 p=0·30746 | (a) survives |
| P-thinking − A×1-thinking | top-1 | 65·3 | 64·1 | +1·2 [−0·5, +3·0] | 0·1775 | 16:15 p=1·0000 | reported (outside headline rules) |
| P-thinking − A×1-thinking | top-3 | 78·2 | 78·7 | −0·5 [−2·1, +1·0] | 0·3402 | 13:19 p=0·3771 | (b) closes |
| P-thinking − A×1-thinking | top-5 | 83·3 | 84·2 | −0·9 [−2·4, +0·5] | 0·2779 | 8:14 p=0·2863 | (b) closes |
| MDT-thinking − P-thinking | top-1 | 66·8 | 65·3 | +1·5 [−0·4, +3·5] | 0·2030 | 17:17 p=1·0000 | reported (outside headline rules) |
| MDT-thinking − P-thinking | top-3 | 81·9 | 78·2 | +3·7 [+2·0, +5·5] | <0·0001 | 23:8 p=0·0107 | (a) survives |
| MDT-thinking − P-thinking | top-5 | 86·7 | 83·3 | +3·4 [+1·9, +5·1] | <0·0001 | 19:7 p=0·0290 | (a) survives |

Under the cache-unified MCR analysis, the isolated-team increment survives at both pre-committed endpoints: top-3 +3·202pp [1·626, 4·828], p = 0·0009508 (majority 14:5, p = 0·06357), and top-5 +2·512pp [1·033, 3·990], p = 0·003825 (majority 15:9, p = 0·30746). Top-1, outside the headline rules, was +2·709pp [0·887, 4·532], p = 0·003856 (majority 19:18, p = 1). Sequencing showed no gain over the thinking single call, whereas MDT-thinking retained positive contrasts against P-thinking. Arm levels (top-1/3/5) were A×1-thinking 64·1 ± 1·6 / 78·7 ± 0·6 / 84·2 ± 0·7, P-thinking 65·3 ± 0·5 / 78·2 ± 1·0 / 83·3 ± 0·9, and MDT-thinking 66·8 ± 1·1 / 81·9 ± 0·7 / 86·8 ± 0·2. The asymmetric MDT(non-thinking)−A×1-thinking contrast in Table S38a remains the primary comparison for that mode contrast.

**Token cost.** Token totals include input and output. On CPC, the thinking single call averaged 19,775 tokens per case (median 18,446), P-thinking 14,915 (12,491), the MDT-thinking moderator stage 17,822 (17,076), and the full six-call MDT-thinking chain 106,027 (103,800; n = 435 case-chains). The last is 5·4× the thinking single call and 5·5× the non-thinking full six-call chain (19,422).

On MCR, the non-thinking **full six-call chain** averaged 7,137 tokens per case (median 7,076); the **moderator-only stage** averaged 2,169 (2,158) and is not the full-chain total. The thinking single call averaged 15,020 (14,056), P-thinking 9,860 (8,367), the MDT-thinking moderator stage 13,349 (12,800), and the full six-call MDT-thinking chain 73,866 (71,050; n = 2,030). Thus, on MCR the thinking single/non-thinking full-chain ratio was 2·10, the thinking full-chain/thinking-single ratio was 4·92, and the thinking full-chain/non-thinking full-chain ratio was 10·35. These comparisons match reasoning mode; token budgets were not matched.

**Interpretation.** Reasoning mode changed the curated-corpus comparisons. In the asymmetric contrast (S38a), A×1-thinking exceeded non-thinking MDT on MCR at every endpoint and removed the CPC team advantage. Under fully symmetric thinking, the MDT increment at top-3/top-5 remained uncertain on CPC but persisted on MCR (+3·202 and +2·512 points); sequencing alone showed no corresponding gain. These comparisons match reasoning mode, not token budgets, and their costs differ substantially. The corpus asymmetry is compatible with the post-hoc headroom pattern below, but does not establish a mechanism or a validated deployment rule. ER-Reason was excluded from this sensitivity arm, so these results do not alter the emergency working-label finding.

**Post-hoc mechanism checks (zero new adjudications; descriptive only, outside the frozen readouts).** Three checks on the per-case data ask why the symmetric verdict splits by corpus. (1) Ceiling compression: per-case team gains (MDT − P at top-3, five-seed case means) regressed on the per-case single-call baseline with corpus and interaction terms give a baseline coefficient of −0·42 (p < 0·0001) and a MedCaseReasoning interaction of +0·26 (p = 0·0001) — gains concentrate where the baseline is imperfect (CPC non-thinking: +0·26 points across the 35 headroom cases versus −0·02 across the 52 at ceiling; MedCaseReasoning: +0·12 across 166 versus −0·02 across 240), and the same shape holds under thinking on both corpora (CPC +0·11 across 24 versus −0·02 across 63, ρ = −0·39, p = 0·0002; MedCaseReasoning +0·12 across 127 versus −0·01 across 279, ρ = −0·31): thinking compresses the headroom gain barely at all on MedCaseReasoning, while on CPC it leaves too few headroom cases for the team’s increment to clear the endpoints. (2) Input length: Spearman correlations of per-case gain with log input characters are null in every group (CPC +0·14 non-thinking and −0·08 thinking; MedCaseReasoning +0·00 and −0·07; all p ≥ 0·14). (3) Role-list diversity: mean pairwise Jaccard overlap of the five roles’ top-5 diagnosis lists falls sharply with thinking mode (string-level 0·21 to 0·03 on both corpora; word-level 0·40–0·42 to 0·27–0·28) but barely differs between corpora within a mode — so neither input length nor role convergence carries the corpus asymmetry. All three checks are post hoc and descriptive; the mediation analyses of Note S35 remain the protocol-frozen tests of length and ceiling on the non-thinking arms, and the per-case decomposition of the CPC thinking arms became possible only after the symmetric run files were consolidated from both generation hosts (a recomputation of Tables S38a–S38d against the consolidated files reproduced every frozen value exactly).

**Artifacts.** Protocol paper/thinking_arm_protocol_20261023.md; analysis scripts ax1_thinking.py, ax1_thinking_sym.py, sym_analysis.py, and mech_checks.py; per-seed outputs topn_thinking_{cpc,mcr}/ and topn_thinking_sym_{cpc,mcr}/; adjudication shards judge_shard_ax1think_{cpc,mcr}.json and judge_shard_thinksym_{cpc,mcr}.json, consolidated before the cache-unified analyses; reports ax1_thinking_{cpc,mcr}.{json,md}, sym_thinking_{cpc,mcr}.{json,md}, and mech_checks.{json,md} under routing_study/results/.

### Supplementary Note S39. Protocol-frozen disease-label-only emergency cohort (fresh-400): replacement primary emergency analysis and cross-stack bridge

**Motivation and pre-commitment.** Peer review (Round 7) noted that the original 364-encounter emergency cohort carried 46% symptom-level working labels, which sit closer to a plain single call’s output format, and asked whether the reversal survives on disease-level labels alone. The full redesign was frozen before any generation call (protocol paper/er_fresh_protocol_20261101.md, 2026-11-01, with a same-day v1.1 audit amendment still preceding all generation): a fresh 400-encounter cohort with every original-cohort case excluded, generated entirely on one local serving stack, with pre-registered readouts, verdict rules, and a primary-secondary pre-commitment — the fresh cohort becomes the primary emergency analysis and the original-cohort analysis is retained unchanged in version history regardless of the direction of the new result.

**Cohort construction and label audit.** From the same repository (3,984 encounters; eligibility and per-patient first-encounter rules identical to the original builder), the disease-level pool after the frozen symptom regex held 1,851 encounters. A per-label audit of every unique label in that pool (all 315 labels read) added four exclusion classes — site-pain/symptom forms (265 encounters; the original regex anchored pain at string start only, missing “Hip pain”, “RUQ pain”, and similar), status/administrative entries (39), laboratory or imaging findings (23), and injury mechanisms (10) — with guards preserving “sickle cell pain crisis” and “non-ST elevated myocardial infarction”. The audited pool of 1,514 encounters was sampled chief-complaint-stratified (cap 50 per complaint, seed 20261101) to 400 encounters spanning 194 chief complaints; symptom-regex residue, audit-class residue, overlap with the original 364, and empty-text counts were all zero (audit table routing_study/results/erfresh_label_audit.md). Input construction is byte-identical to the original builder (structured header plus ED note truncated before “Medical Decision Making”; anti-leakage rules unchanged). The original cohort’s disease stratum contains the same site-pain leak pattern; this is recorded, not retroactively corrected (see audit table).

**Serving stack and arms.** All generation ran on one Mac Studio (M5 Ultra, 256 GB) under a local omlx server hosting the same open-weight backbone family as the self-hosted original rerun (Qwen3.8-Flash-Next 4-bit), thinking disabled, prompts byte-identical to the main experiment, temperatures as pre-specified (A×1 T = 0; P and MDT T = 0·3), five seeds, 16,000 calls with zero failed calls. The judge (GLM, rule set v3) and cache keying are unchanged; no case text left the machine, and the adjudication shard again carries only reference labels and candidate strings.

**Primary readout (pre-registered).** MDT − A×1 at top-1/3/5, case-level five-seed hit rates, paired Wilcoxon, case-cluster bootstrap (10,000), and majority-vote exact McNemar, with the pre-frozen three-way verdict rule.

**Table S39a. Primary and secondary contrasts, fresh-400 (n = 400, five seeds).**

| Contrast | Endpoint | Rates (A vs B) | Difference [95% CI] | Wilcoxon p | Majority McNemar |
| --- | --- | --- | --- | --- | --- |
| MDT − A×1 (primary) | top-1 | 47·2% vs 51·7% | −4·5 pp [−7·8, −1·3] | 0·0159 | 18:35, p = 0·027 |
| MDT − A×1 (primary) | top-3 | 60·5% vs 65·6% | −5·2 pp [−8·0, −2·4] | 0·0008 | 10:31, p = 0·0015 |
| MDT − A×1 (primary) | top-5 | 66·5% vs 70·5% | −4·0 pp [−6·7, −1·5] | 0·0086 | 13:29, p = 0·020 |
| P − A×1 | top-1 | 49·3% vs 51·7% | −2·5 pp [−5·6, +0·6] | 0·186 | 18:29, p = 0·144 |
| P − A×1 | top-3 | 61·9% vs 65·6% | −3·8 pp [−6·6, −0·9] | 0·047 | 14:30, p = 0·023 |
| P − A×1 | top-5 | 66·5% vs 70·5% | −3·9 pp [−6·8, −1·2] | 0·021 | 16:33, p = 0·021 |
| MDT − P | top-1 | 47·2% vs 49·3% | −2·1 pp [−4·8, +0·7] | 0·194 | 21:27, p = 0·471 |
| MDT − P | top-3 | 60·5% vs 61·9% | −1·4 pp [−3·8, +1·0] | 0·106 | 19:24, p = 0·542 |
| MDT − P | top-5 | 66·5% vs 66·5% | −0·1 pp [−2·4, +2·4] | 0·716 | 24:23, p = 1·000 |

Per-seed arm levels (mean ± SD): A×1 51·7 ± 0·2 / 65·6 ± 0·1 / 70·5 ± 0·4; P 49·3 ± 1·5 / 61·9 ± 0·7 / 66·5 ± 1·2; MDT 47·2 ± 0·8 / 60·5 ± 1·3 / 66·5 ± 1·1 (top-1/3/5). Token cost per case: A×1 1,724 (median 1,624); P 2,732; MDT full chain (five role calls plus moderator) 12,606 (median 11,982), 7·3× A×1. Unadjudicated unique pairs: 0.

**Verdict (pre-frozen rule).** A×1 significantly ahead at every endpoint: rule (a), reversal established — the reversal does not depend on symptom-level labels. The three-arm ordering A×1 > P ≥ MDT and the P recall deficit reproduce the original cohort’s structure; the moderated team again adds nothing over plain sampled perspectives on emergency text.

**Cross-stack bridge (pre-registered P5).** Fifty original-cohort cases (case-id-sorted, default_rng(20261101), 50 without replacement) were rerun on the fresh cohort’s local stack — same prompts, temperatures, and seed 1 — and compared with the original-cohort seed-1 outputs on the same cases (S2b-2 reporting convention: magnitudes and directions, descriptive). Arm-level shifts (new stack − original stack) were within noise at every endpoint (all McNemar p > 0·3); the MDT − A×1 estimate moved from −4·0 to −2·0 points at top-3 (shift +2·0, bootstrap CI −16·0 to +20·0) and from −8·0 to −2·0 at top-5 (shift +6·0, CI −6·0 to +18·0), with both stacks negative at top-3 and top-5. With 50 cases and one seed this bridges, rather than tests, the stacks; it bounds the stack contribution to the fresh-cohort estimate well below the effect itself.

**Workup and output-contract sensitivity arms on the fresh cohort (pre-registered P2, P3).** Adding objective results helped every strategy (A×1 +7·2/+7·1/+7·0 points at top-1/3/5; P +7·8/+6·6/+5·9; MDT +5·9/+7·3/+6·6; all p ≤ 0·0002) and the reversal persisted under workup (MDT−A×1 −5·9/−4·9/−4·4, p ≤ 0·0078); the strategy-by-input interaction was null at every endpoint (−1·3/+0·2/−0·4, all intervals spanning zero) — on the disease-label cohort the deficit is input-invariant, unlike the original cohort’s workup interaction. A symptom-permissive output contract (Note S36 clause, byte-identical injection points) narrowed the strict-contract gap at every endpoint (MDT−A×1 −4·5/−5·2/−4·0 strict versus −1·9/−2·5/−1·0 contract; point estimates shrinking 58%/52%/75%, every contract interval spanning zero; gap shift +2·6/+2·7/+3·0, top-5 CI barely excluding zero, p = 0·06), with the symptom-form candidate rate rising as the clause intends (MDT 2·46% → 3·46%). The within-arm response was asymmetric in the direction the forced-differentiation account predicts: the clause left A×1 unchanged (+1·4/+2·5/+1·6 points at top-1/3/5, all p ≥ 0·11) but lifted P (+2·3/+3·0/+3·2, p ≤ 0·015) and MDT (+4·0/+5·2/+4·6, p = 0·0031, 1·8 × 10⁻⁶, and 2·7 × 10⁻⁶ at top-1/3/5) — the same selective rescue of the multi-perspective arms as on the original cohort (Note S36), reproduced here on a cohort whose labels are all disease-level. Read against the S36 pre-committed criteria, the contract closes the ordering deficit on the disease-label cohort descriptively: part of the reversal is an artefact of forced disease guessing under the strict contract, while the pre-registered primary result stands on the strict contract under which all other corpora were evaluated.

**Second-family replication on the fresh cohort (pre-registered P4) and family bridge (P5b).** DeepSeek-V4-Flash-4bit (same local omlx stack, thinking disabled, byte-identical prompts) reproduced the A×1 > P ≈ MDT ordering at every endpoint: MDT−A×1 −6·2/−7·1/−6·5 points (p ≤ 0·0003, majority McNemar concordant), sign-consistent with the Qwen fresh-cohort result (−4·5/−5·2/−4·0) and of larger magnitude; P again trailed A×1 on recall and MDT added nothing over P; the full team chain cost 12,077 tokens per case (7·3× A×1’s 1,646). One serving-level adaptation was required and is disclosed: DeepSeek-V4 verbatim-mimics the escaped {{ }} braces of the P arm’s output-template example (that suffix is concatenated after formatting, so the escapes remain literal), producing unparseable JSON; the parser normalises doubled braces before extraction — prompts and sampling were untouched, and the other arms’ prompts pass through interpolation so were unaffected. The family bridge (the same fixed 50 original-cohort cases, local V4-4bit versus the original cloud v4.1-flash replication, one seed, descriptive) showed arm-level shifts of −4 to −12 points (local below cloud, consistent with 4-bit quantisation) while the MDT−A×1 gap itself shifted +0·0 points at every endpoint (−8·0/−14·0/−14·0 on both stacks): within the second family too, the reversal’s magnitude is serving-invariant even where absolute accuracy is not.

**Judge revalidation on the fresh-400 candidate distribution (P6, first half).** The same two clinicians independently re-rated 100 pairs drawn from the fresh cohort’s adjudicated pairs under the S5-3 protocol verbatim (50 judge-YES / 50 judge-NO by the frozen verdict, boundary pairs — judge vs same-disease heuristic disagreement — prioritised within strata, 63 boundary pairs included, all previously rated pairs excluded, rng seed 77; independent per-string Chinese translations as language aid only, judge verdicts withheld). Agreement reproduced the original-cohort validation almost exactly: clinician–clinician κ 0·849 (raw agreement 0·93; original 0·877/0·94), judge–clinician κ 0·560 and 0·540 (0·78/0·77 raw agreement; original 0·580/0·540), either-rater majority vs judge κ 0·540 (original 0·580). Strata show the same asymmetry as before: judge-YES verdicts were endorsed by 90–92%, judge-NO verdicts by only 62–66% — most judge–clinician discordance is the clinicians reading a pair as the same disease where the judge rejected it. The instrument’s measured reliability on the replacement cohort is therefore unchanged: adequate raw agreement, moderate κ against clinicians whose mutual agreement is substantially higher, and the same reject-side leniency asymmetry. The primary readout inherits exactly the measurement caveat quantified for the original cohort — no worse, and no better (report kappa_report_erf400.json).

**Direction of the residual discordance (post-hoc, descriptive).** Decomposing the discordant pairs shows the judge’s residual error is directional, not random. Clinicians endorsed 20 of the 50 pairs the judge rejected but rejected only 3 of the 50 it accepted (two-sided binomial p = 0·0005); the same direction held in the post-clarification original-cohort validation (20 versus 2, p = 0·0001) and was null in the S5-3 rerun batch (8 versus 13, p = 0·38), pooling to 48 versus 18 (p = 0·0003) across the three valid emergency batches. Qualitatively, the rejected-but-endorsed pairs are predominantly concurrent or aetiologic candidates that encompass the reference within a richer diagnostic bundle — for example “hepatic encephalopathy with secondary seizures” against the reference “seizure”, and “normal pressure hydrocephalus secondary to prior glioblastoma resection” against “glioblastoma”; 16 of the 20 lexically contain the reference’s core string — which clinicians read as containing the correct diagnosis where the frozen v3 rules, keyed to the candidate’s head entity, reject it. Because the validation sample is drawn from unique judged pairs with judge-verdict stratification and boundary enrichment, its arm composition reflects each arm’s candidate-string pool (MDT’s pool is 2·4 times A×1’s), so per-arm comparisons within the sample are uninformative. A full-corpus check on all unique fresh-400 pairs finds no arm differential: candidates lexically containing the reference core string were rejected at 8·8% (A×1, 14/160), 4·3% (P, 14/322), and 3·4% (MDT, 12/348), and aetiologic/concurrent-style containing candidates judged NO numbered 7 (A×1), 8 (P), and 8 (MDT). The judge is therefore stricter than clinicians overall — most visibly on encompassing candidates — and on this false-reject channel the differential is statistically against the single call (8·8% vs 3·4%, 14/160 vs 12/348, two-proportion z = 2·52, p = 0·012): residual judge error of this kind biases the measured MDT−A×1 gap toward the null, so the reversal is, if anything, underestimated. Per-arm false-accept rates remain unestimable without bias, so arm-dependent misclassification is bounded, not excluded. This analysis is descriptive (judge-stratified sample; no unbiased per-arm false-reject rates are implied) and does not alter any frozen readout (pair-level data erf400_discordance_direction.json).

**Blinded physician rating of the fresh-400 discordant pairs (P6, second half; protocol v1.3).** The safety assessment was repeated on the replacement primary cohort under a revision (v1.3, frozen 2026-11-05 before any rating material existed) that inverted the original rating’s question structure. Motivation: the original rating (Note S37) covered the original cohort, and its danger-construct agreement collapse (AC1 0·185) indicated that pairwise danger comparison may carry little ratable signal on this material; the output-contract arm (Note S36) and the discordance-direction analysis above also showed that strict-contract discordant pairs mix in many granularity variants. The revision therefore made the primary question case-level mechanism verification (rescue) and retained the danger comparison as secondary, with structured operational anchors (dangerous omission = an omitted diagnosis that, unmanaged within the ED course, could plausibly cause serious harm; misleading inclusion = a high-ranking entry whose pursuit could directly drive wrong management; S1 scored by a most-severe-consequence × time-window rubric with worked anchor examples).

**Sampling frame (frozen; sampling rng 20261105).** All 53 fresh-400 cases whose top-1 five-seed majority verdicts disagreed between A×1 and MDT (35 A×1 wins, 18 MDT wins — exactly the battery’s McNemar cells); 30 concordant-hit controls (pool 172) for the side-bias check; 15 concordant-miss baselines (pool 175) as the rescue-rate background; a 10-unit calibration batch (excluded from all analyses); and 10 main-batch cases embedded twice (14 duplicate units) for intra-rater reliability — 175 units in total. The sampling-frame file and randomisation sequence were hash-archived at rating-material generation (MANIFEST.sha256).

**Design (two parts, order frozen).** Part A (list-level, blind to the reference label): case plus two anonymised seed-1 top-5 lists in random A/B order, rated on S1 (danger comparison, primary-secondary), S2 (per-list dangerous-omission and misleading-inclusion flags), and D5 (initial-differential preference). Part B (case-level, after the reference was revealed): the losing arm’s anonymised seed-1 list plus the reference diagnosis, answered on whether the list contained an acceptable entry encompassing the correct diagnosis — including undifferentiated-but-pointing formulations, superordinate concepts, or aetiologic-chain statements — Y/N plus a classification (encompassing / related but insufficient / unrelated). Concordant-miss baselines showed one randomly chosen arm’s list under the same question. The calibration batch ran first; after it, raters received a written anchor-interpretation document with expert-consensus worked examples; calibration scores entered no analysis. Three non-author senior physicians (≥1 emergency medicine), blind to strategy, hypotheses, and each other, rated on a local server; case text did not leave the rating environment.

**Pre-declared deviations and design constraints.** (i) Sampling followed the majority vote while display used seed-1 lists (the S37-type discrepancy); in 8 of 83 Part A main units the displayed seed-1 list was inconsistent with the majority-vote frame, disclosed as a pre-declared design deviation. (ii) Rescue judgements in Part B were made after reference revelation, in a different context from the judge’s adjudication; the two are not directly comparable on agreement statistics. (iii) Part B arm anonymity was partially inferable. (iv) Raters could partially infer arm attribution from Part A. Power disclosure (pre-committed): with n = 35 and n = 18, the rescue-rate contrast had 80% power only for differences of roughly 35–40 points.

**Table S39b. Reliability before unblinding (175 units; three raters).**

| Measure | Pairwise Gwet AC1 (three pairs) | Mean | Gate (≥0·6) |
| --- | --- | --- | --- |
| Intra-rater (14 embedded duplicates each) | 14/14, 14/14, 14/14 exact | — | passed |
| S1 danger comparison (Part A) | 0·77 / 0·96 / 0·78 | 0·836 | passed |
| S2 dangerous omission (Part A) | 0·74 / 0·96 / 0·76 | 0·819 | passed |
| S2 misleading inclusion (Part A) | 0·99 / 1·00 / 0·99 | 0·992 | passed |
| D5 initial-differential preference (Part A) | 0·30 / 0·72 / 0·38 | 0·467 | **missed → descriptive** |
| B1 rescue judgement (Part B) | 0·76 / 0·85 / 0·86 | 0·822 | passed |

The structured anchors markedly improved agreement over the original rating (S37 danger AC1 0·185 → 0·836); only D5 remained below the gate and that family is reported descriptively, per the frozen rule.

**Table S39c. Q1 (primary): rescue rate of the losing arm’s list (majority verdict over three raters; denominator = discordant cases lost by that arm).**

| Losing arm | Rescued | Rescue rate | Newcombe 95% CI |
| --- | --- | --- | --- |
| MDT (n = 35) | 29 | 0·829 | — |
| A×1 (n = 18) | 18 | 1·000 | — |
| Difference (MDT-lost minus A×1-lost) | — | −0·171 | −0·327 to +0·026 |

Pre-committed verdict rule: interval excludes 0 and positive → mechanism validated; includes 0 → unadjudicable; negative → refuted. The interval includes 0: **unadjudicable**. The point direction (A×1-lost lists rescued more often than MDT-lost lists) is opposite to the forced-differentiation hypothesis. Background: the concordant-miss baseline rescue rate was 6/15 (0·40) — both arms’ lost lists were rescued far above the background rate, so the judge’s misses on both arms were largely not clinical omissions; the supported positive reading is that judge strictness falls on both arms, while the output contract is the arm-specific moderator.

**Table S39d. Q2 (secondary): danger comparison on the 53 discordant pairs (majority verdict).**

| Measure | Result | 95% CI / test |
| --- | --- | --- |
| S1: “MDT errors more dangerous” among non-tied units | 3/11 (0·273) | exact CI 0·060–0·610; two-sided binomial p = 0·23 (direction opposite to S37’s descriptive lean) |
| S2 dangerous omission: only-A×1 vs only-MDT | 8 : 3 | exact McNemar p = 0·23 |
| S2 misleading inclusion: only-A×1 vs only-MDT | 0 : 0 | p = 1·00 |
| Side-bias check (concordant-hit controls): MDT preference | 0/4 non-tied | no side preference |

All secondary contrasts are non-significant; no safety-equivalence or superiority statement is made (pre-excluded by the frozen protocol).

**Artifacts and disclosure.** Sampling frame and hashes (frame.json, MANIFEST.sha256), unit builder, rating interfaces (erf400_rater.html, erf400_rater_en.html, English version used), analysis script (analyze_erf400_rating.py, hash-archived pre-unblinding in ANALYSIS.sha256), and the three rater exports live under clinical_rating/erf400_rating/; the rating units and rater responses contain ER-Reason-derived case text and remain local under the data use agreement, scheduled for deletion after submission confirmation. Protocol revision v1.3 is part of paper/er_fresh_protocol_20261101.md; ethics coverage follows the existing exemption (No. I-26ZM0145).

**Artifacts.** Protocol paper/er_fresh_protocol_20261101.md; builder build_erfresh_subset.py and audit table erfresh_label_audit.md; generator and progressive judge erfresh_gen.py / erfresh_judge.py; per-seed outputs results/erfresh_omlx/ (primary), erfresh_workup/, erfresh_contract/, erfresh_ds/, erfresh_bridge/, erfresh_ds_bridge/; adjudication shard judge_shard_erfresh.json; analyses fresh400_analysis.py, erbridge_analysis.py, and fresh400_battery_analysis.py with reports fresh400_primary.{md,json}, erfresh_bridge.{md,json}, fresh400_battery.{md,json}, and the discordance-direction analysis judge_validity/erf400_discordance_direction.json under routing_study/results/. ER-derived outputs remain outside the release tree under the data use agreement.

### Archived superseded text (separate file, not part of this supplement)

The verbatim archive of the pre-condensation manuscript (Supplementary Notes S18–S33, Round 6) is provided as a separate file (supplement_archive_separate.md) to keep the peer-reviewed supplement free of superseded numerals. Its framework statements (pooled-seed McNemar counts, pre-rerun emergency numbers, the earlier 15-of-16 multiplicity tally) are superseded by Tables S3, S4, and S6 above.
